# International Multi-site Implementation of Local Cell-Free Protein Biomanufacturing to Advance Health and Research Equity

**DOI:** 10.1101/2025.07.25.25332228

**Authors:** Severino Jefferson Ribeiro da Silva, Quinn Matthews, Séverine Cazaux, Justin R. J. Vigar, Bárbara N. R. Santos, Deyse C. M. Carvalho, Lauren A. Cranmer, David Duplat, Serena Singh, Mohammad Simchi, Paula Benítez-Bolivar, Thaíse Yasmine Cavalcanti, Kaiyue Wu, Renata P. G. Mendes, Tanvi Kale, Ana Luisa Lot Divarzak, Krištof Bozovičar, Jennifer Doucet, Anibal Arce, Cielo Léon, Valentina Ferrando, Jessica Nguyen, Anson Ho, Suelen Cristina de Lima, Pouriya Bayat, Yuxiu Guo, Seray Cicek, Aidan Tinafar, Larissa Krokovsky, Laís Ceschini Machado, Ashyad Rayhan, Idorenyin A. Iwe, Alexander Klenov, Thiago P. G. de Araujo, Jean Phellipe M. do Nascimento, Jurandy J. F. de Magalhães, Marcela Guevara-Suarez, Patricia García Canete, Kodjo Ayi, Moiz Charania, Marcelo H. S. Paiva, Ian Crandall, Tony Mazzulli, Gabriel da Luz Wallau, Abelardo Silva-Júnior, Adriana Bernal, Chaitanya Athale, Alexander A. Green, Scott C. Weaver, Camila González, Fernán Federici, Lindomar Pena, Keith Pardee

**Affiliations:** Department of Pharmaceutical Sciences, Leslie Dan Faculty of Pharmacy, University of Toronto, Toronto, Ontario, Canada; ANID - Millennium Science Initiative Program - Millennium Institute for Integrative Biology (iBio), Santiago, Chile; Institute for Biological and Medical Engineering, Schools of Engineering, Medicine and Biological Sciences, Pontificia Universidad Católica de Chile, Santiago, Chile; Department of Virology, Aggeu Magalhães Institute (IAM), Oswaldo Cruz Foundation (Fiocruz), Recife, Pernambuco, Brazil; Department of Microbiology and Immunology, University of Texas Medical Branch, Galveston, TX, USA; Centro de Investigaciones en Microbiología y Parasitología Tropical (CIMPAT), Department of Biological Sciences, Universidad de los Andes, Bogotá, Colombia; Department of Biomedical Engineering, Boston University, Boston, MA, USA; Division of Biology, IISER Pune, Dr. Homi Bhabha Road, Pashan, Pune 411008, India; Department of Chemical and Biological Engineering, Northwestern University, Evanston, IL 60208, USA; Department of Biophysics and Radiobiology, Federal University of Pernambuco (UFPE), Recife, Pernambuco, Brazil; LSK Technologies Inc., Kitchener, Ontario, Canada; Liberum Biotech Inc., Kitchener, Ontario, Canada; Department of Entomology, Aggeu Magalhães Institute (IAM), Oswaldo Cruz Foundation (Fiocruz), Recife, Pernambuco, Brazil; Institute of Biological and Health Sciences, Federal University of Alagoas (UFAL), Maceió, Alagoas, Brazil; University of Pernambuco (UPE), Serra Talhada Campus, Serra Talhada, Pernambuco, Brazil; Public Health Laboratory of the XI Regional Health, Pernambuco, Brazil; Applied genomics research group, Vicerrectoría de Investigación y Creación, Universidad de los Andes, Bogotá, Colombia; Department of Clinical Laboratories, School of Medicine, Pontificia Universidad Católica de Chile, Santiago, Chile; Life Sciences Center, Federal University of Pernambuco (UFPE), Academic Center of Agreste, Caruaru, Pernambuco, Brazil; Department of Microbiology, Sinai Health System/University Health Network, Toronto, M5G 1X5, ON, Canada; Bioinformatics Core, Aggeu Magalhães Institute (IAM), Oswaldo Cruz Foundation (Fiocruz/PE), Brazil; Department of Arbovirology and Entomology, Bernhard Nocht Institute for Tropical Medicine, Hamburg, Germany; Federal University of Santa Maria (UFSM), Santa Maria, Rio Grande do Sul, Brazil; Laboratory of Molecular Interactions of Agricultural Microbes (LIMMA), Department of Biological Sciences, Universidad de Los Andes, Bogotá, Colombia; Molecular Biology, Cell Biology & Biochemistry Program, Graduate School of Arts and Sciences, Boston University, Boston, MA 02215, USA; Biological Design Center, Boston University, Boston, MA 02215, USA; World Reference Center for Emerging Viruses and Arboviruses, University of Texas Medical Branch, Galveston, TX, 77555, USA; Institute for Human Infections and Immunity, University of Texas Medical Branch, Galveston, TX, 77555, USA; Department of Mechanical and Industrial Engineering, University of Toronto, Toronto, Ontario, Canada

## Abstract

Limitations in global access to research and healthcare capacity undermine equity, sustainability, and resilience, particularly in resource-limited settings. Molecular diagnostics and biologic therapeutics are set to revolutionize medicine; however, our dependence on centralized biomanufacturing, and the concomitant cold chain logistics, restrict access to these benefits. While these constraints become particularly evident during global health crises, they reflect a chronic and unmet global challenge. Here, with research teams in North and South America and Asia, we challenge the conventional top-down paradigm by innovating community-driven solutions that empower underserved populations to actively participate in the bioeconomy, producing what they need, when, and where they need it. Our approach leverages decentralized, low-burden biomanufacturing technologies—built on cell-free protein synthesis and open-source hardware—to enable local, on-demand production of critical biologics, including high-value growth factors, vaccines, and diagnostic enzymes, demonstrating performance comparable to commercial gold standards. This platform, implemented at ten sites worldwide, supported patient trials targeting globally relevant pathogens, including SARS-CoV-2, chikungunya, and Oropouche viruses. Together, these initiatives lay the foundation for a new era of globally inclusive biomanufacturing, where innovation goes beyond geographic boundaries, and communities everywhere are empowered to respond to global challenges with enhanced speed, autonomy, and equity.

## MAIN

Emerging biotechnologies hold transformative potential to strengthen economic and health security, while benefiting the planet. Advances in precision medicine, molecular diagnostics, and synthetic biology are driving improvements in global health outcomes, reducing healthcare costs, enhancing food security, and expanding equitable access to care(*1–5*). Meanwhile, biotechnology innovations are fueling economic growth by creating new industries and employment opportunities. Additionally, biotechnology supports environmental sustainability through cleaner and more efficient solutions, such as designer enzymes, microbes, and crops, helping to preserve the planet’s health for future generations(*3, 6–8*).

Yet, the current limited and uneven distribution of biotechnology capacity exacerbates existing inequities, particularly between high-income and low- and middle- income countries (LMICs). Access to advanced tools, such as molecular diagnostics, life- saving treatments, and biomanufacturing infrastructure, remains largely concentrated in wealthier regions, thereby restricting access to transformative solutions for communities that need them the most. Notably, one of the main drivers behind this global challenge is the reliance on centralized production systems, which require sophisticated, capital-intensive infrastructure and cold supply chains that are often unavailable in resource-limited settings (**Fig. 1a**)(*9, 10*).

**Fig. 1:**
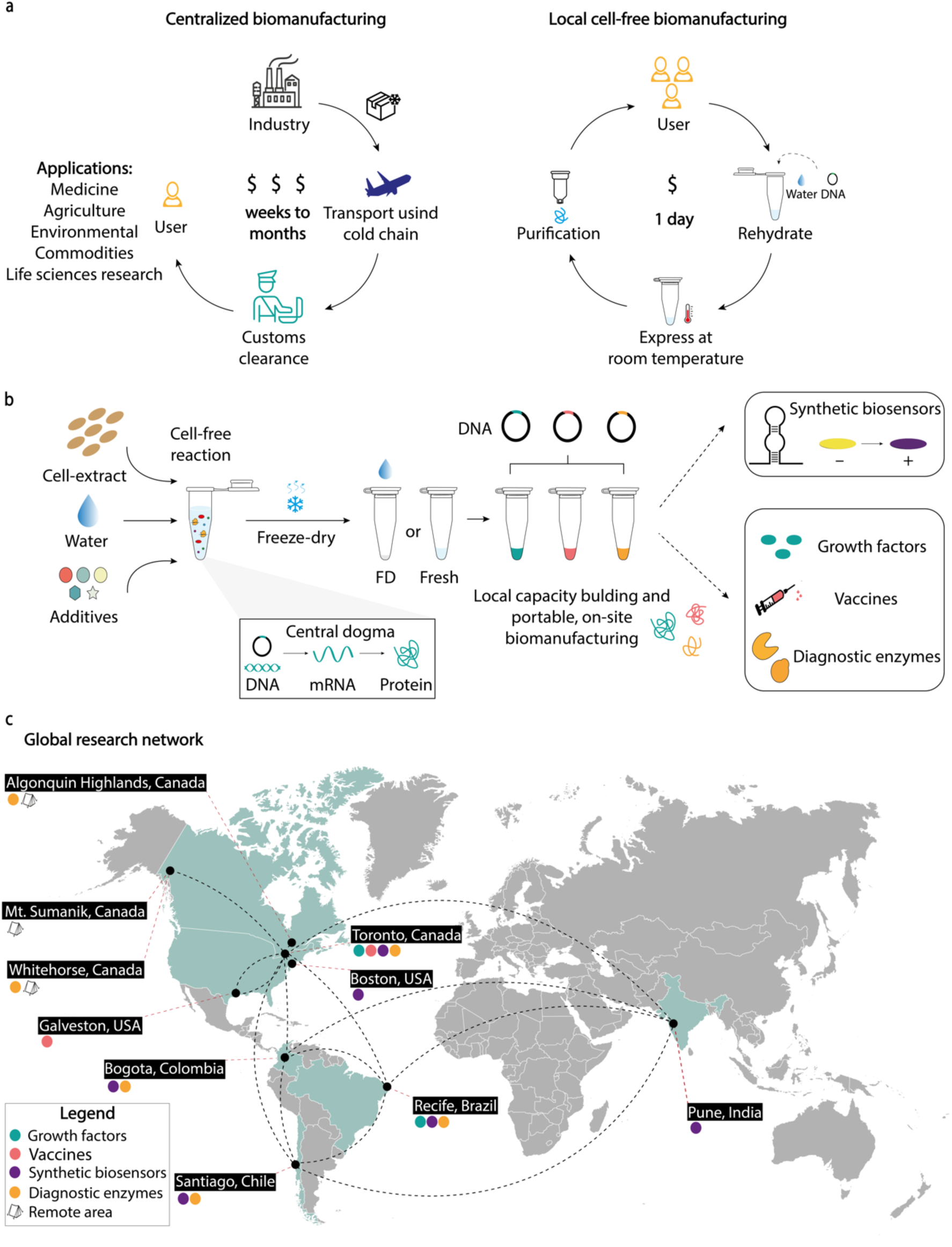
Decentralizing biomanufacturing enables equitable biotechnology access by enhancing local capacity. **(a)** Traditional centralized biomanufacturing approaches rely on costly, time-intensive workflows that require cold-chain logistics and customs clearance, limiting access to biotechnology in resource-limited settings. Although this centralized biomanufacturing model is well-established and offers significant benefits to those who can afford them, in the form of consistent product performance with quality control, the high cost to lower resourced nations is limiting, especially during global events of supply chain disruptions, like in public health emergencies, natural disasters, and military conflicts. This was particularly evident during the COVID-19 pandemic when countries in the Global South struggled to obtain essential diagnostic reagents, highlighting the fragility of dependence on centralized biomanufacturing systems. Moreover, users in the Global South often face 1- to 4-month delays in receiving bioreagents, which cost two to three times more than in countries in the Global North. Cell-free biomanufacturing, in contrast, enables decentralized, rapid, and cost-effective on-site production of bioreagents—including growth factors, vaccines, biosensors, and diagnostic enzymes—accessible to any user within one day using FD-CFPS reaction mixtures and DNA templates. This synthetic biology platform provides scalable molecular biosynthesis for global health, industry, research, and education. **(b)** Schematic of the cell-free reaction process illustrating how fresh or freeze-dried lysates, combined with molecular instructions (DNA template) and essential molecules, enable transcription and translation to produce the desired product. This can enhance global biotechnology capacity building by enabling on-site production of diagnostic and therapeutic tools with minimal infrastructure. Looking ahead, we envision a system that combines FD-CFPS and on-site biomanufacturing, enabling rapid and reliable diagnostics to monitor patients for infection and bedside therapeutic production for real-time patient care. **(c)** Our global research network demonstrates the feasibility of driving decentralized biomanufacturing across diverse regions and settings. Collaborative efforts span the Global North and Global South; we establish the reproducibility and scalability of cell-free expression and low-cost purification platforms for producing high-value bioproducts. Experimental locations include Canada, the USA, Chile, Brazil, Colombia, India, and three geographically distant sites in Canada, highlighting the role of international partnership in this study.

The repercussions of access inequality are particularly evident in healthcare, where disparities in biomanufacturing capacity and biologics availability disproportionately impact underserved regions, marked by slow diagnostic deployment, delayed disease control programs, uneven resource distribution, and limited local capacity for fundamental life sciences research(*11–15*). Outbreaks of viruses such as Zika, chikungunya, and Oropouche outbreaks in Latin America, alongside the COVID-19 pandemic, have highlighted these vulnerabilities(*16–20*). These combined factors, along with real-world logistical challenges such as customs regulations, prolonged shipping delays, import restrictions, and taxes, underscore the urgent need for local, decentralized manufacturing to help LMICs build capacity in biotechnology.

By prioritizing the development of accessible scientific capacity, we can establish research systems that are resilient, adaptable, and locally sustainable, enabling the scaling of effective solutions on a global scale(*21–23*). One promising avenue to improving access is cell-free protein synthesis (CFPS), which offers the potential to empower communities through affordable, low-burden, on-site production of critical bioreagents, diagnostic tools, and therapeutic agents(*24–36*). CFPS operates without living cells, using biomolecular machinery from crude cell lysates or reconstituted components to synthesize RNA and proteins of interest(*37*). Freeze-dried, activated by simply adding water, they can be stored, and distributed at ambient temperature, easily used by scientists and non-scientists alike (**Fig. 1b**)(*25, 38*). Their incorporation into the biomanufacturing ecosystem can add technical diversity and augment the capacity of centralized manufacturing hubs, while reducing challenges in LMICs with cold-chain logistics, customs delays, and cross-border regulatory barriers(*25, 39*).

Indeed, the World Health Organization (WHO) and the United Nations (UN) have recognized the great potential of CFPS and stressed the importance of decentralized biomanufacturing and capacity building in biotechnology(*40*). This was echoed recently at the Conference of the Parties (COP16) to the UN Convention on Biological Diversity(*40*), and the WHO emphasized that the inequity in access is unacceptable(*41*).

To address the urgent need for expanded global capacity in biotechnology, we report an international effort to implement and validate decentralized cell-free biomanufacturing (**Fig. 1c**). We begin by establishing a technology transfer network for standardized and reproducible CFPS production across five countries spanning North America, South America, and Asia. This molecular capacity is paired with low-burden, companion hardware, including a 3D-printed hand-powered centrifuge and an open-source diagnostic reader, as alternatives to capital-intensive laboratory infrastructure.

Next, we demonstrate that decentralized biomanufacturing, implemented through our standardized framework, expands access to essential and high-value biologics. We first demonstrate the versatility of CFPS-based biomanufacturing with the production and validation of a repertoire of 11 growth factors commonly used in mammalian cell culture and relevant to regenerative medicine and stem cell-based therapies. We then establish vaccine production capacity—critical for areas with limited healthcare access—by producing a COVID-19 subunit-based vaccine candidate that elicits a robust immune response when administered to an animal model. The capacity for vaccine production is then paired with a wide-ranging demonstration of the potential for the local manufacture of molecular diagnostics in diverse settings, from national reference diagnostic laboratories to resource-constrained environments. Here, with the demonstration of on-site enzyme production within a single day, we establish disease diagnostic programs for 16 clinically relevant pathogens, including the emerging tropical pathogen of concern Oropouche virus, linked to neuroinvasive disease and congenital anomalies such as microcephaly(*42*), and highly pathogenic H5N1 virus. Of these, the diagnostics for SARS-CoV-2, chikungunya, and Oropouche are validated through patient trials (116 samples) across four countries in the Global North and the Global South, achieving 90%–100% accuracy in comparison to the gold-standard RT-qPCR. Prioritizing accessibility, affordability, and reproducibility, we show that cell-free biomanufacturing is a transformative tool for expanding global health equity and democratizing participation in the bioeconomy.

## RESULTS

### Building tools and systems for effective technology transfer and deployment

To establish multi-site biomanufacturing capacity across an international network of collaborating laboratories, we validated the stability, transferability, reproducibility, and practical use of CFPS systems in diverse geographic regions, spanning both resource-rich and resource-limited environments. First, using freeze-dried CFPS (FD-CFPS) shipped at ambient temperature between sites, we standardized reaction preparation protocols across five laboratories in Canada, Chile, Brazil, Colombia, and India. These sites span latitudes from 43°N to 33°S and represent a range of laboratory capabilities and shipping conditions, with variation in transport time, temperature, and humidity. Here, we employed superfolder green fluorescent protein (sfGFP) as a model protein product and used fluorescein isothiocyanate (FITC) standard curves to ensure reproducibility between laboratories despite variable equipment, providing an essential on-site Quality Control (QC) step **(Supplementary Notes 1,2)**.

Consistent with previous studies(*25, 26, 38, 43*), our results revealed that FD-CFPS reactions could be stored at ambient temperatures (e.g., 22–24 °C) for at least two weeks with no significant loss of activity (**Fig. 2a**, **Fig. S1)**, allowing for global shipping without cold chain logistics (**Fig. 2b**). After normalization, sfGFP fluorescence exhibited an average coefficient of variation (CV) of only ∼10 % (SD ± 2.68) across all five sites (n = 5) **(Fig. S2)**. Together, this established a standardized, globally transferable CFPS workflow, enabling bioproduction across the international team.

**Fig. 2:**
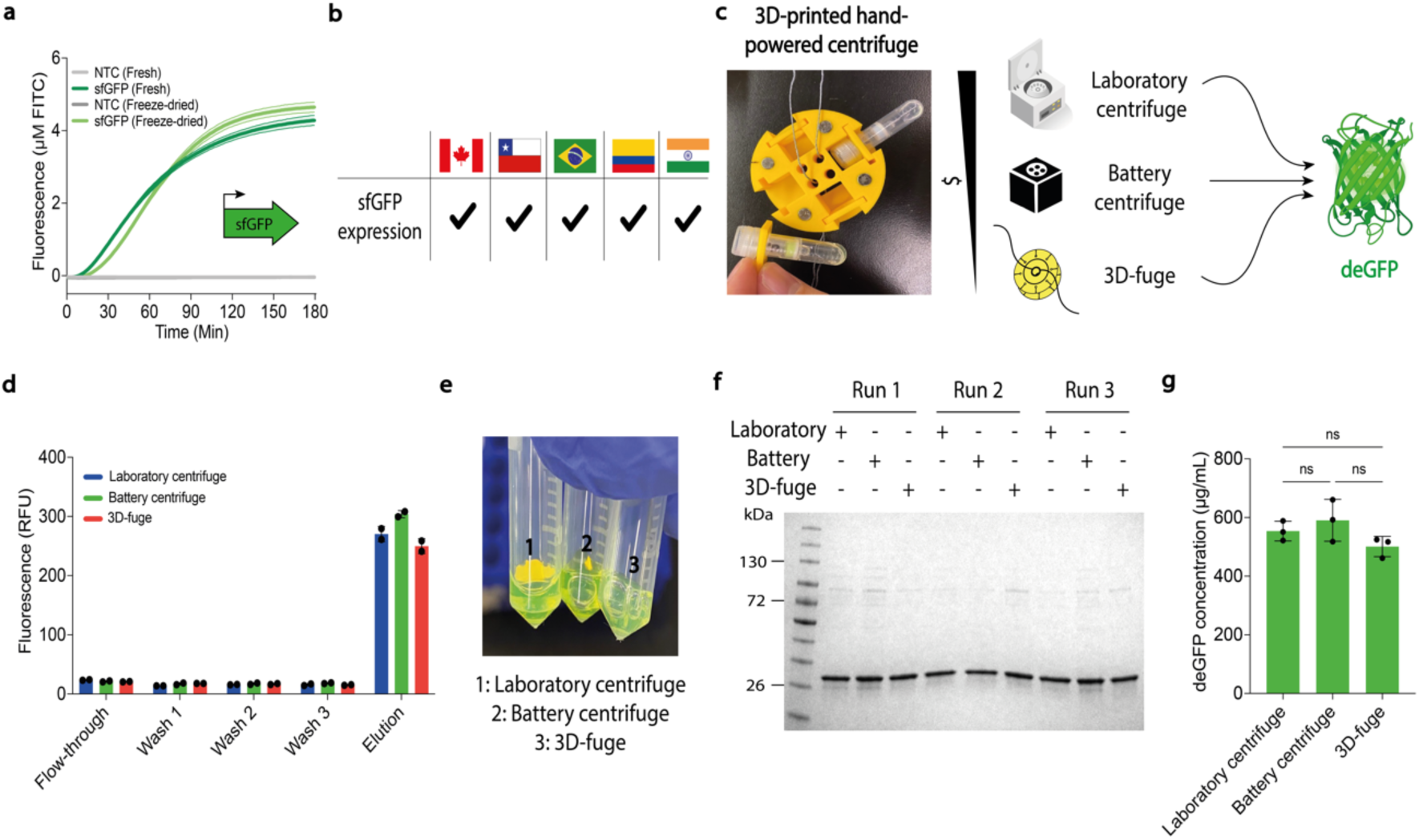
Standardized protocols and portable CFPS systems and labware enable intercontinental distribution to support the decentralization of biotechnology. **(a)** CFPS activity was assessed using sfGFP fluorescence standardized to FITC curves. FD-CFPS reactions stored at ambient temperature are compared to fresh reactions. In this representative experiment, sfGFP fluorescence was measured every ∼2 minutes for 3 h using cell-free lysates produced on-site in Chile. **(b)** FD-CFPS reactions were shipped (from Chile) without cold-chain logistics to researchers in Canada, Brazil, Colombia, and India, who could synthesize sfGFP upon rehydration. **(c)** Different centrifuges used to affinity-purify deGFP. The photograph shows an in-house designed, low-cost, open-source 3D-printed hand-powered centrifuge. In this representative experiment, deGFP was produced using cell-free lysates (prepared in Canada) and transported to Colombia at ambient temperature, where the experiments were conducted. **(d)** deGFP fluorescence was tracked during purification protocols using three different centrifuges and a standard plate reader. Data are shown as mean ± SD, n = 3. **(e)** Photographs of purified deGFP all showed a vivid green color. **(f)** Purified deGFP (MW = ∼ 27 kDa) from all three purification methods was analyzed via 4- 20% gradient SDS-PAGE in biological triplicate. The molecular weight ladder (in kilodaltons) is shown on the left. **(g)** Total soluble protein levels were quantified using a bicinchoninic acid (BCA) kit, showing similar yields across methods. Data are shown as mean ± SD of biological replicates, n = 3. Statistical differences were determined by one-way ANOVA with Tukey’s post hoc multiple comparisons test: ns p > 0.05. Abbreviations are: ns, not significantly different; NTC, non-template control.

All protein expression was performed using CFPS(*44*) reactions prepared from widely available *E. coli* (BL21[DE3] or Shuffle) strains, depending on the application. Crude lysates were produced locally in basic low-containment microbiology laboratories at sites in North or South America, using inexpensive inputs such as peptone and yeast extract. Here, CFPS reactions were either used at the production site or freeze-dried and shipped to team members across the globe.

Recognizing that biologics produced through CFPS typically require purification for downstream applications, we next addressed the need for affordable and portable instruments to support this process. Centrifugation-based affinity chromatography of hexa-histidine [His_6_] or streptavidin-tagged products can be performed using readily available resin-packed microcentrifuge columns to provide a simple and low-burden strategy for purification. To explore the practicality of this approach, we systematically tested three devices: a conventional benchtop laboratory centrifuge (∼$10,000 USD), a low-cost commercial battery-powered centrifuge ($149 USD, SpiniOne 2020 Portable Centrifuge), and our in-house designed 3D-printed hand-powered centrifuge ($3 USD, referred to as the 3D-fuge)(*45*) (**Fig. 2c**, **Fig. S3, Table S1).**

After a week at ambient temperature, FD-CFPS reactions (1 mL) were rehydrated with template DNA encoding His_6_-tagged deGFP and incubated for 16 hours overnight (24 °C). Reactions were then divided into three equal parts for deGFP purification using Ni-NTA resin columns with three different centrifugation strategies. Tracking fluorescence levels of the eluate and eluent throughout the purifications, we found similar performance across methods (**Fig. 2d**). The quality of the purified product was then analyzed using fluorescence measurements, electrophoretic analysis, and a colorimetric-based quantification assay, which confirmed consistent protein yields (CV: 8.18%, mean: 548.28 µg/mL eluate, and SD ± 44.86) and purity above 90 % across all purification strategies (**Fig. 2e-g**, **Fig. S4**).

Having established the concept of cell-free biomanufacturing in controlled laboratory settings, we next set out to demonstrate its potential for enabling on-site protein production in the absence of conventional laboratory infrastructure. FD-CFPS reaction mixtures (prepared in Toronto, Canada) were transported 250 km north to a rustic location in the Algonquin Highlands (Ontario)—selected to simulate remote conditions—and to 4,600 km north to Whitehorse (Yukon) in Northern Canada, where bioreagent supply chain constraints limit the capacity for research and teaching **(Fig. S5a,b).** Equipped with only essential supplies, all easily stowed in a backpack like a “lab-in-a-box”, FD-CFPS reactions were rehydrated with template DNA encoding His_6_-tagged deGFP, incubated overnight at ambient temperature (22–24 °C), and purified using the battery-powered and 3D-printed hand-powered centrifuges. Consistent with our previous findings, deGFP expression was confirmed through fluorescence at each temporary research site **(Fig. S5c-f)** and later verified in the laboratory using electrophoresis and a colorimetric-based protein quantification assay **(Fig. S5g).** To demonstrate mobile deployment under extreme conditions, deGFP expression and purification were also carried out under austere conditions during a helicopter flight and hike to Sumanik ridge (Yukon) in Northern Canada **(Fig. S5h-m)**. These findings demonstrate that local high-yield, high-purity protein production can be achieved in diverse settings using low-burden FD-CFPS coupled with either standard laboratory infrastructure or minimal, low-cost tools.

### Local production yields growth factors with commercial-grade bioactivity

Growth factors are signalling proteins that play an essential role in both normal physiology and disease processes(*46, 47*). Accordingly, they are utilized throughout life sciences research(*48*), stem cell therapy(*49*), cell-cultured meat production(*50*), and as therapeutics to promote regeneration(*51*) and treat cancer, autoimmune, and rare diseases(*52–55*). Growth factor manufacturing has traditionally been centralized, relying on capital-intensive downstream processing and cold chain distribution. These requirements create barriers to accessing these biologics in low-resource contexts, where mammalian cell culture can add significantly to existing local research capacity. Through this lens, we sought to develop a pipeline that would allow for on-site production and *in vitro* validation of these typically costly bioproducts (**Fig. 3a**).

**Fig. 3:**
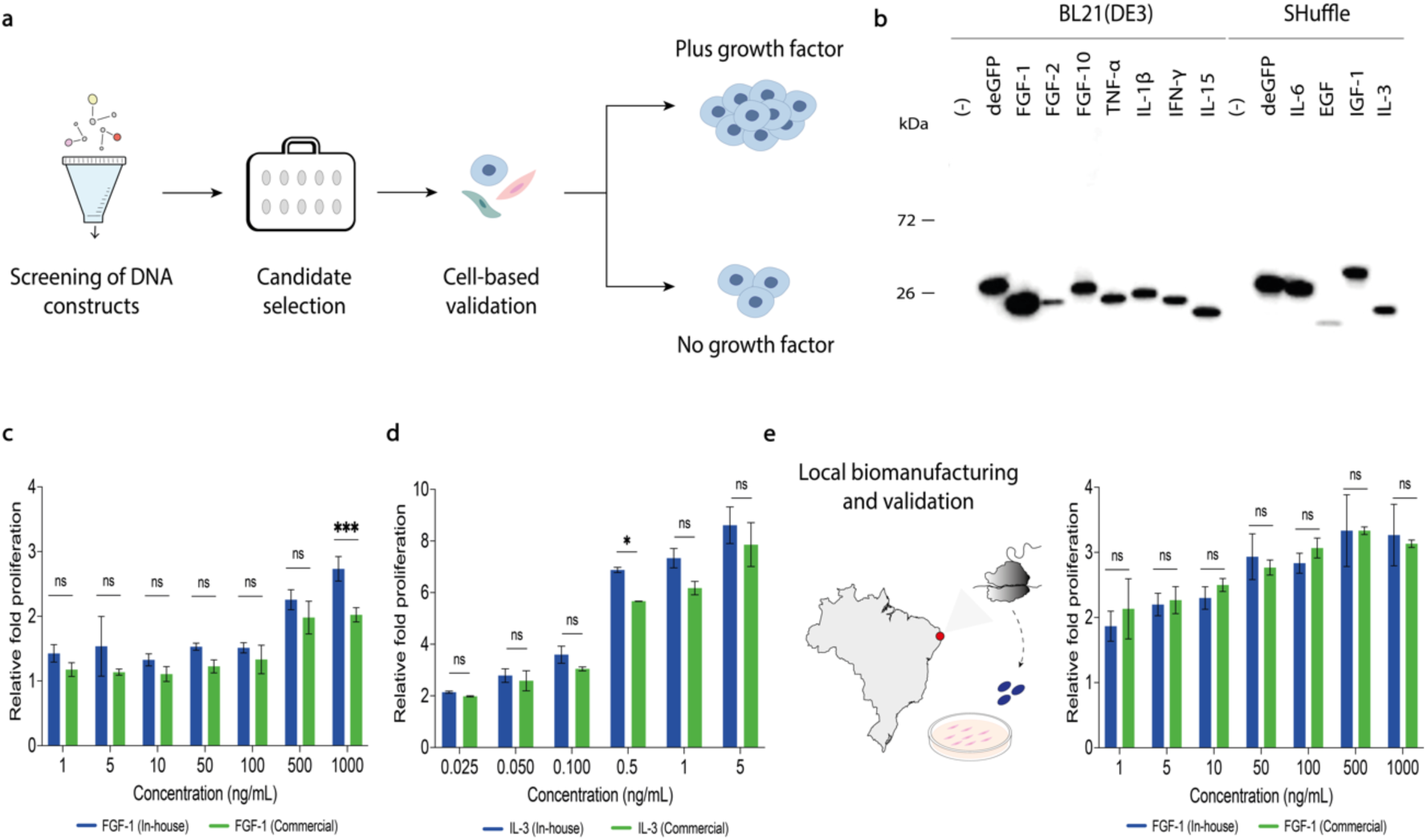
A CFPS manufacturing platform enables local production of functional growth factors. **(a)** Schematic representation of the pipeline for growth factor candidate selection and cell-based testing. **(b)** Western blot analysis of 11 high-value CFPS-derived growth factors. FGF-1, FGF-2, FGF-10, TNF-α, IL-1β, IFN-γ, and IL-15 were expressed in *E. coli* BL21 (DE3) in-house cell-free lysates. IL-6, EGF, IGF-1, and IL-3 were expressed in SHuffle-based in-house cell-free lysates. Detection was performed using an anti-His- HRP antibody. Data shown are from one representative biological replicate out of three independent experiments. **(c)** *In vitro* proliferation assay comparing on-demand, locally produced in Canada (blue) and commercial (green) FGF-1 growth factors in NIH-3T3 cells. Cells were individually treated with varying concentrations (1, 5, 10, 50, 500, and 1000 ng/mL) of growth factor. Luminescence was measured using a conventional plate reader. Data are shown as mean ± SD, n = 3. **(d)** *In vitro* proliferation assay comparing on-demand, locally produced (blue) and commercial (green) IL-3 growth factors in TF-1 cells. Cells were individually treated with varying concentrations (0.025, 0.05, 0.1, 0.5, 1, and 5 ng/mL). This representative data was obtained using reagents produced on-site in Canada. Data are shown as mean ± SD, n = 3. **(e)** Growth factor expression and cell-based validation in a low-resource setting. *In vitro* proliferation assay using FGF-1 and NIH-3T3 cells. Cells were individually treated with varying concentrations (1, 5, 10, 50, 500, and 1000 ng/mL) of both in-house-produced (blue) and commercial (green) growth factors. This representative data was obtained using reagents produced on-site in Brazil. Relative fold proliferation was plotted as the fold-change compared to untreated, serum-starved cells under the same experimental conditions. Data are shown as mean ± SD, n = 3. Statistical differences were determined by two-way ANOVA with Šídák’s post hoc multiple comparisons test: ns p > 0.05, * p < 0.05, ** p < 0.01, *** p < 0.001. Abbreviations are: ns, not significantly different; kDa, kilodaltons.

We began by designing DNA linear templates encoding 11 high-value growth factors, selected for their therapeutic and clinical significance(*52, 53, 56*): fibroblast growth factor 1 (FGF-1), FGF-2, FGF-10, tumor necrosis factor alpha (TNF-α), interleukin- 1 beta (IL-1β), IL-3, IL-6, IL-15, interferon-gamma (IFN-γ), epidermal growth factor (EGF), and insulin-like growth factor 1 (IGF-1) **(Table S2)**. CFPS was first optimized under small-scale conditions (10 μL reactions, 21 °C, 16 hours), with expressions confirmed by Western blot analysis. As expected, the position of the His_6_-tag (N- or C-terminal) significantly impacted protein expression, with the placement, in some cases, leading to reduced or even complete loss of expression **(Fig. S6, Supplementary Data 1)**. Fortunately, with top-performing expression templates identified, all 11 growth factors were successfully expressed using in-house, lysate-based CFPS from BL21(DE3) or SHuffle *E. coli* strains, the latter being selected to enhance disulfide bond formation, along with a fusion protein strategy (e.g., SUMO protein), when required (**Fig. 3b**)(*36, 57*).

To assess the functional activity of the synthesized growth factors, we selected FGF-1, IL-3, and IL-15 for cell-based proliferation assays, due to their clinical relevance in treating cardiovascular disorders(*52*), bone marrow failure(*58*), and cancer(*53*), respectively. CFPS expression and centrifugal purification yielded proteins with purity above 90% and endotoxin levels meeting standard guidelines (≤ 0.1 EU/mL) **(Fig. S7)**(*59*). The bioactivity of FGF-1, IL-3, and IL-15 growth factors was tested using NIH-3T3 mouse embryonic fibroblast cell line, TF-1 human erythroleukemia cell line, and primary human CD3+ T cells, respectively. In-house FGF-1, IL-3, and IL-15 enhanced proliferation with approximate log_10_(EC_50_) values of 2.7 ng/mL, -0.6 ng/mL, and 2.4 μg/mL, respectively. Titrations also revealed that in-house growth factors stimulated proliferation comparable to their commercial counterparts, except for IL-15, which retained partial functional activity (**Fig. 3c,d**, **Fig. S7).**

With functional activity confirmed for in-house FGF-1, we next sought to determine the feasibility of on-site growth factor biomanufacturing in resource-constrained settings. While working in Brazil, we successfully produced FGF-1 using FD- CFPS reactions and, similarly to experiments conducted in Canada, achieved high purity (> 90 %) **(Fig. S8)** and low endotoxin levels (≤ 0.1 EU/mL) **(Fig. S8)**(*59*). Functional validation using NIH-3T3 mouse embryonic fibroblasts demonstrated that synthesized FGF-1 exhibited comparable potency in cell proliferation assays (**Fig. 3e**), confirming the capacity of CFPS for distributed growth factor production.

### Cell-free produced Nuvax elicits a robust IgG-specific immune response in mice

Vaccines are among the most effective tools for reducing disease burden(*60*). However, vaccine production is centralized and resource-intensive, precluding rapid response and limiting access in low-resource settings(*61*). Building on our previous demonstration of vaccine synthesis using high-cost recombinant CFPS(*25*) and other studies on conjugate vaccines from bacterial lysates(*26, 43*), we sought to establish a low-cost, lysate-based platform for decentralized vaccine production. Here, we focus on a promising SARS-CoV-2 N-protein-based vaccine for broad-spectrum protection against COVID-19(*62–65*), with vaccine immunogenicity assessed using a murine model (**Fig. 4a**).

**Fig. 4:**
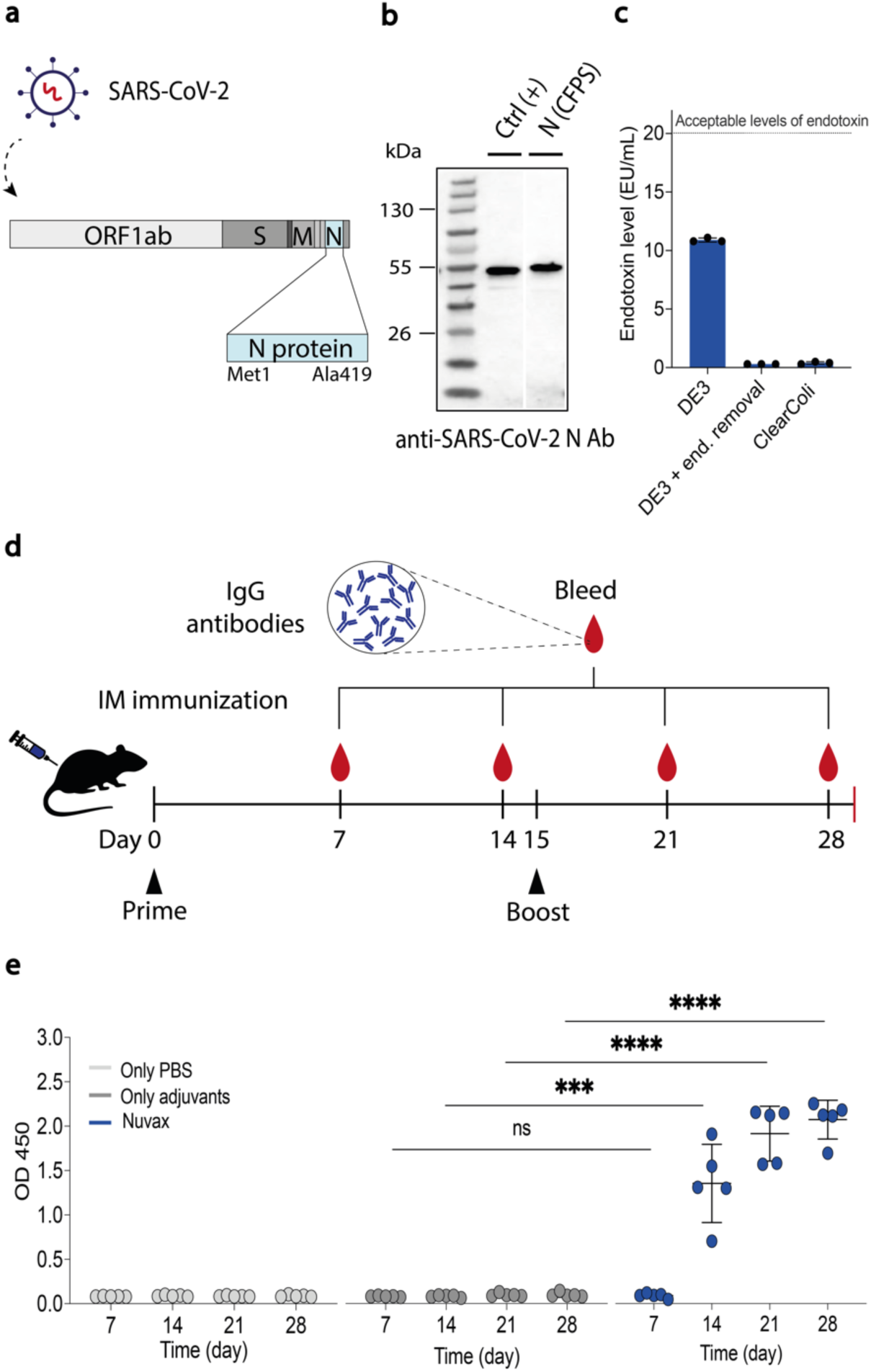
On-site cell-free production of Nuvax COVID-19 vaccine and induction of IgG response in mice. **(a)** Schematic of the SARS-CoV-2 genome; the coding sequence for the SARS-CoV-2 nucleocapsid protein was optimized for translation in CFPS and ultimately used for Nuvax formulation. **(b)** Western blot analysis of CFPS-produced SARS-CoV-2 nucleocapsid protein exhibiting a distinct band at the expected molecular weight (49 kDa). The SARS-CoV-2 nucleocapsid protein from SinoBiologicals, a commercially available recombinant protein, was used as a positive control (Ctrl +). N (CFPS) refers to the antigen expressed using in-house CFPS reactions. The molecular weight ladder (in kilodaltons) is shown on the left. Data shown are from one representative biological replicate out of three independent experiments. **(c)** Endotoxin levels in CFPS-produced SARS-CoV-2 nucleocapsid protein. Three different strategies were used for antigen expression: 1) cell-free lysates made from *E. coli* BL21(DE3); 2) cell-free lysates from *E. coli* BL21(DE3) followed by endotoxin removal; 3) cell-free lysates prepared from ClearColi^TM^ BL21(DE3), an engineered *E. coli* strain used to express endotoxin-free recombinant proteins, making it ideal for expression of therapeutic products. The dashed line indicates the standard guidelines for subunit-based vaccine formulations (<20 EU/mL). Data are shown as mean ± SD, n = 3. **(d)** Immunization schedule, indicating prime, boost and bleed timepoints, for the CFPS-derived Nuvax. Intramuscular (IM) vaccination was administered on days 0 (prime) and 15 (boost), and blood was drawn on days 7, 14, 21, and 28 to evaluate for induction of IgG production. **(e)** Robust neutralizing antibody response following Nuvax administration alongside group controls. Analysis was performed using ELISA and measured at OD 450 nm using a conventional plate reader. The data are presented as the mean ± SD for each group, with individual data points shown (control group that received only PBS, n = 5; control group that received only adjuvants, n = 5; Nuvax group, n = 5). Statistical differences were determined by two-way ANOVA with Tukey’s post hoc multiple comparisons test: ns p > 0.05, * p < 0.05, ** p < 0.01, *** p < 0.001, **** p < 0.0001. Abbreviations are: ns, not significantly different; Ctrl, control; N, nucleocapsid; End, endotoxin; Ab, antibody; IM, intramuscular; CFPS, cell-free protein synthesis.

We began by expressing the nucleocapsid antigen in two in-house CFPS reactions based on BL21(DE3) and SHuffle *E. coli* strains, and two commercial CFPS kits (10 μL, 24 °C, 16 hours). Western blot analysis (anti-His_6_-tag and anti-SARS-CoV-2 nucleocapsid) confirmed nucleocapsid expression at the expected molecular weight (49 kDa) **(Fig. S9a,b)** and the presence of the correct epitope, as determined by an anti-nucleocapsid antibody (**Fig. 4b**).

To obtain a high-purity, low-endotoxin product suitable for therapeutic use, we combined two-step spin column purification (Ni-NTA (His_6_-tag) and Strep-Tactin^TM^ (Strep II tag)) with endotoxin removal for BL21(DE3)-derived products using poly(ε-lysine) treatment, and also generated cell lysates from ClearColi^TM^ BL21(DE3), an engineered strain designed to not elicit the endotoxin response(*66*). Both approaches yielded a soluble nucleocapsid antigen with endotoxin levels below the accepted limits for subunit-based vaccine formulations (<20 EU/mL) (**Fig. 4c**)(*67*). From a 1 mL in-house

ClearColi^TM^ CFPS reaction, we produced antigen sufficient for four individual human doses (assuming a dose of 5 μg)(*68*) with greater than 90% purity **(Fig. S9c).**

With vaccine production protocols in place, we assessed the immunogenicity of the vaccine antigen in a murine model. Starting with a 1 mL ClearColi^TM^ BL21(DE3)- derived CFPS reaction (1 mL, 24 °C, 16 hours), 23.2 μg of soluble protein was produced using the two-step purification. The nucleocapsid antigen was then combined with adjuvants (pattern recognition receptor agonist CpG-2395 and aluminum salts [alhydrogel]) to enhance immune response(*69*), producing a novel COVID-19 vaccine formulation we designated as Nuvax.

To evaluate the induction of an anti-SARS-CoV-2 nucleocapsid IgG antibody (Ab) response, we conducted vaccination trials with BALB/c mice and Nuvax at the World Reference Center for Emerging Viruses and Arboviruses in Galveston, Texas, USA. A mouse test group received doses of Nuvax containing 1 μg/animal on days 0 (prime) and 15 (boost), while control groups were vaccinated with phosphate-buffered saline (PBS; mock) or adjuvant only (**Fig. 4d**). Blood was collected on days 7, 14, 21, and 28 for analysis of IgG production. Strong induction of anti-SARS-CoV-2 nucleocapsid IgG Ab production in Nuvax-vaccinated mice was confirmed two weeks after the prime dose, while no Ab induction was observed in either of the control groups (**Fig. 4e**).

### Developing low-cost synthetic biosensors in response to public health crises

Access to diagnostics responsive to local health priorities is crucial for health and economic resilience(*70*), yet half of the world’s population lacks meaningful access to diagnostics(*71*). This underscores the importance of local capacity for diagnostics development that can be directed to local needs, with production costs calibrated to domestic economics and needs, such as tropical and mosquito-borne infections.

To address this need and building on previous work demonstrating low-burden molecular diagnostics(*29, 39, 72, 73*), we developed and validated toehold switch-based sensors for SARS-CoV-2 detection (**Fig. 5a**). Using an open-source computational design algorithm **(see Extended Methods)**, we generated 142 toehold sensors and screened them based on ON/OFF activation ratios measured by β-galactosidase (*lacZ*) activity (**Fig. 5b**). With a top performing sensor identified (D07), we then focused on detecting target nucleic acids at clinically relevant concentrations. Here, an isothermal nucleic acid amplification method, RT-LAMP (reverse transcriptase loop-mediated isothermal amplification), was placed upstream of the toehold switch as a pre-amplification step in the diagnostic workflow(*74, 75*). When paired with RT-LAMP amplicons in PURExpress cell-free protein expression reactions, the D07 switch demonstrated robust diagnostic performance, achieving diagnostic accuracy of 90.91% (95% CI 58.72% to 99.77%) in a patient trial using samples with Ct value ≤30 (**Fig. 5c**, **Tables S3,4).**

**Fig. 5:**
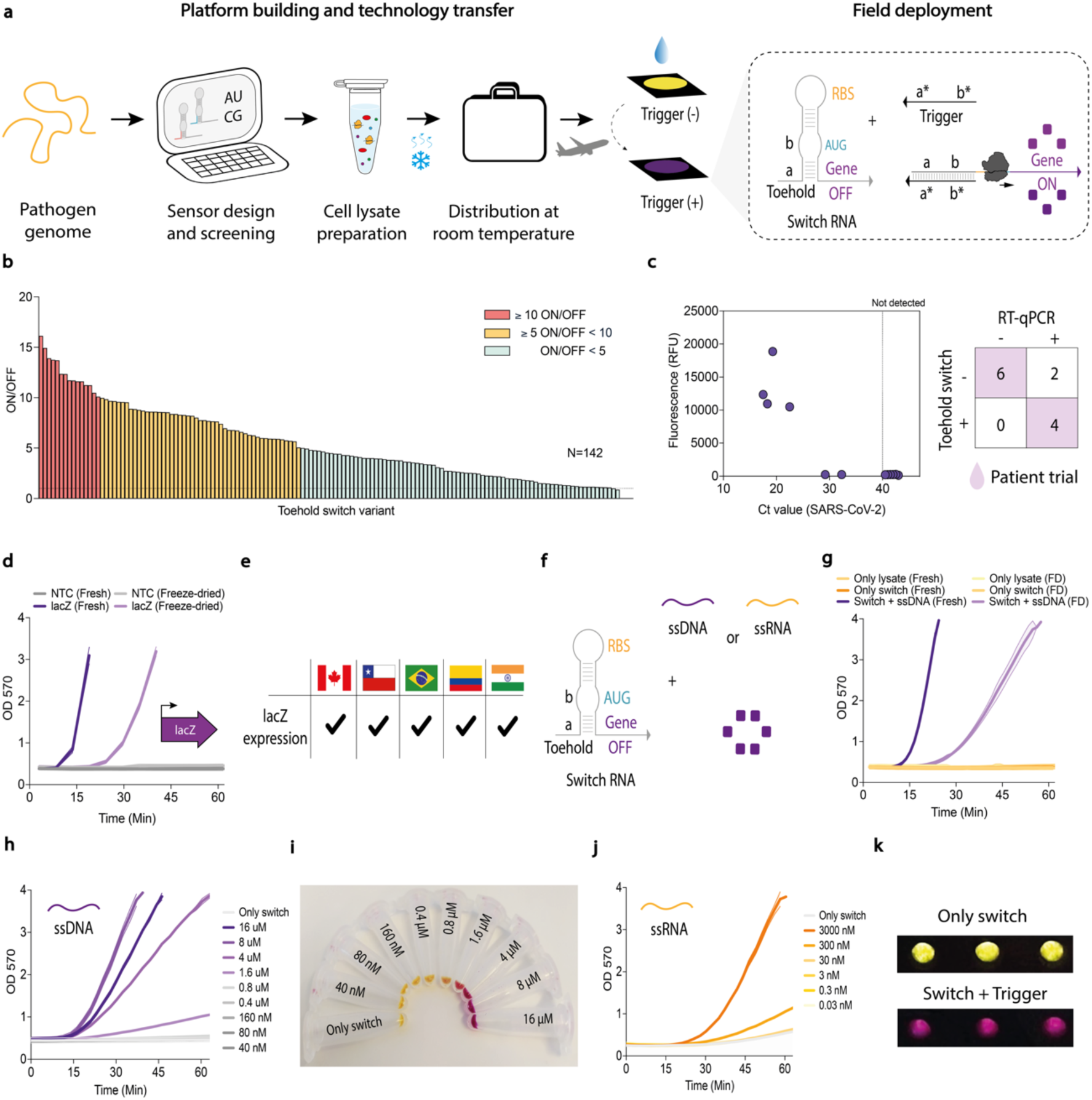
Development, global distribution, and validation of toehold switch diagnostics. **(a)** Schematic representation of the toehold switch-based diagnostic workflow, from computational design to experimental validation in diverse locations. **(b)** Screening of 142 toehold switch variants that translationally regulate the expression of the *lacZ* gene. D07 was identified as the top-performing sensor based on ON/OFF activation ratios in the presence of synthetic trigger nucleic acid. **(c)** The D07 toehold switch demonstrated robust SARS-CoV-2 diagnostic performance in PURExpress when activated with RT-LAMP nucleic acids from nasopharyngeal swabs. Ct value of the patient samples as measured independently by RT-qPCR (nucleocapsid gene), plotted against the resulting fluorescence (state molecule), a proxy of toehold-switch activation. **(d)** *lacZ* expression constructs, transcriptionally and translationally driven by a T7 promoter and a canonical Shine-Dalgarno sequence, respectively, were distributed to multiple international sites. Lysate performance, measured by β-galactosidase activity, was assessed by measuring absorbance at 570 nm over 1 h. This representative data was obtained using cell-free lysates produced on-site in Chile. Data are shown as mean ± SD, n = 3. **(e)** Reproducibility assessment for β-galactosidase expression performed across multiple international sites (Canada, Chile, Brazil, Colombia, and India). **(f)** Schematic of a toehold switch and the different nucleic acid triggers (ssDNA and ssRNA) used in these assays. **(g)** Activation of the D07 toehold switch in fresh vs. freeze-dried lysates. ssDNA trigger was added to D07 toehold switch-containing lysate reactions, and β-galactosidase activity was measured by absorbance at 570 nm for 1 h. Successful activation was observed in Chile, Canada, and Colombia, while inconsistent results were noted in other locations due to shipping-related lysate degradation. Representative data was obtained using cell-free lysates produced on-site in Chile. Data are shown as mean ± SD, n = 3. **(h)** Titration of up to 16 µM synthetic ssDNA trigger into D07 toehold switch-containing lysate reactions. β-galactosidase activity was assessed by measuring absorbance at 570 nm over 1 h. This representative data was obtained using cell-free lysates produced on-site in Chile. Data are shown as mean ± SD, n = 2. **(i)** Photograph confirms the activation of the D07 toehold switch and subsequent enzyme activity. **(j)** Titration of up to 3000 nM *in vitro* transcribed synthetic ssRNA trigger into D07 toehold switch-containing cell-free lysate reactions. β-galactosidase activity was assessed by measuring absorbance at 570 nm over 1 h. This representative data was obtained using cell-free lysates produced on-site in Chile. Data are shown as mean ± SD, n = 2. (k) Photograph confirmed the activation of the D07 toehold switch by synthetic RNA and subsequent enzyme activity. A positive reaction resulted in a color change from yellow to purple, while a negative reaction remained yellow. Abbreviations are: NTC, non-template control; FD, freeze-dried; RBS, ribosome binding site; Min, minutes.

With the diagnostic system validated (D07 toehold switch + RT-LAMP), we sought to advance the test toward a format suitable for production in diverse geographic and resource-limited settings. Toehold switch-based diagnostics typically rely on commercial recombinant CFPS (PURExpress, NEB), which is costly ($7.80 USD per test compared to $0.069 USD for in-house CFPS reactions)(*76*) and requires cold-chain distribution in liquid form(*76*). To adapt the toehold sensors to low-cost *E. coli* lysate-based CFPS, we optimized test parameters and evaluated the effect of dialysis on lysate preparation **(Fig. S10)**(*76*). We also integrated Tus-Ter protection^69^ into linear DNA constructs encoding the toehold sensors, which improved their stability and activity by reducing the impact of endogenous exonucleases present in lysate-based CFPS reactions **(Fig. S11).**

To evaluate assay reproducibility in different laboratory environments, constitutive *lacZ* expression constructs were tested at five international sites, validating the *E. coli* lysate-based CFPS performance and the feasibility of running the reporter assay in different settings (**Fig. 5 d,e**, **Fig. S12).** This test was then repeated for the D07 toehold switch by distributing DNA constructs and FD-CFPS (prepared in Santiago, Chile) to multiple sites (**Fig. 5f,g**). While successful in Canada, Chile, and Colombia, test performance was inconsistent in other locations **(Fig. S13)**, where shipping and customs delays impacted CFPS activity, highlighting the everyday challenges faced by LMIC researchers.

Team members in Canada and Chile validated protocols for cross-site standardization by measuring D07 sensor activation using a synthetic ssDNA target, which can be shipped as a stable control (**Fig. 5h-i**, **Fig. S14)**, alongside conventional RNA target (**Fig. 5j**) and a demonstration of colorimetric readout (**Fig. 5k**). We found that such stable controls and defined standard operating procedures are crucial for ensuring reproducibility across collaborative networks and for supporting the future development of decentralized diagnostic programs. Additionally, the challenges associated with shipping highlighted the need for local manufacturing of essential diagnostic components.

### Building an efficient diagnostic testing pipeline combining locally manufactured LAMP/RT-LAMP reactions with open-source hardware

In response to these logistical challenges, we focused on developing simple and robust molecular diagnostics based on LAMP(*77*). This method is a powerful standalone tool for nucleic acid detection with limited laboratory infrastructure (no thermal cycler), with at least 12 approved RT-LAMP-based tests for SARS-CoV-2 under the United States FDA Emergency Use Authorization (EUA)(*13, 77–81*). Here, we demonstrate a pipeline (comprising steps I-IX) for the on-site production of LAMP enzymes (<24 hours) and the development of diagnostic tests for 16 clinically relevant pathogens **(Fig. S15)**. This includes the development of two new molecular tests targeting the emerging mosquito-borne Oropouche virus in Latin America(*82*) and highly pathogenic avian influenza A (H5N1) virus(*83*), in response to these ongoing global health threats.

At the center of this pipeline was the on-site manufacture of diagnostic reagents demonstrated in resource-limited settings and test validation through patient trials for three endemic infections across four countries (116 samples). DNA detection with LAMP requires only one enzyme, Bst DNA Polymerase Large Fragment (Bst LF)(*77*), which can be extended to RNA targets (RT-LAMP) with the addition of a reverse transcriptase, such as the Moloney Murine Leukemia Virus Reverse Transcriptase (M-MLV) used here(*78*). Using fresh CFPS reactions (500 μL, overnight at ambient temperature), we successfully expressed and purified functional Bst LF (191.5 μg) and M-MLV (120.4 μg), sufficient for over 3,500 and 6,000 reactions, respectively **(Fig. S16, Supplementary Note 3)**.

Having established low-burden enzyme production (step I), we next set out to create diagnostic tests (step II) for 16 pathogens: *Borrelia burgdorferi, Mycobacterium tuberculosis, Plasmodium falciparum, Leishmania donovani, Leishmania braziliensis,* human immunodeficiency virus 1 (HIV-1), chikungunya virus (CHIKV), monkeypox virus (MPXV), Zika virus (ZIKV), dengue virus serotype 2 (DENV-2), West Nile virus (WNV), Mayaro virus (MAYV), SARS-CoV-2, Powassan virus (POWV), avian influenza A (H5N1) virus, and Oropouche virus (OROV) (**Fig. 6a**, **Table S5)**. Considering the importance of optimization in diagnostic development(*13, 78*), we began by refining LAMP and RT-LAMP reaction conditions (step III), including reagent and supplement concentrations (e.g., guanidine hydrochloride [GuHCl]) to improve reaction performance **(Fig. S17)**(*84*). Using these optimized conditions, we benchmarked the activity and fidelity of our in-house LAMP/RT-LAMP reactions (blue data) against a commercial kit (green data), finding equivalent performance for DNA and RNA targets across all 16 pathogen sequences with signal detection in as little as 10-20 minutes (step IV) (**Fig. 6b**, **Fig. S18, see Methods for details)**. To demonstrate that molecular diagnostic tests can also be transported without cold chain logistics (step V), we freeze-dried our in-house produced RT-LAMP reactions with results confirming that they could be distributed into a field-stable format **(Fig. S19)**.

**Fig. 6:**
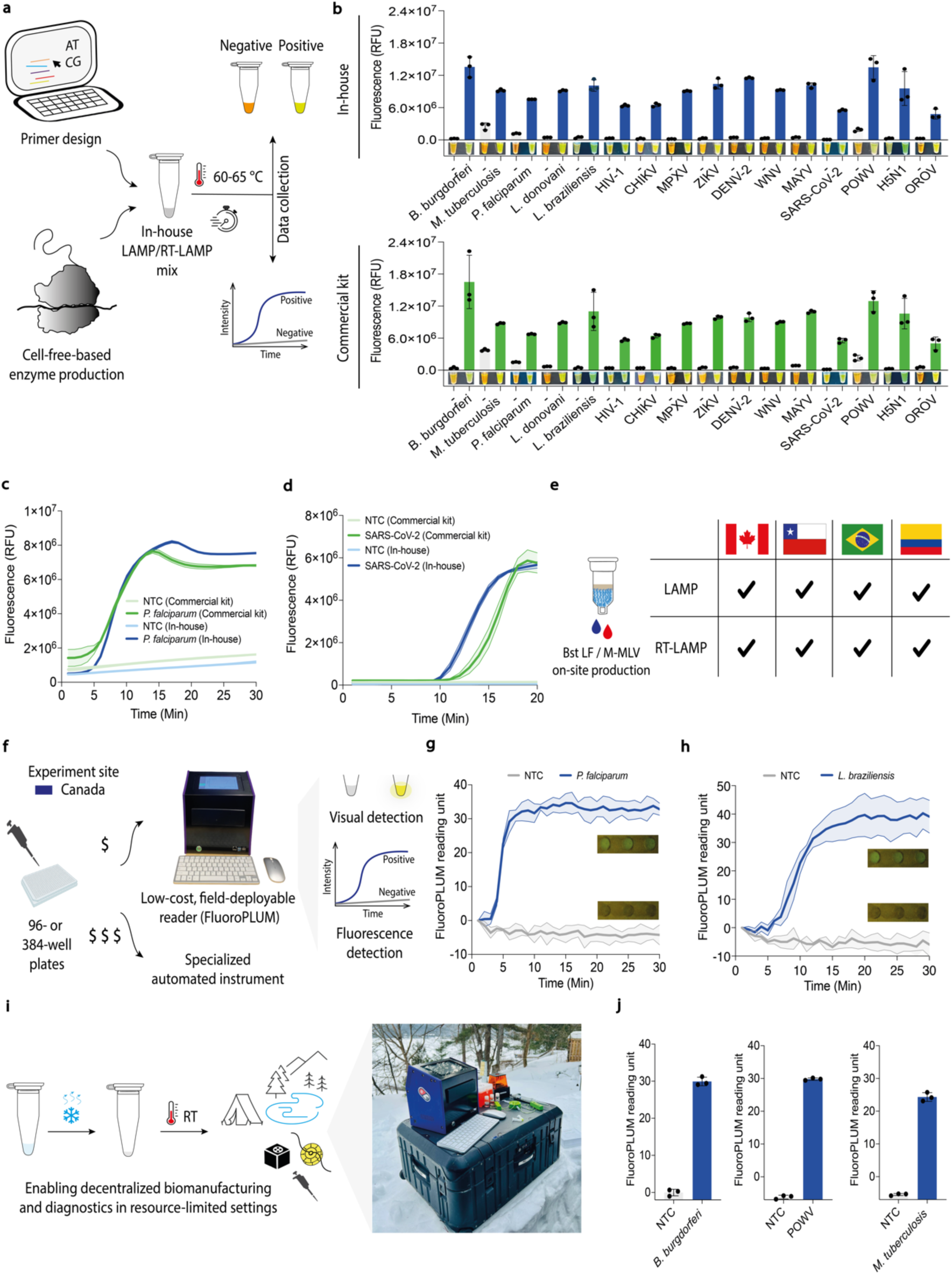
Local cell-free biomanufacturing and open-source hardware enhance diagnostic capacity and access in resource-limited settings, matching commercial gold standards. **(a)** Schematic representation of the pipeline used to create the molecular diagnostic systems. Detection was carried out using real-time fluorescence or visual detection. For fluorescence measurements, amplicons were visualized by adding 1x LAMP fluorescent dye or 10 μM SYTO 9 Green Fluorescent Nucleic Acid dye if FluoroPLUM was used, with fluorescence reads every minute. For visual detection, the naked eye confirmed amplification under natural light by adding SYBR Gold Nucleic Acid Stain (diluted 1:10) to tube caps before reaction incubation, followed by mixing after incubation. A positive reaction resulted in a color change from orange to yellow, while a negative reaction remained orange. **(b)** End point fluorescence levels (25 minutes for all targets) for 16 pathogens, comparing in-house (blue) and commercial LAMP systems (green). Data are shown as mean ± SD, n = 3. **(c)** Real-time fluorescent experiments for synthetic *P. falciparum* DNA amplification using a conventional qPCR instrument. This representative data was obtained using reagents produced on-site in Canada. Data are shown as mean ± SD, n = 3. **(d)** Real-time fluorescent experiments for synthetic SARS-CoV-2 RNA amplification using a conventional qPCR instrument. This representative data was obtained using reagents produced on-site in Canada. Data are shown as mean ± SD, n = 3. **(e)** Reproducibility assessment of LAMP and RT-LAMP performed in Canada, Chile, Brazil, and Colombia. Independent experiments conducted by different teams across diverse settings, using a standardized protocol, confirmed the reproducibility of the results. The consistent results reinforce the feasibility of decentralized enzyme production. **(f)** Comparison of the low-cost, portable FluoroPLUM and a conventional high-cost qPCR machine for assessing in-house-produced LAMP reactions, with results showing comparable performance across both platforms, confirming the utility of FluoroPLUM as an affordable alternative for high-throughput measurements in low-resource settings. **(g)** Real-time fluorescence and visual outputs of LAMP reactions using *P. falciparum* synthetic DNA (2 nM) and NTC measured with the low-cost, portable FluoroPLUM. This representative data was obtained using reagents produced on-site in Canada. Data are shown as mean ± SD, n = 3. **(h)** Real-time fluorescence and visual outputs of LAMP reactions using *L. braziliensis* synthetic DNA (2 nM) and NTC measured with the low- cost, portable FluoroPLUM. This representative data was obtained using reagents produced on-site in Canada. Data are shown as mean ± SD, n = 3. **(i)** Deploying low-cost cell-free lysates and open-source hardware enables decentralized biomanufacturing, facilitating the implementation of diagnostic programs in resource-limited settings. **(j)** End point fluorescence levels (30 minutes for all targets) for *B. burgdorferi*, POWV, and *M. tuberculosis* measured using the low-cost, portable FluoroPLUM. This representative data was obtained using diagnostic reagents manufactured on-site in Algonquin Highlands, Ontario, Canada. Data are shown as mean ± SD, n = 3. Abbreviations are: (-) or NTC, non-template control; RT, room temperature; Min, minutes; HIV-1, human immunodeficiency virus 1; CHIKV, chikungunya virus; MPXV, monkeypox virus; ZIKV, Zika virus; DENV-2, dengue virus serotype 2; WNV, West Nile virus; MAYV, Mayaro virus; POWV, Powassan virus; OROV, Oropouche virus.

With in-house enzyme activity validated, we advanced the project toward on-site enzyme production using three different centrifugal devices for protein spin column purification. Side-by-side comparisons of a benchtop laboratory centrifuge, a battery-powered centrifuge, and our 3D-fuge found comparable yield, purity, and functional activity **(Fig. S20)**. We then expanded these efforts into a multi-site reproducibility assessment (step VI), shipping FD-CFPS reaction mixtures (prepared in Toronto, Canada) to partner laboratories in Chile, Brazil, and Colombia for on-site production of Bst LF and M-MLV **(Fig. S21)**. For this phase of the project, we used *P. falciparum* (as a DNA target) and SARS-CoV-2 (as an RNA target) as examples of globally relevant pathogens, with analytical parameters confirming high sensitivity at clinically relevant concentrations (2 fM or 5 copies/μL) and specificity for both targets **(Fig. S22)**(*85*). Using synthetic nucleic acid templates, in-house systems (LAMP and RT-LAMP modalities) with on-site manufactured reagents (enzymes and buffers) were benchmarked against a commercially available kit at each test site. Fluorescence (**Fig. 6c-e**, **Fig. S23a-c),** colorimetric, and electrophoresis readouts confirmed comparable performance and fidelity to commercial kits **(Fig. S23d,e)**. Here, inter-assay variability consistently remained low across both in-house and commercial systems, with average CVs of 4.77% for the in-house system and 5.99% for the commercial kit across all participating laboratories (n = 4), confirming high reproducibility(*86*).

To enable distributed high-capacity testing, we previously developed PLUM(*29*), a portable, low-cost, open-source plate reader. Here, we introduce the next-generation optical reader, FluoroPLUM, which enables the fluorescence monitoring of LAMP reactions in 96- or 384-well plate formats (step VII) **(see Methods for details)**(*87*). This device provides incubation at 65 °C and quantitative monitoring of assay progression. In-house RT-LAMP reactions were run alongside a conventional qPCR instrument for direct comparison (**Fig. 6f**), with results showing comparable performance across both platforms **(Fig. S24)**. To further validate FluoroPLUM’s functionality, additional laboratory tests were performed to detect human parasites (*P. falciparum* and *L. braziliensis*). The tests were performed on both synthetic (**Fig. 6g,h**) and cultured- pathogen templates at two locations **(Fig. S25)**, and again delivered results comparable to those of gold-standard techniques (qPCR and microscopy) **(Fig. S25)**.

The growing demand for molecular diagnostics in austere settings highlights a critical need, whether for environmental science or human health(*88*). Having established the potential for local production in laboratory settings, we next tested the concept of distributed biomanufacturing at two temporary research sites to simulate remote work (step VIII) (Algonquin Highlands, ON, and Whitehorse, YT) in Canada (**Fig. 6i**, **Fig. S26, Supplementary Video).** On-site work successfully expressed and purified Bst LF and M-MLV using the battery-powered and 3D-printed hand-powered centrifuges **(Fig. S26)**. Once produced, the enzymes were used in combination with our novel diagnostic tests to diagnose tick-borne pathogens (*B. burgdorferi,* the causative agent of Lyme disease, and POWV, the causative agent of Powassan virus encephalitis), which are becoming increasingly prevalent in parts of Canada due to climate change(*89, 90*), and *M. tuberculosis*, which remains a widespread public health concern in First Nations communities across Canada, particularly in Canada’s North (**Fig. 6j**, **Fig. S26)**(*91*).

### Multi-center and international patient trial assessment of locally manufactured diagnostic reagents

With our diagnostic testing pipeline implemented across diverse geographic and resource-limited settings, we expanded our efforts to establish disease diagnostic programs in regions of endemic infection (step IX). We selected a representative subset of clinically relevant pathogens for subsequent field-based clinical testing. This included SARS-CoV-2, owing to its ongoing global impact(*92*), and CHIKV and OROV, which are of particular concern in tropical and sub-tropical regions(*18, 82*). Using locally manufactured diagnostic reagents, we rigorously validated our in-house diagnostic systems through patient trials in Canada, Chile, Brazil, and Colombia, with results directly compared to those obtained in parallel using US CDC gold-standard RT-qPCR assays (**Fig. 7a**).

**Fig. 7:**
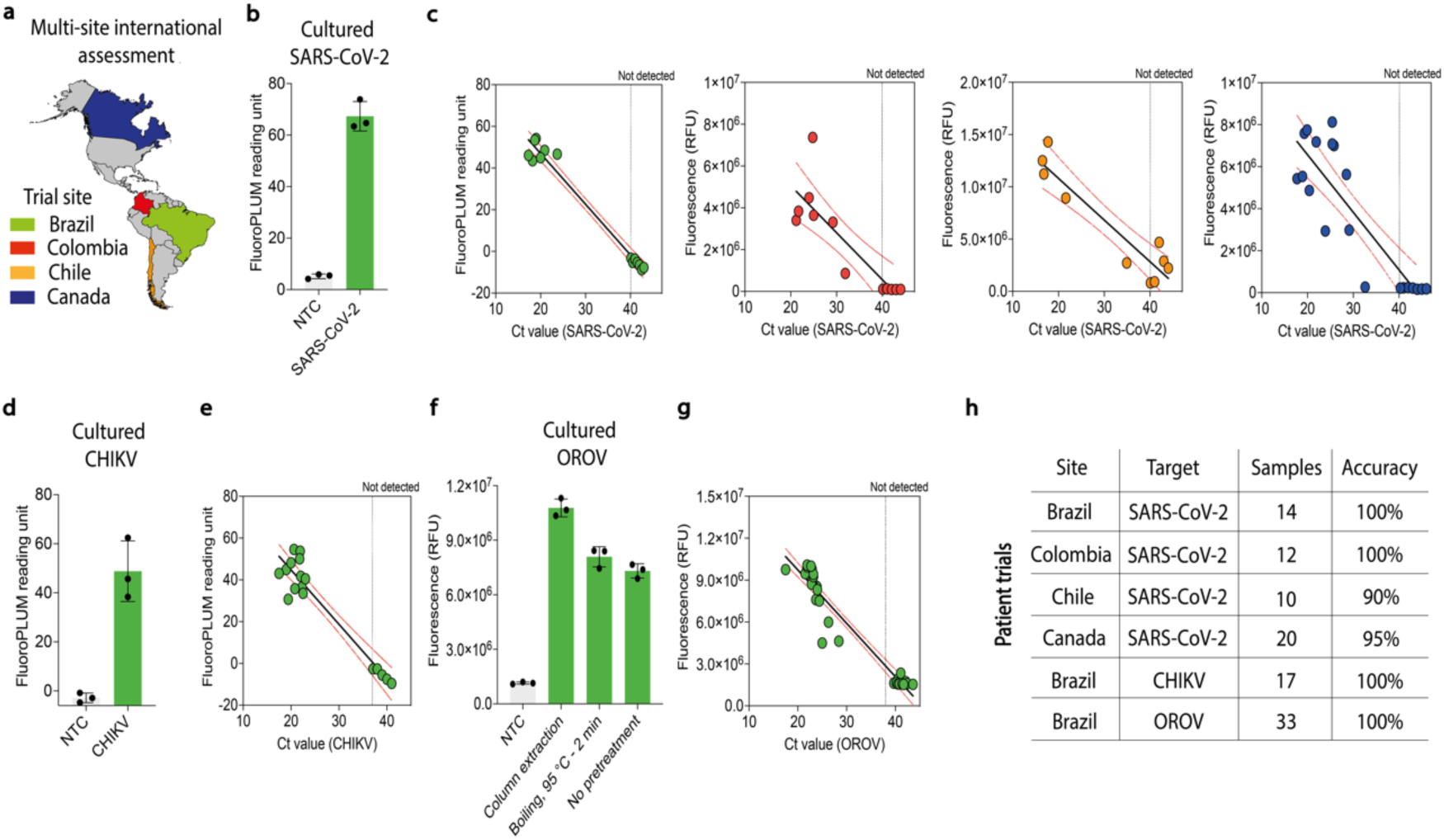
A scalable, affordable platform brings decentralized disease detection in regions of endemic infection. **(a)** Countries where diagnostics were manufactured on-site and successfully used to establish disease diagnostic programs. Colors correspond to the data shown in subsequent panels, representing the countries where the data was collected. **(b)** The functionality of on-site-produced RT-LAMP reactions was further tested with RNA isolated from cultured SARS-CoV-2 virus. Fluorescence measurements after 20 minutes of incubation were plotted. Data are shown as mean ± SD, n = 3. **(c)** Patient trials for a locally produced SARS-CoV-2 diagnostic program were conducted in four different countries: Brazil, Colombia, Chile, and Canada. RNA samples isolated from patients were analyzed via RT-LAMP, where fluorescence increases indicate successful amplification. Fluorescence after 20 minutes (y-axis) was plotted against corresponding Ct values obtained via CDC RT-qPCR gold-standard assays (x-axis). In-house diagnostic tests demonstrated diagnostic accuracy ranging from 90% to 100%. The dashed line represents the threshold value defined for RT-qPCR analysis. See Supplementary Information for detailed analysis. **(d)** The functionality of on-site-produced RT-LAMP reactions was further tested with RNA isolated from cultured chikungunya virus. Fluorescence measurements after 30 minutes of incubation were plotted. Data are shown as mean ± SD, n = 3. **(e)** Patient trial for chikungunya virus was conducted in Brazil using locally produced, on-demand diagnostics. RNA samples isolated from patient samples were analyzed via RT-LAMP using 10 μM SYTO 9 Green fluorescent nucleic acid dye, where fluorescence increases indicated successful amplification. Fluorescence after 30 minutes (y-axis) was plotted against the corresponding Ct values obtained using the CDC RT-qPCR gold-standard assays (x-axis) in parallel. The dashed line represents the threshold value defined for RT-qPCR analysis**. (f)** Cultured Oropouche virus was processed using three methods: (1) commercial column extraction, (2) simple boiling at 95 °C for 2 minutes, and (3) direct use of the cultured virus without pretreatment. Each sample was tested using on-site-produced RT-LAMP reactions with LAMP fluorescent dye to detect the virus, with all three methods yielding successful detection. Data are shown as mean ± SD, n = 3. **(g)** Patient trial for Oropouche virus was conducted in Brazil using locally produced, on-demand diagnostics. RNA samples isolated from patient samples were analyzed via RT-LAMP using 1X LAMP fluorescent dye, where fluorescence increases indicated successful amplification. Fluorescence after 40 minutes (y-axis) was plotted against the corresponding Ct values obtained using the RT-qPCR gold-standard assays (x-axis). The dashed line represents the threshold value defined for RT-qPCR analysis. **(h)** In parallel with the CDC RT-qPCR gold standard, in-house diagnostic tests for SARS-CoV-2, chikungunya, and Oropouche viruses demonstrated accuracy ranging from 90% to 100%. During this project phase, RNA quality and integrity were verified in all patient samples using the human endogenous controls for Ribonuclease P (RT-qPCR) and Beta-actin (RT-LAMP). See Supplementary Information for detailed analysis (Tables S6-12). Abbreviations are: NTC, non-template control; CHIKV, chikungunya virus; OROV, Oropouche virus; Ct, cycle threshold.

We first validated our in-house RT-LAMP system using cultured SARS-CoV-2 (**Fig. 7b**) in preparation for pathogen detection from extracted COVID-19 patient samples. With enzyme inputs produced at each site, in-house RT-LAMP reactions were tested in a double-blinded format on a total of 66 specimens in four countries (Canada, Chile, Brazil, and Colombia), spanning the Global North and South (**Fig. 7c,h**, **Fig. S27)**. Compared to the CDC RT-qPCR assay(*93*), we achieved diagnostic accuracies between 90 and 100%, with an average performance of 96.2% **(Tables S6-10 for detailed analysis).**

To demonstrate the extensibility of the approach, we then expanded these efforts to diagnose CHIKV, another arbovirus with worldwide distribution and clinical importance, which can lead to severe and potentially fatal disease(*18*). As before, we began by confirming the ability of our in-house RT-LAMP system to detect cultured CHIKV (**Fig. 7d**). Working in a national reference laboratory for arbovirus diagnostics in Brazil, the epicenter of chikungunya epidemics in the Americas with over 1.6 million reported cases to date(*18*), we conducted a patient trial. In a double-blinded study design, a total of 17 extracted patient samples were tested using our in-house RT-LAMP system in parallel to the US CDC RT-qPCR protocol as a gold-standard comparison (**Fig. 7e,h**, **Fig. S28)**(*94*), which translated to a diagnostic accuracy of 100% (95% CI 80.49% to 100%) **(Table S11)**.

With the growing threat of the mosquito-borne OROV in Latin America(*17, 82*), we sought to highlight the potential for the real-time development of low-burden Oropouche testing and on-site biomanufacturing to respond to urgent public health needs. Using cultured OROV **(Fig. S29a)**, we first confirmed its detection following a simple boiling step for viral lysis (e.g., 95 °C for 2 minutes) as well as directly without sample pretreatment (**Fig. 7f**, **Fig. S29b)**.

To further validate the utility of this diagnostic assay in clinical settings, we performed RT-LAMP directly on 33 serum samples collected from suspected cases of arboviral infection in Brazil, the epicenter of the ongoing Oropouche outbreak(*82*), and then compared the results to the gold-standard RT-qPCR assay in a double-blinded format (**Fig. 7g,h**, **Fig. S29c,d)**(*95*). Compared to RT-qPCR, we found 100% (95% CI 89.42% to 100%) diagnostic accuracy **(Table S12).** These findings highlight the reliability of locally produced diagnostic reagents and their potential to strengthen disease diagnostic programs in diverse settings, from clinics to remote areas, effectively addressing healthcare needs locally and in underserved communities.

## DISCUSSION

Here, with researchers from both well-resourced and resource-limited settings, we collaboratively designed scalable and adaptable biomanufacturing solutions to address pressing global challenges. This study lays the foundation for fundamental shifts in biotechnology manufacturing practices in LMICs and developing nations, moving from reliance on centralized and outsourced production facilities to adopting decentralized, local production platforms. To support this transformation, we designed, validated, and implemented novel systems in research teams across North America, South America, and Asia. The approach incorporated invaluable firsthand insight into how conventional reagent procurement can bottleneck scientific progress in resource-limited settings and, importantly, positioned technology development to effectively pilot and refine innovation. Having now applied local biomanufacturing to augment research applications in resource-limited settings, we see this approach as having the potential to significantly accelerate locally driven fundamental life sciences research, healthcare, and, more broadly, the global bioeconomy.

We began with the development of a research network for open-source technology transfer and capacity-building, fostering collaboration with research partners on three continents and supporting on-the-ground implementation and standardization (**Figs. 1,2**). Through this initiative, we leveraged two key technologies to enable local biomanufacturing. This included low-burden CFPS reactions that can be distributed and stored at ambient temperature, and affordable, portable, user-friendly, open-source hardware. With efficient, low-cost systems in place, rapid on-site production of high value bioproducts, including growth factors, vaccines, and diagnostic enzymes, became achievable within a single day and at a fraction of the typical cost **(Appendix 1)**. This is something that has typically not been possible outside of bioproduction laboratories. Using molecular, cell-based, animal model, and clinical sample testing, these bioproducts were rigorously validated in a series of proof-of-concept experiments and multi-site clinical trials. Direct comparisons with high-cost commercial reagents, the current gold standards, demonstrated similar performance, efficiency, precision, and reproducibility (**Figs. 3-7**).

Economic feasibility and accessible raw materials and hardware were central factors guiding technology development, ensuring that solutions could be realistically implemented within LMIC communities. As such, a primary goal of this project was to develop a platform for the affordable biofabrication of diagnostics and therapeutics at the point-of-use. Our estimated cost per 1 mL CFPS reaction is ∼$5.5 USD, which is comparable to other published in-house *E. coli*-based CFPS systems(*43*) and approximately 70 times less expensive than *E. coli*-based CFPS commercial systems **(Appendix 1).** While distributed cell-based expression could also technically achieve the related production, it comes with a requirement for significant on-site infrastructure. The cell-free approach allows for local equipped laboratories or ventures to make CFPS and supply the research community. In many ways, this approach reflects the early days of molecular biology where laboratories made their own reagents, but, with the advances in cell-free production, the technical bar is within reach of most laboratories.

These factors position cell-free local biomanufacturing to serve as a compelling approach for affordable local production of bioreagents in both low- and high-resource settings. This is exemplified in our RT-LAMP clinical diagnostics demonstrations, where diagnostic enzymes for 3,500-6,000 reactions were produced within one day (from 0.5 mL CFPS reactions, ∼$2.75 cost of goods), which translates to just $0.27 USD per in-house molecular test, a stark contrast to the $1.40 USD per RT-LAMP reaction using commercial kits and $8 USD per RT-qPCR gold-standard sample test **(Appendix 1)**. While capital intensive, it is important to acknowledge that the cost of commercial products also includes the costs of R&D, marketing, distribution, and profit margins(*96*). Local cell-free biomanufacturing, in contrast, offers a low-burden option for users to bypass these costs by moving production to the point-of-use, particularly for bioreagents no longer under intellectual property protection.

Beyond ensuring affordable consumables, building capacity for biotechnology relies on other equally critical elements. These include infrastructure, human resource development, and funding investments to support self-sustaining biofabrication, allowing communities to operate independently over time. To minimize reliance on capital-intensive laboratory infrastructure, our work was paired with low-burden and open-source hardware. Central to this work is the use of our hand-powered 3D centrifuge (3D-fuge, $3 USD) (**Fig. 2c**)(*45*) for low-burden protein purification and a diagnostic reader (FluoroPLUM, ∼$500 USD) (**Fig. 6f**). This device reads multi-well diagnostic tests anywhere using battery-operated onboard computing, image-based quantification, cloud connectivity for global operation, and data management(*29, 87*). With these tools and low-burden CFPS reactions, we launched disease diagnostic programs for 16 clinically relevant pathogens. This initiative ultimately supported patient trials targeting SARS-CoV-2, CHIKV, and OROV (116 samples) in four countries worldwide, with performance comparable to RT-qPCR and aligned with the WHO’s criteria for novel point-of-care diagnostic tests (**Fig. 7**, **Tables S6-12)**(*97*).

To the best of our knowledge, this study is the first to translate cell-free biomanufacturing from the lab to real-world use across diverse international settings, including those historically with limited access to the bioeconomy. This was made possible through high-contact and inclusive collaboration (biweekly meetings, two-way exchanges) across the six countries. Materials and knowledge transfer across research hubs not only fostered mutual progress but also encouraged the sharing of diverse cultural perspectives and expertise to address global and local challenges with appropriately designed solutions(*98, 99*). The support from both local and international funding agencies was crucial to the scientific capacity built here. Along with funding agencies, the active engagement of governments, philanthropic institutions, and private-sector partners is vital to sustain and expand such efforts on a global scale(*100*).

As biotechnology advances solutions to global challenges, this report, along with the work of others(*26, 27, 101–105*), provides examples of what extending research capacity can do to reshape global health and life sciences, foster equity, and redefine what is possible. Moreover, these approaches also hold potential to advance education, environmental monitoring, materials science, national security, astropharmacy, personalized medicine, and agriculture(*25, 37, 106–109*). In this context, we hope our study will serve as a model for how interdisciplinary and cross-border knowledge sharing can accelerate global scientific progress and empower local communities **(Supplementary Note 4)**.

Given their immense potential, the transformative biotechnologies that are emerging for human health must be delivered equitably, empowering all populations to shape their own health priorities. While better distribution is a short-term solution, the root of the problem is that many needs are regional (e.g., mosquito-borne infections), and when the capacity to develop solutions is not available, needs are left unmet, or pricing set beyond local means. This has been recently highlighted by the WHO(*41*) and commentaries(*10, 100*). With a clear-eyed focus on building research capacity for low-resource environments, this project developed and demonstrated tools for deployable biomanufacturing to bolster hands-on molecular training and health research capacity. More broadly, these tools seek to make all research and healthcare more resilient to global trade and supply chain disruptions while accelerating life sciences and applied research. With their low cost and operational simplicity, we see our platforms and similar disruptive technologies(*26, 101, 103, 106, 110*) as part of a new generation of tools that will help shape a future where bioreagents, advanced diagnostics, and life-saving therapeutics are accessible to all.

## METHODS

### Experimental models

#### Mice

Female and male BALB/c mice (6 weeks) were obtained from The Jackson Laboratory (strain no. 000651, USA). Mice were maintained on a standard diet (*ad libitum*) in an accredited facility under controlled temperature (22 ± 3 °C), humidity (50 ± 20%), and with a light / dark cycle of 12 h each. The animal study protocols were approved by the Institutional Animal Care and Use Committee of the University of Texas Medical Branch (UTMB; protocol number #1708051A). Animal experiments were conducted in accordance with the National Institutes of Health Guide for the Care and Use of Laboratory Animals.

#### Mammalian cell culture

Mammalian cells were maintained with 5% CO_2_ at 37 °C in a conventional incubator. The type of culture medium varied according to the specific cell line. Vero CCL-81 (ATCC; CCL-81) and NIH 3T3 mouse fibroblast (ATCC; CRL-1658) cells were cultured in Dulbecco’s Modified Eagle’s Medium (DMEM) (Gibco). Vero E6 (ATCC; CRL-1586) cells were cultured in Minimum Essential Medium (MEM) (Gibco). TF-1 (ATCC; CRL-2003) cells were cultured in RPMI 1640 Medium (Gibco). Primary human CD3+ T cells (ImmunoCult) were cultured using ImmunoCult-XF T Cell Expansion Medium (STEMCELL Technologies).

#### Bacterial strains

*E. coli* BL21(DE3) (NEB, C2527I), ClearColi BL21(DE3) (Biosearch Technologies, 60810-1), *E. coli* SHuffle (NEB, C3028J), *E. coli* BL21 (DE3)-Gold-ΔLac (Addgene, 99247), and *E. coli* BL21(DE3) Star/CRISPRi+(*76*) strains were used for preparing cell-free lysates*. E. coli* BL21 (DE3)-Gold-ΔLac has a deletion of the endogenous lac operon and is the strain used to produce cell-free lysates in combination with *lacZ*-containing genetic circuits. *E. coli* NEB 5-alpha (NEB, C2987I) was used for plasmid cloning and purification. Bacterial strains were grown in Luria broth (LB) medium (except when preparing cell-free lysates) at 37 °C in conventional shaking incubators.

### Experimental procedures

#### General template design and preparation for cell-free expression

DNA sequences encoding proteins of interest were obtained from Addgene or the scientific literature, codon-optimized for *E. coli* (Integrated DNA Technologies (IDT) Codon Optimizer), and purchased as linear gene fragments or plasmids from Twist Bioscience, unless otherwise specified. For enzyme expression, coding sequences (CDS) were inserted in the T7p14 backbone with either an N- or C-terminal His_6_ and/or Strep- tag II. For growth factor and vaccine manufacturing, screening was performed using linear DNA templates flanked by terminal Ter sites as described previously(*111*). Gene fragments were amplified by PCR using High-Fidelity DNA Polymerase (NEB, M0491L) following the manufacturer’s protocols and purified using a QIAquick PCR Purification Kit (Qiagen, 28106) before cell-free expression **(see Extended Methods)**. Following screening, top DNA-performing designs were synthesized and cloned into plasmids (Twist Bioscience). Plasmid DNA was purified using the EZNA Plasmid Midi Kit (Omega Bio-Tek, D6904) for CFPS reactions. All DNA sequences used are available **in Supplementary Data 1-2.**

#### Virus strains and preparation

Oropouche virus (OROV-AL11_2024-08-13), chikungunya virus (PE2016-480), and SARS- CoV-2 (46519/Brazil/PE-FIOCRUZ-IAM4372/2021) strains used in this study were isolated from patient samples in Brazil. After isolation, all viruses were propagated and similarly titrated using the 50% Tissue Culture Infectious Dose (TCID_50_) method with titers ranging from 10^5^ to 10^7^ TCID_50_/mL. Then, aliquots were prepared and stored at - 80 °C before use.

#### *Plasmodium falciparum* culture and conventional microscopy

Parasites (*Plasmodium falciparum*; 3D7A strain) were maintained in continuous culture as previously described(*112*) with fresh A+ red blood cells (RBCs) obtained from healthy blood donors in a culture media containing 2.5 mg/ml Albumax as previously described(*113*). Rings and mature parasites were obtained from a parasitized RBC culture synchronized on a Percoll gradient, as previously described(*112*). Non-parasitized RBCs were maintained under the same culture conditions as the parasitized RBCs and served as a negative control. Blood smears were visualized using gold-standard microscopy-based methods.

Blood was obtained from healthy donors under a protocol approved by the Research Ethics Board of the University of Toronto (protocol number #22556), which required verbal informed consent from all donors.

#### Leishmania braziliensis culture

Promastigotes were cultured in Schneider medium (SERVA, 4752) for six to seven days at 25–26°C. After this period, the parasite was harvested and processed for use in molecular reactions.

#### Nucleic acid (DNA and RNA) extraction

DNA was extracted from cultured pathogens using the QIAamp DNA Mini Kit (Qiagen, 51304) according to the manufacturer’s instructions. Viral RNA was extracted from supernatants of virus-infected cells or patient samples (nasopharyngeal swabs for SARS-CoV-2 or serum for arbovirus testing) using the QIAamp Viral RNA Mini Kit (Qiagen, 52906) following the manufacturer’s instructions. Processed DNA and RNA were stored at –20 °C and –80 °C, respectively, prior to downstream applications.

#### Toehold switch and LAMP primer design

A set of 142 toehold switch sensors was generated using an integrated in silico design algorithm **(see Extended Methods, Supplementary Data 3)**(*39*). Design specifications for the toehold switch sensors can be found at: https://github.com/AlexGreenLab/TSGEN. Genome sequences were aligned using MAFFT (version 7)(*114*), and LAMP primers were designed using the PrimerExplorer V5 software (https://primerexplorer.jp/lampv5e/index.html), unless otherwise specified. All DNA sequences are available **in Supplementary Data 5.**

#### Sensor platform building

Toehold switch sensors were assembled using conventional molecular tools. In brief, toehold switch constructs were first amplified from DNA templates (IDT) using PCR and then inserted into a pCOLA-Duet backbone in frame with the *lacZ* reporter gene using NEBuilder® HiFi DNA assembly. For initial sensor screening, each toehold switch sensor was tested against its corresponding trigger elements using PURExpress (NEB, E6800L) according to the manufacturer’s protocols, as described previously(*29, 39*). Subsequently, the top-performing sensor was selected for further optimization to work in in-house cell-free lysates. In-house reactions were assembled essentially as described in our previous efforts(*76*) and supplemented with a phosphoenol pyruvate (PEP)-based energy buffer and a nucleotide solution(*27*). In brief, reactions were assembled on ice in a final volume of 15 μL and contained circular plasmid-based DNA inputs encoding the corresponding toehold switch (2nM) and trigger ssDNA (4-8 μM) or RNA (3000 nM). To extend efforts to use linear DNA encoding toehold switches as input for reactions using in-house lysates, Ter sites were incorporated by the forward and reverse primers via PCR **(Supplementary Data 4)**, as described previously(*111*).

#### Synthetic target preparation for molecular reactions

Synthetic nucleic acid controls were synthesized as linear fragments or cloned into pUC57 (Twist Bioscience, IDT or Sangon Biotech Co.) **(Supplementary Data 5,6).** Each target was PCR amplified using Q5 High-Fidelity DNA Polymerase (NEB, M0491L). To generate RNA targets, first a T7 promoter was added to the DNA template during the PCR step, the linear DNA was then used as template in an *in vitro* transcription reaction performed using the HiScribe T7 Quick High Yield RNA Synthesis Kit (NEB, E2050S) according to the manufacturer’s instructions and primers listed in **Supplementary Data 7.** RNA samples were treated with DNase I (NEB, M0303A), purified using the RNeasy MinElute Cleanup Kit (Qiagen, 74204), and DNA and RNA were stored at –20 °C and - 80°C, respectively. Full-length synthetic RNA representatives of the SARS-CoV-2 variants and other respiratory pathogens were obtained from Twist Bioscience.

#### In-house cell-free lysate preparation

Lysates used to activate toehold sensors were prepared using *E. coli* BL21(DE3)-Gold-ΔLac following the protocols described previously (*76, 115*) with minor modifications **(Supplementary Note 1)**. Lysates used for protein expression were prepared as described previously(*44*); however, Solution A was made without putrescine. Lysates were aliquoted, flash-frozen in N_2(l)_, and stored at −80 °C until use. Freeze-thaw cycles were avoided. Due to the multi-site nature of this work, some minor modifications to the protocol were required based on the locations. **See Supplementary Information for more details.**

#### Lyophilization of molecular components

Cell-free lysates were flash-frozen as described previously **(see Extended Methods)**(*39, 43*). After lyophilized, packaged lysates were kept at room temperature or shipped to the corresponding laboratories worldwide without cold chain logistics. DNA templates were added during pellet rehydration as necessary.

#### Cell-free protein synthesis (CFPS) reaction setup

CFPS reactions using in-house lysates were assembled on ice as described previously with minor modifications(*44*). Briefly, reactions contained 33.3% crude lysate, 14.7 % buffer A, and 14.0 % buffer B containing phosphoenol pyruvate (PEP). CFPS reactions using commercial cell-free lysates, including the NEBExpress cell-free *E. coli* protein synthesis system (NEB, E5360S), S30 T7 extract-based kit (Promega, L1130), and Juice (Liberum, CF0001.B), were prepared according to the manufacturer’s protocols. For initial screening of DNA constructs, 10 μL reactions were assembled in 0.2-mL thin-walled tubes containing 15 nM of the appropriate linear DNA construct supplemented with Tus protein or GamS nuclease inhibitor (NEB, P0774S) as described previously(*111*). To assist in the folding of proteins with multiple disulfide bonds, PURExpress® Disulfide Bond Enhancer (NEB, E6820S) was used as needed. To scale up protein production, 0.5– 1 mL CFPS reactions were carried out in 15 or 50-mL conical tubes and incubated with shaking at 80 rpm. For the expression of growth factors, CFPS was conducted at 21°C for 16 hours. For all other proteins, CFPS was performed at ambient temperature (24 °C) for 16 hours. CFPS reactions were centrifuged at 20,000 x g for 5 minutes to remove insoluble protein fractions and aggregates from the supernatant.

#### Centrifugation-based affinity purification

Protein purification was carried out using NEBExpress Ni spin columns (NEB, S1427L) and/or Strep-TactinXT 4 Flow high-capacity resin (IBA, 2-5010-010) according to the manufacturer’s instructions. For His_6_-tagged products, proteins were eluted in two fractions of 150 μL each using elution buffer (20 mM sodium phosphate pH 7.4, 300 mM NaCl, and 500 mM imidazole). For Strep-tagged products, proteins were eluted in three fractions of 100 μL each using an elution buffer containing 100 mM Tris-/HCl (pH 8.0), 150 mM NaCl, 1 mM EDTA, and 50 mM biotin. Purified Bst LF and M-MLV were adjusted with 25 % glycerol, aliquoted, flash-frozen in N_2(l)_, and stored at –80 °C. Therapeutic proteins were eluted in a conventional buffer, followed by endotoxin removal (except when expressed in ClearColi^TM^ cell-free lysates), then buffer-exchanged to PBS pH 7.4, adjusted with 25 % glycerol, and stored at -80 °C. Protein quantification was performed using the Pierce BCA protein assay kit (Thermo Fisher, 23225), and absorbance was measured with a microplate reader (BioTek) at 562 nm. All fractions collected during purification were analyzed via SDS-polyacrylamide gel electrophoresis (SDS-PAGE). The Color Prestained Protein Standard, Broad Range (10–250 kDa; NEB, P7719) was used as a molecular weight marker in SDS-PAGE. Signal intensity was determined for the appropriate bands using the ImageJ software (Version 1.53k).

#### LAMP and RT-LAMP reaction setup

Reactions were carried out in triplicate using designated pipettes and filter tips. LAMP or RT-LAMP reactions were assembled in a 10 μL final volume containing primer mix at a final concentration of 1X (0.2 μM for F3 and B3 primers, 1.6 μM for FIP and BIP primers, 0.4 μM for LF and LB primers), unless otherwise noted. Isothermal and salt buffers were prepared according to these online protocols (https://www.protocols.io/view/low-costlamp-and-rt-lamp-bsejnbcn), and all oligos were synthesized by IDT **(Supplementary Data 5)**.

In-house LAMP/RT-LAMP reactions were first optimized **(see Extended Methods).** After optimization, the optimal conditions for all parameters were selected for further experiments. In brief, LAMP reaction mixtures contained 1X isothermal buffer (20 mM Tris-HCl, 10 mM (NH4)2SO4, 50 mM KCl, 2 mM MgSO4, 0.1% Tween 20, pH 8.8), 4 mM MgSO_4_, 1.4 mM deoxynucleotides triphosphates (dNTPs) (NEB, N0446S), 10X primer mix, and our in-house produced Bst DNA Polymerase Large Fragment (5.46 ng/μL for LAMP and 7.31 ng/μL for RT-LAMP reactions). For RNA detection, reactions were supplemented with the Moloney Murine Leukemia Virus Reverse Transcriptase (M-MLV) (2.15 ng/μL, **see Supplementary Note 3 for more details**). To this mixture, 1.0 μL of template (nuclease-free water, synthetic DNA/RNA inputs, extracted DNA/RNA, or supernatants of virus-infected cells) was added. To benchmark in-house reactions, WarmStart LAMP 2X master mix (NEB, E1700S) reactions were simultaneously prepared according to the manufacturer’s instructions.

Reactions were assembled in 96-well or 384-well formats and then incubated using a qPCR instrument (QuantStudio 3 or 5, Applied Biosystems, USA), FluoroPLUM (LSK Technologies, now part of Nicoya), or a conventional thermal cycler at an isothermal temperature (60-65 °C), followed by inactivation at 80 °C for 5 minutes (**see Supplementary Table S5 for more details)**. Results were visualized using three approaches: real-time fluorescence monitoring, visual colorimetric detection, and gel electrophoresis **(see Extended Methods)** (*13, 87*).

#### qPCR and RT-qPCR assays

For qPCR reactions, QuantiNova SYBR Green PCR master mix (Qiagen, 208052) and corresponding primer sets were prepared in a final volume of 10 μL. For RT-qPCR, samples were assayed according to protocols recommended by the US Centers for Disease Control and Prevention (CDC) or the Pan American Health Organization (PAHO)(*93–95*). RT-qPCR reactions were performed using the QuantiNova Probe RT-PCR Kit (Qiagen, 208354) according to the manufacturer’s instructions. Briefly, each reaction was prepared to a final volume of 10 μL with standardized primer and probe concentrations of 0.8 μM for forward and reverse primers, 0.1 μM for the probe, and 3.5 μL of template (nuclease-free water, extracted RNA, or *in vitro* transcribed RNA). All reactions were assayed in 96- or 384-well plates using the Applied Biosystems QuantStudio 3 or 5 instruments. Primers and probes for each pathogen were synthesized by IDT and are listed in **Supplementary Data 6**.

#### Cell-based proliferation assays

Cell-based proliferation assays were conducted to evaluate the functional activity of our in-house-produced growth factors in comparison to their commercial counterparts. In brief, FGF-1, IL-3, and IL-15 were tested in NIH-3T3 mouse embryonic fibroblast cells, TF-1 human erythroleukemia cells, and primary CD3+ T cells, respectively **(see Extended Methods for more details).** To evaluate cell proliferation, the CellTiter-Glo® 2.0 Cell Viability Assay (Promega, 9242) was used to measure ATP levels, which serve as a direct indicator of the number of viable cells in culture(*116*). Following the protocol, plates were read for luminescence using a BioTek microplate reader. Commercial growth factors used in this study included recombinant human FGF1 protein (SinoBiological, 10013-HNAE), recombinant IL-3 protein (PeproTech, 200-03), and recombinant human IL-15 protein (SinoBiological, 10360-HNAE).

#### Mice immunization and IgG antibody detection by ELISA

Animals were randomly divided into three groups for assessment of immunogenicity (n = 5 per group). Mock treatment mice received PBS alone. Another mouse group received only adjuvants, including Alhydrogel 2% (500 μg for each; InvivoGen, vac-alu-50) and CpG-ODN 2395 (50 μg for each; InvivoGen, vac-2395-1). The third and last mouse group were immunized with a Nuvax formulation containing the cell-free-based nucleocapsid antigen (1 μg per animal per injection) plus CpG-ODN 2395 (50 μg per mouse per injection) and Alhydrogel 2% (500 μg per mouse per injection) (ratio 1:50:500). Injections (50 μL) were administered intramuscularly (thigh muscles of the hind limb) on days 0 (prime) and 15 (boost). Blood samples were collected on days 7, 14, 21, and 28 after treatment and used to test for anti-SARS-CoV-2 nucleocapsid IgG Ab production. Vaccine-induced, N-specific binding IgG in mice serum samples (diluted 1:100) was measured by indirect ELISA using the SARS-CoV-2 nucleocapsid protein IgG Ab ELISA Kit (ABclonal, RK04178) as described in the manufacturer’s instructions. Plates were read at 450 nm wavelength using a microplate reader (BioTek).

#### Western blot analysis

For Western blot analysis, 1 μL of CFPS reactions was loaded onto SDS-PAGE 4–20% Mini-Protean TGX Precast Protein Gels (Bio-Rad, 4561093 and 4561096) followed by transfer to nitrocellulose membranes (0.45 μm) using a Trans-Blot Turbo transfer system (Bio-Rad). Briefly, membranes were blocked with TBST + 5% skim milk for 1 h at room temperature. Membranes were then probed with an anti-His-HRP antibody (Southern Biotech, 4603-05) diluted in TBST (1:10000) for 1 h at room temperature. Following this step, membranes were washed three times with TBST with a 5-minute waiting time after each wash. They were then developed with Clarity Western ECL substrate (Bio-Rad, 1705061) for 3-5 minutes and imaged using a ChemiDoc MP imaging system (Bio-Rad). For nucleocapsid antigens, membranes were probed with an anti-SARS-CoV-2 nucleocapsid primary antibody (SinoBiological, 40588-T62) and an anti-rabbit IgG (HRP) (Abcam, ab6721) secondary antibody with 1:2500 and 1:5000 dilutions, respectively. The SARS-CoV-2 nucleocapsid recombinant protein (SinoBiological, 40588-V07E) was used as a positive control to evaluate the expression of the vaccine antigen in cell-free lysates.

#### Endotoxin removal and quantification

Endotoxin removal was done using the Pierce High-Capacity Endotoxin Removal Spin Column Kit (Thermo Fisher Scientific, 88274) according to the manufacturer’s instructions, unless otherwise noted. Following this step, endotoxin levels were measured using the Pierce Chromogenic Endotoxin Quantification Kit (Thermo Fisher Scientific, A39552). Endotoxin-free ultra-pure water (Sigma-Aldrich, TMS-011-A) was used during all steps.

#### FluoroPLUM fabrication

FluoroPLUM (LSK Technologies, now part of Nicoya) was fabricated as previously reported(*29*), with some modifications. To enable fluorescence-based detection, bandpass filters were incorporated into the system. All hardware design files (laser-cut, 3D-printed, schematics) from the previous version are available at https://github.com/PardeeLab/zikaproject_hardware. GUI and workflow code are at https://github.com/PardeeLab/zikaproject_plumcode.

#### 3D-fuge fabrication

Hand-powered centrifuges (3D-fuges) were fabricated as described previously, with minor modifications(*45*). Briefly, 3D-fuge components were printed using an Ultimaker 3 extended extrusion 3D printing machine (Ultimaker, Netherlands) with Polylite (PLA) filament (2.85 mm diameter). Neodymium magnets (1/8-inch thick, 1/4-inch diameter, McMaster-Carr, OH, USA) were attached to the printed parts using Gorilla Glue (OH, USA). A 104 cm length of Dorisea Extreme Braid 500 lb 2.0 mm fishing line was coiled 50 times around the centrifuge before operation. Design specifications for the 3D-fuge used here are available at: https://github.com/PardeeLab/3D-fuge-PardeeLab-2025.

### Data analysis

All experiments were performed independently in at least three biological replicates, each containing three technical replicates. Data are shown from one representative experiment out of three conducted with similar results (mean ± SD, n = 3). Statistical analyses (Student’s t-test and ANOVA) were performed on GraphPad Prism 10 (GraphPad Software). MedCalc software (version 19.2.0, MedCalc Software, Belgium) was used to perform the probit analysis for analytical sensitivity experiments. Diagnostic parameters(*117*) were established using an online tool provided by MedCalc (https://www.medcalc.org/calc/diagnostic_test.php).

### Patient sample collection

In Canada, nasopharyngeal swab samples were collected from individuals suspected of having respiratory illness at the clinical diagnostics laboratory of Mount Sinai Hospital in Toronto. In Brazil, nasopharyngeal swabs and serum samples were employed for genomic surveillance and later provided for this study. In Chile, nasopharyngeal swab samples were obtained from individuals during the pandemic and processed by the Laboratory of Microbiology at the Medical Center of Pontificia Universidad Católica de Chile, which later provided the RNA samples for this study. In Colombia, nasopharyngeal swab samples were collected previously for the Uniandes COVIDA project and later provided for this study.

### Ethics approval for patient trials

This study was approved by the research ethics board (REB) at the University of Toronto under the protocols 46252 and 39531 for arbovirus and COVID-19 studies, respectively. Sampling and sample testing was approved by the Pernambuco State Haematology and Hemotherapy Foundation (HEMOPE-PE, Brazil), Institutional Review Board (IRB) (CAAE: 43877521.4.00000.5195), by the Federal University of Alagoas (UFAL, Brazil) IRB (CAAE: 65701122.8.0000.5013), by the ethics committee from Universidad de los Andes (Comité de Ética de la Investigación de la Universidad de los Andes) under the approval number 1181, and by the ethics committee from the Pontificia Universidad Católica de Chile (Comite Ético Científico de Ciencias de la Salud UC) under the approval number 200624012. Given the nature of this project, all patient trials were conducted in full compliance with local and international regulations, including the ethical principles for medical research involving human samples as outlined in the World Medical Association’s Declaration of Helsinki.

### Data availability

The main data supporting the results in this study are available within the paper and its Supplementary Information. All the experimental raw data are provided as a Source Data file.

## Supporting information

This PDF file includes: Additional experimental details Supplementary Notes 1 to 4 Figs. S1 to S29 Tables S1 to S12 References.

## Data Availability

The main data supporting the results in this study are available within the paper and its Supplementary Information.

## Acknowledgements

We thank the members of all participating laboratories involved in this global research network for their technical support, fruitful discussions, and invaluable feedback. We thank Gustavo Barbosa de Lima, Keilla Maria Paz e Silva, Mayara Matias de Oliveira Marques da Costa, Diego Arruda Falcão, Andreza Pâmela Vasconcelos, and all other health professionals for their invaluable support in processing the patient samples used in Canada, Chile, Brazil, and Colombia. We thank Mônica Beatriz Mathor for providing the NIH-3T3 mouse embryonic fibroblast cell line used in the cell proliferation assays conducted in Brazil. We sincerely thank the staff of all participating institutions for their exceptional support. In particular, we appreciate their assistance with issuing official documents that enabled team members to travel internationally, as well as their warm hospitality and efforts to receive and accommodate visiting researchers throughout the project’s course. In addition to our efforts, we appreciate the dedication of other researchers and authorities worldwide who are working to build biotechnology capacity to advance global equity.

## Author contributions

S.J.R.d.S. designed and performed experiments, co-supervised the project, and co-wrote the manuscript. Q.M., S.C., and J.R.J.V. designed and performed experiments and co-wrote the manuscript. S.J.R.d.S, Q.M., S.C., J.R.J.V., B.N.R.S., D.D., P.B.B., T.K., A.L.L.D., A.A., and V.F. performed most molecular-based experiments. Q.M. adapted the 3D- printed centrifuge (3D-fuge) for cell-free protein purification. M.S. modified the 3D-fuge by replacing the nut-and-bolt closure with a magnetic closure. S.J.R.d.S. and L.A.C. designed and built growth factor constructs. S.J.R.d.S., L.A.C., T.Y.C., and K.B. designed and conducted affinity-based growth factor purification and cell-based experiments. S.J.R.d.S, Q.M., and J.R.J.V. designed and built vaccine constructs. S.J.R.d.S, Q.M, D.C.M.C., S.C.W., and K.P. designed and planned vaccine-related experiments. D.C.M.C. performed animal work for the vaccine trial. K.W. and A.A.G. designed the toehold switches. S.J.R.d.S. integrated the Tus-Ter protection system with the toehold switches. Q.M., A.T., and A.K. designed and built diagnostic enzyme constructs. S.J.R.d.S., J.N., and A.H. designed the LAMP primers. S.J.R.d.S. developed and implemented an in-house LAMP/RT-LAMP diagnostic workflow for pathogen detection. Y.G., S.C., and K.P. developed the FluoroPLUM diagnostic reader. K.A. and I.A. provided *P. falciparum* cultures. T.P.G.d.A., J.P.M.d.N., J.J.F.d.M., M.G.S., P.G.C., M.H.S.P., T.M., G.L.W., and A.S.J. contributed to patient sample collection and acquisition. S.J.R.d.S., S.C., J.R.J.V., B.N.R.S., D.D., P.B.B., R.P.G.M., C.L., S.C.d.L., L.C.M., and L.K. contributed to the patient trials., S.S., J.D., P.B., L.K., A.R., I.A.I., M.C., and A.B. supported the project. S.J.R.d.S., Q.M., S.C., J.R.J.V., B.N.R.S., D.C.M.C., L.C., D.D., and K.W. performed data analysis, interpretation, and wrote the manuscript. S.S. and J.D. helped write and review the manuscript. K.P. supervised the project, designed experiments, and co-wrote the manuscript, along with L.P., F.F., C.G., A.A.G., and C.A. All authors edited and approved the final version of the manuscript.

