## Supplementary material for "International Multi-site Implementation of Local Cell-Free Protein Biomanufacturing to Advance Health and Research Equity": This PDF file includes: Additional experimental details Supplementary Notes 1 to 4 Figs. S1 to S29 Tables S1 to S12 References.

*Silva et al.*

**This PDF file includes:**

Additional experimental details

Supplementary Notes 1 to 4

Figs. S1 to S29

Tables S1 to S12

References

**Other Supplementary Materials for this manuscript include the following:**

Supplementary Data 1 to 7 (DNA sequences)

Appendix 1 (Cost breakdown)

Appendix 2 (Material list)

Movie S1 (Showcasing decentralized biomanufacturing in low-resource settings)

### Additional experimental details

#### Lyophilization of molecular components

CFPS reactions were flash-frozen as described previously(*1*). Sucrose at 771 mM was used as a cryoprotectant. In brief, CFPS reactions were placed in a microcentrifuge tube with a parafilm lid or a lid with three holes punched into it. The tubes were placed upright in a rack and partially submerged in liquid nitrogen for 5 minutes to pre-freeze the samples. Following this, the rack and tubes were removed from the liquid nitrogen and transferred to a glass lyophilization chamber. The chamber was connected to a freeze-drying machine (Labconco), which had been pre-cooled to −80 °C, and the samples were lyophilized overnight under vacuum. After lyophilization, the glass chamber was disconnected from the lyophilizer, and the tubes were immediately flushed with nitrogen gas to prevent moisture absorption. The dried samples were then placed into vacuum-sealable bags, along with three desiccant packs and two oxygen absorbers. The bags were purged with nitrogen gas and sealed using an impulse heat vacuum sealer.

#### Computational design of toehold switches

An updated version of the selection algorithm described previously(*2, 3*) was used to identify toehold switches. The algorithm facilitated the selection of six promising designs from a set of 142 candidate toehold switches generated from each target RNA. Candidate sensors were designed to bind to the single-stranded loop region of the LAMP product. Putative toehold switches were generated at 1-nt increments along the target RNA, and multiple ensemble defect levels were computed for each sensor based on its deviation from the ideal secondary structure of the toehold switch. Ensemble defects were calculated for the toehold switch 5′ end through to the 3′ end of the hairpin (*d*min_sensor), the toehold domain of the toehold switch (*d*toehold), the binding site of the toehold switch within the target RNA (*d*binding_site), and the toehold switch region starting with the base immediately 3′ of the target RNA binding site and extending beyond the last base on the 3′ end of the hairpin (*d*active_sensor). The parameter *d*active_sensor was intended to provide a measure of any secondary structures in the activated toehold switch that could interfere with translation after binding to the target RNA.

In addition to ensemble defects, the equilibrium fraction *f* of target/toehold switch complexes in a system with equimolar concentrations of target and toehold switch RNAs was calculated as a measure of the affinity of the two RNAs. In practice, this parameter was almost always equal to 1. Designs that produced in-frame stop codons in the output gene were eliminated from further consideration. Each parameter was then normalized such that its maximum value across the set of putative designs for a given target RNA was equal to 1. These normalized parameters, designated by an overscore, were then inserted into a scoring function *s*. Toehold switches displaying the lowest *s* values and screened to have $f > 0.9$ were selected for experimental testing.

**Design of the toehold switch for LAMP amplicons**

One challenge in integrating the LAMP assay for toehold-switch diagnostics is that the toehold switches were previously designed to target linear ssRNA. Although, in theory, they can work with ssDNA, it has not been demonstrated before. Plus, the ssDNA region in the dumbbell-shaped LAMP reaction amplicons is within the loop region, which might make it difficult for toehold switches to bind.

To design toehold switches that would work for the ssDNA loop region of LAMP products, we extended the toehold domain used to initiate strand-displacement reactions. This change should encourage the binding between the ssDNA target and the toehold switch RNA. To increase the likelihood of getting a functional switch, we designed a total of 142 switches targeting SARS-CoV-2 spike, nucleocapsid, open Reading Frame 1b (ORF1b), as well as human 18s rRNA and β-actin mRNA as a human endogenous control. The screening has yielded switches with an ON/OFF ratio above 10. Within them, we selected a top-performing SARS-CoV-2 spike toehold switch (D07) for further characterization.

**Linear DNA template preparation for cell-free expression**

PCR was used to amplify gene fragments using primers targeting the Ter sites (the primers used for PCR **are listed in Supplementary Data 7)**. PCR reactions were assembled using Q5 High-Fidelity DNA Polymerase (NEB, M0491L) according to the manufacturer's protocols for a 50-µL final volume. Reactions were carried out in a ProFlex thermocycler (Applied Biosystems) with a program consisting of a single cycle of initial denaturation for 30 s at 98 °C, 35 cycles of 10 s at 98 °C, 20 s at 72 °C, and 30 s at 72 °C, followed by a final extension step for 5 min at 72 °C. Following amplification, the

PCR product was purified using a QIAquick PCR Purification Kit (Qiagen, 28106). Gel electrophoresis was then used to verify the DNA quality before proceeding to cell-free expression.

**Cell-based protein expression**

The Tus protein (Addgene, 165959) was expressed and purified as described previously(*4*), with the minor modification that *E. coli* BL21 (DE3)-dLac strain was used instead of *E. coli* BL21 (DE3) cells.

**Optimization of the LAMP/RT-LAMP assay**

To optimize an in-house LAMP/RT-LAMP for DNA and RNA detection with equivalent performance to commercial reagents, various reaction settings, including enzyme concentration, $Mg^{2+}$ concentration, dNTP concentration, and the addition of GuHCl (Sigma-Aldrich, G3272-25G), were tested. After optimization, the optimal conditions for all parameters were selected for further experiments. Then, the optimized assays were evaluated and benchmarked side by side against a commercial kit in Canada, Chile, Brazil, and Colombia.

**Visualization of LAMP amplicons**

The amplicons were visualized for fluorescence measurements at the indicated time points by adding 1x LAMP fluorescent dye (NEB, B1700S) or 1x EvaGreen (Biotium, 31000). The qPCR reading was set for SYBR green reading, and read every minute. In the experiments performed with FluoroPLUM, 10 µM SYTO 9 Green Fluorescent Nucleic Acid dye (Invitrogen, S34854) was used(*5*). For visual detection, amplification products were visualized by the naked eye under natural light by adding 1.5 µL of SYBR Gold Nucleic Acid Stain (Invitrogen, S11494) diluted 1:10 in nuclease-free water to the center of the tube caps before reaction incubation and mixing, as described previously(*6*). A colour change from orange to green indicated a positive sample, while a negative reaction remained orange. At the end of the reactions, a smartphone camera was used to photograph the reaction tubes, and the amplicons were analyzed using agarose gel electrophoresis (1.5%).

**Cell-based proliferation assays**

**FGF-1 proliferation assay**

NIH-3T3 mouse embryonic fibroblast cells were seeded at a density of 6 x $10^3$, cells per well into a black opaque-walled 96-well plate with DMEM + 10% FBS in a 37 °C $CO_2$

incubator. 24 hours later, the medium was replaced, and the cells were starved with DMEM + 0.11% FBS. After 48 hours, the medium was replaced with 50 µL of DMEM, followed by dilutions ranging from 1 to 1000 ng/mL. After 72 h, the plate was left to stabilize at room temperature for 30 min. CellTiter-Glo® 2.0 Cell Viability Assay (Promega, 9242) was equilibrated to room temperature, and 100 µL/well was added to the cells (1:1 ratio to culture medium). The plate was placed on a thermomixer for 2 min at 300 rpm, incubated at room temperature for 10 min, and luminescence was read using a BioTek Neo microplate reader.

**IL-3 proliferation assay**

TF-1 cells were seeded at a density of 1 x $10^4$ cells per well into a black opaque-walled 96-well plate and then treated with various concentrations of IL-3 ranging from 0.025 to 5 ng/mL. After 72 hours, the plate was left to stabilize at room temperature for 30 minutes. CellTiter-Glo® 2.0 Cell Viability Assay (Promega, 9242) was thawed and equilibrated to room temperature, and then 100 µL/well was added to cells (1:1 ratio to culture medium). The plate was placed on a thermomixer for 2 minutes at 300 rpm and then incubated at room temperature for 10 minutes. Luminescence was read using a BioTek Neo microplate reader.

**IL-15 proliferation assay**

T cells were seeded at a density of 1 x $10^4$ cells per well into a black opaque-walled 96-well plate and then treated with various concentrations of IL-15 ranging from 1 to 500 µg/mL. Cells were incubated for 72 hours at 37°C in a $CO_2$ incubator. After 72 hours, the plate was left to stabilize at room temperature for 30 minutes. CellTiter-Glo® 2.0 Cell Viability Assay (Promega, 9242) was thawed and equilibrated to room temperature, and then 100 µL/well was added to cells (1:1 ratio to culture medium). The plate was placed on a thermomixer for 2 minutes at 300 rpm. The plate was then incubated at room temperature for 10 minutes. Luminescence was read using a BioTek Neo microplate reader.

**SARS-CoV-2 nucleocapsid vaccine design**

The coding sequence (CDS) of the nucleocapsid protein from the Severe Acute Respiratory Syndrome Coronavirus 2 (SARS-CoV-2) isolate Wuhan-Hu-1 (GenBank: MN908947.3). This CDS was optimized for expression in E. coli K12 using the Integrated DNA Technologies Codon Optimization Tool for gBlocks synthesis **(Supplementary Data**

**1).** The optimized sequence was then input into the RBS Calculator v2.2 (design mode) to maximize the target translation initiation rate in *E. coli* BL21(DE3) (NZ_CP053602)(*7*). The top-performing design was selected and synthesized as a clonal gene (Twist Bioscience) **(Supplementary Data 2)**.

**Supplementary Note 1**

**Optimized protocol for cell-free lysate preparation for integration with toehold switch-based sensors**

This protocol was adapted from the following sources:

Sun, Z. Z., Hayes, C. A., Shin, J., Caschera, F., Murray, R. M., Noireaux, V. Protocols for Implementing an *Escherichia coli* Based TX-TL Cell-Free Expression System for Synthetic Biology. *J. Vis. Exp*. (79), e50762, doi:10.3791/50762 (2013).

Levine, M. Z., Gregorio, N. E., Jewett, M. C., Watts, K. R., Oza, J. P. Escherichia coli-Based Cell-Free Protein Synthesis: Protocols for a robust, flexible, and accessible platform technology. J. Vis. Exp. (144), e58882, doi:10.3791/58882 (2019).

Guzman Chavez, F, Haseloff, J. Solutions for CFPS (Version 1.1)- Haseloff Lab V.2. Protocols.io. doi:10.17504/protocols.io.bmi2k4ge (2020). <https://www.protocols.io/view/solutions-for-cfps-version-1-1-haseloff-lab-dm6gprk71vzp/v2>

Guzman Chavez, F, Haseloff, J. Cell-free extract, 4x Wizard mix and CFPS reaction preparation- Haseloff Lab V.3. Protocols.io. doi: 10.17504/protocols.io.bu3xnypn (2021). <https://www.protocols.io/view/cell-free-extract-4x-wizard-mix-and-cfps-reaction-q26g7bk23lwz/v3>.

Arce Medina, A. Step 1: Preparing S12 cell extracts using bead-beater. Protocols.io. doi:10.17504/protocols.io.ieacbae (2017). <https://www.protocols.io/view/step-1-preparing-s12-cell-extracts-using-bead-beat-ieacbae>

Guzman-Chavez F, Arce A, Adhikari A, Vadhin S, Pedroza-Garcia JA, Gandini C, Ajioka JW, Molloy J, Sanchez-Nieto S, Varner JD, Federici F, Haseloff J. Constructing Cell-Free Expression Systems for Low-Cost Access. ACS Synth Biol. 2022 Mar 18;11(3):1114-1128. doi: 10.1021/acssynbio.1c00342 (2022).

Mullin, A.C., Slouka, T., Oza, J.P. Simple Extract Preparation Methods for E. coli-Based Cell-Free Expression. In: Karim, A.S., Jewett, M.C. (eds) Cell-Free Gene Expression. Methods in Molecular Biology, vol 2433. Humana, New York, NY. doi: 10.1007/978-1-0716-1998-8_2 (2022).

### Materials

#### Cell lysate preparation

**Table 1**. Materials needed for the preparation of cell lysates.

| Material/Solution | Quantity | Volume | Concentration |
|---|---|---|---|
| Flask | 5 | 2 L | |
| Nalgene flask for centrifugation | 4 | 500 mL | |
| Sterile 2xYT media | | 2L | See composition |
| Sterile LB media | | 100-200 mL | |
| IPTG | | 2 mL | 1 M |
| Sterile Potassium monobasic phosphate | | 20 mL x L (can be prepared in higher quantity and stored at 4°C) | 1 M |
| Sterile Potassium dibasic phosphate | | 40 mL x L (can be prepared in higher quantity and stored at room temperature - freezes at 4°C) | 0.5 M |
| S30B | | approximately 300 mL x batch (can be prepared in higher quantity and stored at 4°C) | See composition |
| DTT | | < 2mL | 1M |
| Sterile $dH_2O$ | | approximately 150 mL x L | |

CPFS precursor solutions:
The PEP-based energy buffer and Tx-TL solutions required for performing the CFPS reaction are prepared according to the following protocol without any substantial modification:

https://www.protocols.io/view/solutions-for-cfps-version-1-1-haseloff-lab-dm6gprk71vzp/v2

This is relevant for the last section about performing a cell-free reaction once the extracts have been prepared.

**Methods**

**Solutions for cell-extract preparation**

**Table 2**. Solutions needed for the preparation of cell lysates.

| **Solution** | **Materials** | **Instructions** |
|---|---|---|
| 2xYT | Tryptone<br>Yeast Extract<br>NaCl | Weight 16 g of tryptone, 10 g of yeast extract and 5g of NaCl. Dissolve in 800 mL of $dH_2O$ and autoclave. Prepare 2 bottles of 800 mL of sterile 2xYT for this protocol. |
| Potassium Monobasic Phosphate (1 M) | For 500 mL preparation | Dissolve 78.005 g in 400 mL $dH_2O$ first and then gauge to 500 mL. Around 60 mL should be necessary to reach the final volume. Autoclave. |
| Potassium Dibasic Phosphate (0.5 M) | For 1 L preparation | Dissolve 134.035 g in 700 mL $dH_2O$ under heating and magnetic stirring. In our case we used 75°C x 240 rpm until dissolution. Gauge to 1000 mL with $dH_2O$. Autoclave. |
| S30B | Hemimagnesium Glutamate<br>Potassium Glutamate<br>Tris 2 M<br>$dH_2O$ | Dissolve 5.44 g of hemimagnesium glutamate and 12.195 g of potassium glutamate in 700 mL $dH_2O$. Adjust the pH to 8.2 with Tris 2 M. Gauge to 1000 mL with $H_2O$. Autoclave. |

**Step-by-step protocol for cell-free lysate preparation**

Strain: *E. coli* BL21DE3 GoldJM1 dLac

Overexpressed protein: T7 RNA Polymerase

Day 1:

**Cell culture**

1. Inoculate a culture tube from the strain's glycerol stock with 5 mL of sterile LB media and proper antibiotics. In this case, we used Kanamycin (stock: 50 mg/mL) and Carbenicillin (stock: 50 mg/mL), 5 µL each. Prepare a negative control culture (LB + antibiotics + sterile tip without the strain).
   Alternatively, prepare an LB agar plate with both antibiotics and inoculate the plate with the strain's glycerol stock to obtain separate colonies. *Note: By taking this step, it will be defined as Day 1 of the protocol.*
2. Incubate overnight at 37°C x 210 rpm.

Day 2:

1. Scale the culture into a sterile 1000 mL flask, inoculating 100-200 mL of sterile LB medium with antibiotics with the culture or a single colony from Day 1. Prepare a negative control of the culture (a culture tube with sterile media and antibiotics, plus a tip without the strain should be sufficient).
2. Incubate overnight at 37°C x 210 rpm. Calculate the incubation time so that no more than 16-17 hours of incubation elapse until the next inoculation.

Day 3:

1. Reconstitute the 2xYT media with phosphates. To do so, add 20 mL of sterile monobasic phosphate and 40 mL of sterile dibasic phosphate in a laminar flow chamber or under sterile conditions.
2. Mix by inverting the bottles a couple of times.
3. Add 1mL of both antibiotics (Carbe50 and Kan50). Mix again by inversion.
4. Inoculate each bottle with the culture from Day 2, so that the culture has an initial $OD_{600}$ of 0.1.
   *Note: Some protocols specify that inoculation should be performed at an optical*

*density (OD) of 0.05, while others recommend an OD of 0.1. We haven't seen differences in extract efficiency, but $OD_{600}$ 0.1 saves time.*

5. Add sterile $dH_2O$ to bring the culture to a final volume of 1000 mL in each bottle.
6. Pour 400 mL of the culture into each 2 L flask and incubate at 37°C, 210 rpm.
7. Measure the $OD_{600}$ after 60-90 minutes and then every 20-30 minutes until the $OD_{600}$ reaches 0.4 - 0.6.
8. Induce all cultures with IPTG 1 mM (final concentration). *Note: Other protocols employ a gentler induction with IPTG at 0.4 mM to prevent toxicity. We haven't encountered a specific issue using IPTG at 1 mM, but this is something to consider.*
9. Incubate again at 37°C x 210 rpm for 90 minutes. The final $OD_{600}$ should reach 2-2.5.
10. During the induction, ensure that all necessary items are ready for the subsequent steps. In particular: i) prepare ice, ii) Make sure the S30B is cold (on ice or at 4°C), iii) Cool down the centrifuge to 4°C for further recollection and washing of the cell cultures, iv) prepare DTT 1M if it wasn't ready.

**Cell recollection and washing**

11. After 90 minutes, incubate the culture on ice to stop the growth. Pour the 2 L culture into 4 x 500 mL Nalgene bottles.
12. Centrifuge at 4°C for 10 to 20 minutes at 4500 rpm, until the supernatant appears clear.
13. During centrifugation, reserve 300 mL of cold S30B and add 300 µL of 1 M DTT. Mix by inverting the bottle a couple of times.
14. For each Nalgene bottle, discard the supernatant and resuspend the pellet in 50 mL of cold S30B + DTT.
15. Centrifuge at 4500 rpm x 12 min (4°C) and discard the supernatant.
16. Resuspend each pellet in 10 mL of cold S30B + DTT.
17. Pool all fractions in a cold and clean 50 mL Falcon tube.
18. Centrifuge at 4500 rpm x 12 min (4°C) and discard the supernatant.
19. Meanwhile, weigh a void, cold, and clean 50 mL Falcon tube.

20. When centrifugation ends, weigh the tube with the pellet and subtract the Falcon tube’s weight to get the humid pellet’s mass. *Note: For a 2 L culture, you should obtain a mass of at least 8-10 g of humid pellet.*

**Lysis method: Bead-beating**

21. For each g of pellet, add 0.9 mL of cold S30B with additional DTT (2 µL per mL in total) and 5 g of beads. <u>Important:</u> Separate the beads into 3 weighing boats and add them one by one, vigorously homogenizing between each addition. The third edition will be challenging to homogenize; however, it is crucial to ensure that beads are evenly distributed in the cell suspension through vigorous vortexing.
22. Cut the end of a p5000 tip in a slight diagonal orientation to enlarge the diameter of the tip end. As if it were a pastry bag, pour the cell and bead mixture with the cut tip into 2.5 mL culture tubes. Tubes should be filled, ensuring no air bubbles, up to the edge of the tube, with a flat meniscus. Pour some cell mixture at the center of the cap, so that when closing the tube, there is no air between the cap and the tube. See Sun et al(*8*).

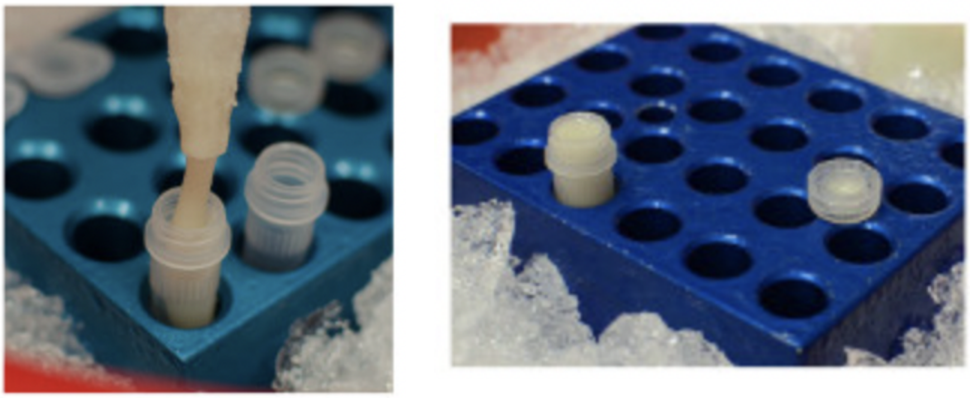

**Figure 1.** Pouring the 2.5 mL culture tube. Adapted from Sun et al(*8*).

23. Beat the beads for 30 seconds using a Fast-Prep Bead-Beater (or equivalent). Use safety glasses when operating the bead-beating machine.
24. Build a filter apparatus according to the instructions provided by Sun et a(*8*).

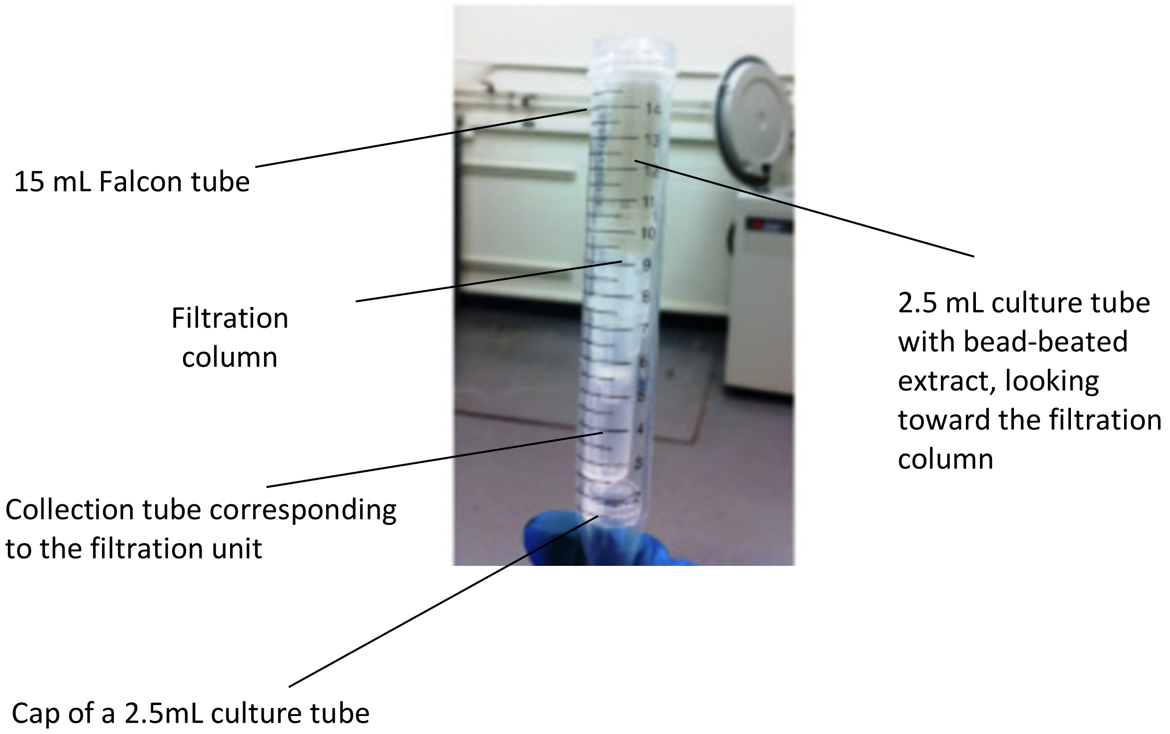

**Figure 2**. Filtration apparatus. Adapted from Sun et al (*8*).

25. Centrifuge at 4°C, 4700 rpm x 25 minutes.
26. If the lysis and filtration were performed correctly, two distinguishable phases should appear: a translucent supernatant (containing the proteins) and a turbid pellet (comprising cell debris). See below.

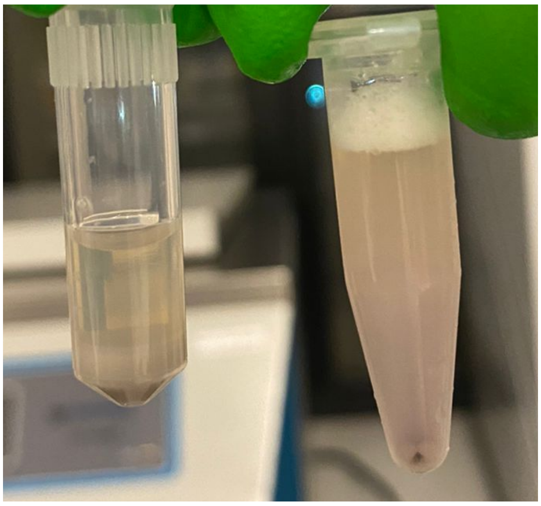

**Figure 3**. On the left: a cell extract prepared with bead-beating. Two phases can be distinguished, the supernatant being the phase of interest. On the right: a cell extract prepared with sonication.

27. Rescue the supernatant from each tube, taking care not to disturb the pellet, and transfer it into clean 1.5 mL tubes.
28. Centrifuge at 4°C for 10 minutes at 12000 rpm.
29. If no post-lysis method will be used, go to step 29. Otherwise, go to the post-lysis section.
30. Collect the supernatant without disturbing any pellets in clean 1.5 mL tubes and keep at -80°C.

**Post-lysis: Run-off and dialysis**

*This step reproduces Sun et al.(8), with minor modifications.*

1. After step 27, collect the supernatant without disturbing any pellets and pour into clean 1.5 mL tubes.
2. Incubate at 37°C for 60 minutes. This run-off reaction allows the Tx-Tl machinery to get free of any current Tx-Tl process and be available for subsequent cell-free reactions.
3. Centrifuge at 4°C for 10 minutes at 12000 rpm.
4. Collect the supernatant without disturbing the pellets and pour it into clean 1.5 mL tubes.
5. Prepare a 1 L beaker with a magnetic stirrer, containing 900 mL of cold S30B and 1 mM DTT. Hydrate one or two 10k MWCO dialysis cassettes for a couple of minutes.
6. Load the extracts in the dialysis cassette, up to 2.5 mL per cassette.
7. Keep the system on ice and dialyze for 60 minutes. This dialysis step should clean the extract buffer and remove small proteins and molecules from the extract.
8. After dialysis, remove the extract from the cassette and pour it into clean 1.5 mL tubes.
9. Store at -80°C. Flash-freezing is not critical if you do not have liquid nitrogen (we never flash-freeze our extracts for this reason).

***Note: It has been observed twice that freezing the pellet and proceeding to lysis the day after is detrimental to cell-free lysate activity. Therefore, splitting the protocol is not recommended. This applies to protocols that use bead-beating for lysis. Other protocols with sonication permit freezing the pellet.***

**Cell-free reaction for toehold switch D07 interaction with synthetic trigger:**

A cell-free reaction will have the following components:

i) Lysate extract;

ii) Tx-Tl solutions and energy buffer;

iii) Substrate for *lacZ* gene reporter: chlorophenol red-beta-D-galactopyranoside (CPRG);

iv) D07 toehold switch (DNA);

v) Trigger (DNA and RNA);

vi) Nuclease-free water;

**Table 3**. Composition of a cell-free reaction for D07 interaction

| **Reagent** | **Volume (µL)** |
|---|---|
| CFE | 5 |
| Wizard 4X PEP | 4 |
| CPRG (15mg/mL) | 1 |
| DNA | 1.2 nM |
| RNA | 0.8-3 µM |
| Nuclease free-water | up to 15 µL |
| **Total** | 15 µL |

**Supplementary Note 2**

### Guidelines for performing a FITC calibration curve

#### FITC preparation

- Fluorescein Isothiocyanate (https://www.thermofisher.com/order/catalog/product/46425)
- DMSO (https://itwreagents.com/italy/en/product/dimethyl+sulfoxide+%28reag.+usp%2C+ph.+eur.%29+for+analysis%2C+acs/131954)
- PBS 5X
- Amber tubes
- Analytical Balance
- Micropipettes and tips

#### FITC measurement

- Plate reader (Ref: Biotek Synergy HTX) with optic filter 485/20ex, 516/20em or equivalent
- 384-well flat-bottom dark optic plate (dark on the sides but with optic wells) + corresponding seals

#### Methods

**1) FITC preparation**

1.1. Fluorescein Isothiocyanate is a powder that must be kept in the dark at -20°C.

1.2. Keeping a liquid stock of FITC is not recommended as it is unstable over time. However, for short periods (weeks), it is possible to make a temporary stock using DMSO and keep it in the dark at -20°C.

1.3. Prepare a 10 mM stock of FITC by dissolving the corresponding amount of powder in DMSO. Prepare eight dilutions of the stock in PBS 1X, as follows. The stocks can be prepared in 1.5 mL amber tubes (or traditional clear tubes foiled with aluminum) and stored at -20°C.

**Table 1**. FITC concentrations used for the calibration curve.

| 1 | 2 | 3 | 4 | 5 | 6 | 7 | 8 |
|---|---|---|---|---|---|---|---|
| PBS 1X (blank) | 0.01 µM | 0.33 µM | 0.1 µM | 0.33 µM | 1 µM | 3.33 µM | 10 µM |

**2) FITC measurement**

2.1. Technical triplicates of 5 µL for each concentration were loaded in a 384-well flat-bottom plate. This means that, in total, we used 16 wells, each with 5 µL, to cover the 8 points of the calibration curve.

2.2. Before measuring CFE fluorescence, it is recommended to measure FITC alone with the plate reader to ensure that the signal does not saturate, even at the highest concentration of FITC. If it saturates, it is recommended to lower the gain so that all the fluorescent signals can be detected.

2.3. FITC was loaded simultaneously with sfGFP samples in cell-free reactions (concentrated at 2 nM for sfGFP), and the kinetics were measured at 37 °C for 180 minutes, monitoring fluorescence through the green channel with the following characteristics: excitation, 485/20 nm; emission, 516/20 nm.

**3) Data analysis**

3.1. Plotting the raw data of FITC samples to ensure the signal is stable over time.

3.2. Plotting a boxplot for each concentration incorporates mean, quartiles, min, max, and median. This also allows us to observe how dispersed the measurements are over time for each concentration.

3.3. Determining the linear range of the data. A log-log scale will probably have to be used to make the determination of this linear range easier to identify.

3.4. Performing a linear regression with at least 5 points in the linear range.

The resulting equation will be associated with an $R^2$ parameter to indicate how good the fit is. It should have the following form:

log([FITC]) = a*log([Fluorescence]) + b

Then, in the analysis of fluorescent samples, equivalent FITC units can be recovered by the formula:

[FITC] equivalent = 10**(a*log(fluorescence of the sample) + b)

Example of calibration curve

Here, we attach a shareable code for building a FITC calibration curve:

https://colab.research.google.com/drive/19R8og3T3leZdIJYdyHa-rVzztRsmXa9h?usp=sharing

Below are the graphs obtained from the analysis of the fluorescence intensity of each FITC concentration.

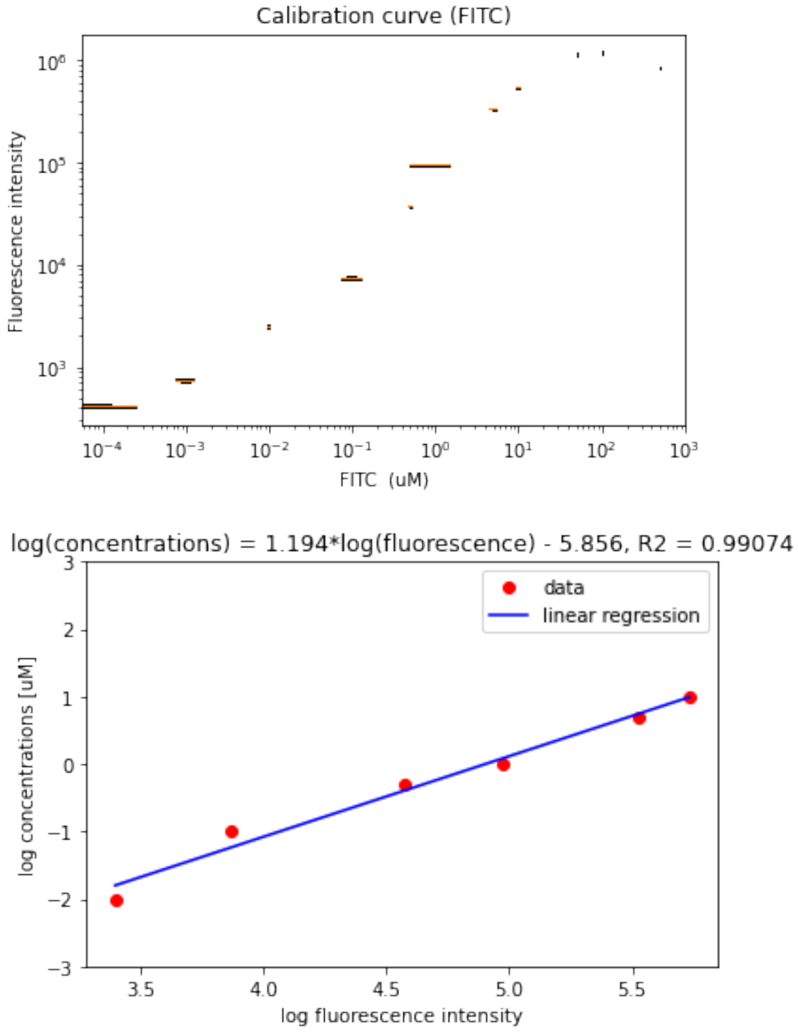

**Figure 1**. Left: box plots of fluorescence intensity with their respective FITC concentrations. Right: Linear interpolation over the linear range of FITC concentrations.

**Supplementary Note 3**

**Standard operating procedures manual for the production of diagnostic enzymes**

**Main goal**

- Instruct users on expressing Bst LF and M-MLV in CFPS reactions.
- Instruct users on performing in-house LAMP/RT-LAMP reactions.

**Instruments**

- Plate reader;
- qPCR machine;
- FluoroPLUM;

**Materials (please see the material list, Appendix 2);**

**Protocol**

- CFPS reactions were prepared as described by Levine et al(*9*).

1) **Bst LF and M-MLV expression**

   1.1. Bst LF and M-MLV are expressed in cell-free lysates prepared from *E. coli* BL21(DE3).

   1.2. *E. coli* BL21-based cell-free lysates and CFPS reactions are prepared as described by Levine et al(*9*). *Note: PEP has been used as an energy source.*

   1.3. Resuspend the CFPS reaction in nuclease-free water and add plasmid DNA to a final concentration of 15 nM. Use a vortex to mix the reaction and move it to 15-mL or 50-mL sterile Falcon tubes.
   *Note: This reaction is typically prepared for a final volume of 500 µL.*

Please see the table below:

| **Component** | **Volume (µL)** | **Final concentration** |
|---|---|---|
| Solution A | 73.5 | 14.7% |
| Solution B | 70.0 | 14.0% |
| Cell-free lysate | 166.5 | 33.3% |
| Nuclease-free water | x | to reach the final volume |
| Template (DNA) | x | 15 nM |
| Total | 500 µL | |

1.4. Incubate all reactions at 24°C for 14-16h (overnight).

1.5. Mixtures should be incubated with gentle shaking at 80 rpm.

1.6. Following overnight expression, proceed with protein purification.

**2) Bst LF and M-MLV purification**

- Bst LF and M-MLV have been purified using the NEBExpress® Ni Spin Column Reaction Protocol (NEB #S1427). *Note: This protocol was established using the NEB website as a primary reference: (https://www.neb.com/en-ca/protocols/2019/08/28/nebexpress-ni-spin-column-reaction-protocol-neb-s1427).*
- It is recommended that the expression of the tagged protein of interest be confirmed by first running a sample on an SDS-PAGE gel.
- During the entire protein purification process, keep all tubes on ice. If available, perform all centrifugation steps at 4 °C to preserve protein stability.

Prepare all required buffers according to the protocol below:

2X IMAC Buffer (0.04 M Sodium Phosphate, 0.6 M NaCl, pH 7.4).

2M Imidazole (2M Imidazole, pH 7.4).

| | Lysis/Binding Buffer: 20 mM sodium phosphate; 300 mM NaCl, pH 7.4; | Wash Buffer: 20 mM sodium phosphate; 300 mM NaCl; 5 mM Imidazole, pH 7.4; | Elution Buffer: 20 mM sodium phosphate; 300 mM NaCl; 500 mM Imidazole, pH 7.4; |
|---|---|---|---|
| 2X IMAC Buffer | 7.5 mL | 5.0 mL | 2.5 mL |
| 2M Imidazole | - | 0.025 mL | 1.25 mL |
| Water | 7.5 mL | 5.0 mL | 1.25 mL |
| Total | 15.0 mL | 10.0 mL | 5.0 mL |

2.1. Column Preparation

2.1.1. Twist to remove the bottom tab of the column, loosen the top cap, and place the column into the provided collection tube.

2.1.2. Centrifuge column at 800 x g for 1 minute to remove and discard the storage buffer.

2.1.3. Add 250 µL of lysis/binding buffer directly to the column.

2.1.4. Centrifuge the column at 800 x g for 1 minute (except when using the battery or 3D centrifuges), then discard the lysis/binding buffer.

2.1.5. Place the column in a new 2 ml microcentrifuge tube.

2.2. Lysate Binding

2.2.1. Before protein purification, centrifuge the CFPS reactions at 18000-20000 x g for 5 minutes to remove protein aggregates. *Note: Save 5 µL of the supernatant in a new 2 mL microcentrifuge tube.*

2.2.2. Add 500 µL of the protein sample extract directly to the column.

2.2.3. Tap the column to mix the lysate with the resin, and then allow binding to occur for 2-3 minutes. *Note: Prolonged mixing may result in more nonspecific binding proteins.*

2.2.4. Centrifuge the column at 800 x g for 1 minute and collect the flow-through.

2.2.5. Place the column in a new 2 ml microcentrifuge tube.

2.3. Column Wash

2.3.1. Add 250 µL of wash buffer to the column and centrifuge at 800 x g for 1 minute.

2.3.2. Repeat this wash step twice, collecting each wash in a separate 2 ml microcentrifuge tube, for a total of three washes.

2.4. Protein Elution

2.4.1. Place the column in a new 2 ml microcentrifuge tube.

2.4.2. Add 150 µL of elution buffer to the column.

2.4.3. Centrifuge at 800 x g for 1 minute to collect the first elution.

2.4.4. Repeat the elution step by adding 150 µL of elution buffer to the column and centrifuging at 800 x g for 1 minute to obtain the second elution. *Note: typically, >90% of the bound protein is eluted following the second elution.*

2.4.5. Combine elutions #1 and #2 in a 2 mL Eppendorf tube, yielding approximately 300 µL of combined elution.

*2.4.6.* Adjust the eluted protein to include glycerol. *Note: The final enzyme stock will contain 25% glycerol.*

*2.4.7.* Prepare aliquots containing 20 µL of Bst LF and M-MLV and store the final protein stock at -80 °C until use.

2.4.8. Analyze the clarified cell lysate (load), flow-through, washes (1, 2, and 3), and elution fractions by SDS-PAGE.

2.5. SDS-PAGE

2.5.1. Upon protein purification, run an SDS-PAGE gel to confirm protein expression.

2.5.2. Heat all samples at 98 °C for 5 minutes—load 2-5 µL for each sample.

2.5.3. Run the gel at 180V for 45 minutes.

2.5.4. See Figure S15 for an example of results.

#### 3) Protein quantification

CFPS products were quantified using the Pierce™ BCA Protein Assay Kit (Thermo Scientific, 23227). *Note: The assay should be done in duplicate or triplicate, and the protocol described here was adapted from the ThermoFisher Scientific website (https://www.thermofisher.com/order/catalog/product/23227).*

3.1. Preparation of diluted albumin (BSA) standards

- To prepare the protein standards, dilute the contents of one 2 mg/mL Albumin Standard (BSA) ampule into several clean 1.5 mL Eppendorf tubes using the same diluent as your samples. *Note: Refer to the table below for guidance on preparing a dilution series ranging from 2000 µg/mL to 25 µg/mL.*
- You may optionally use other protein quantification methods (e.g., Bradford assay, Nanodrop). However, remember that you may need to perform a screening step to determine the optimal enzyme concentration for your molecular assay.

Preparation of diluted albumin (BSA) standards:

| **Preparation of albumin standard curve (2000-25 µg/mL)** | | |
|---|---|---|
| Final BSA concentration (µg/mL) | Volume of diluent* (µL) | Volume and source of BSA (µL) |
| A - 2000 | 0 | 300 µL of stock |
| B- 1500 | 125 | 375 µL of stock |
| C- 1000 | 325 | 325 µL of stock |
| D- 750 | 175 | 175 µL of vial B dilution |
| E- 500 | 325 | 325 µL of vial C dilution |
| F- 250 | 325 | 325 µL of vial E dilution |
| G- 125 | 325 | 325 µL of vial F dilution |
| H- 25 | 400 | 100 µL of vial G dilution |
| I-0 (Blank) | 400 | 0 |

*The diluent used is the NEB elution buffer.

3.2. Preparation of the BCA working reagent:

3.2.1. Use the following formula to determine the total volume of WR required:

(# standards + # unknowns) × (# replicates) × (volume of WR per sample) = total volume WR required

Example: for the standard test-tube protocol with 2 unknown samples, each measured in duplicate (2 replicates), the total WR volume should be calculated as follows:

(9 standards + 2 unknowns) × 2 replicates = 22 wells, plus 2 extra wells for pipetting error; therefore, a total of 4.8 mL of WR is required.

3.2.2. Prepare WR by mixing 50 parts of BCA Reagent A with 1 part of BCA Reagent B (50:1, Reagent A:B). For the above example, combine 4,705 mL of Reagent A with 94 µL of Reagent B.

*Note: After the initial addition of Reagent B to Reagent A, a transient turbidity may appear, which quickly disappears upon gentle mixing, resulting in a clear, green working reagent (WR).*

3.3. Microplate procedure

3.3.1. Pipette 10 µL of each standard or unknown sample replicate into a 96-well plate (working range = 2000-25 µg/mL, plus blank). *Note: All samples should be tested in duplicate.*

3.3.2. Dispense 200 µL of the WR into each well. Mix thoroughly using a plate shaker for 30 seconds to ensure homogeneity.

3.3.3. Cover the plate with aluminum foil and incubate at 37°C for 30 minutes.

3.3.4. Cool the plate to RT (37°C). Then, use a microplate reader to measure the absorbance at or near 562 nm.

See Figure below for an example of results:

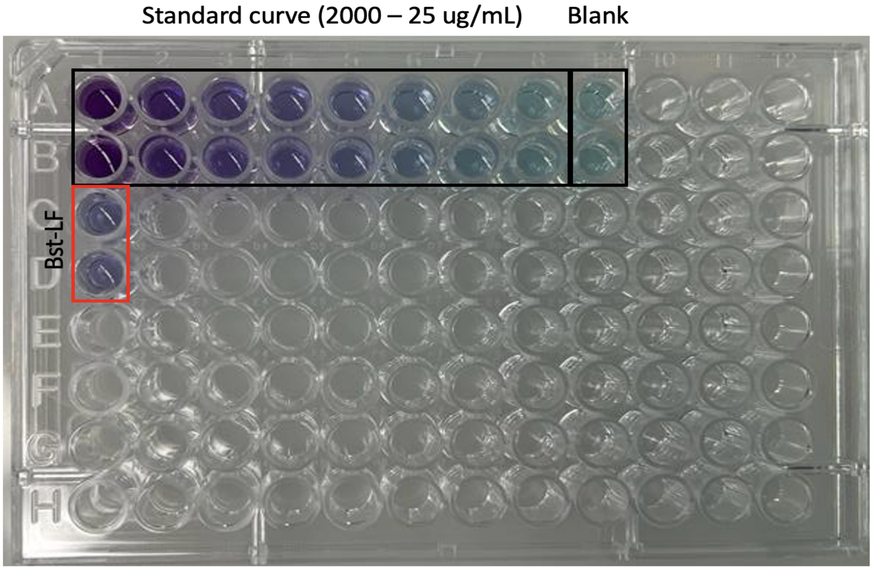

4) **Preparation of positive controls for molecular reactions**

Upon receiving the synthetic gBlocks DNA, resuspend the pellet in X µL of nuclease-free water to achieve a final 10 ng/µL concentration. Note: PCR amplification was performed using Q5® High-Fidelity DNA Polymerase (NEB, M0491).

For positive control preparation:

- DNA targets: use F3 and B3 (or forward and reverse) primers at a 10 µM stock concentration.
- RNA targets: use T7-F3 and B3 (or T7-forward and reverse) primers at 10 µM stock concentration.

Resuspend all primers to a final concentration of 100 µM.

Prepare a master mix for six reactions: one no-template control (NTC) and four reactions containing DNA.

*Note: Use the four DNA reactions for column purification. In this protocol, we used the QIAquick PCR Purification Kit (Qiagen, 28106), but similar kits may also be suitable.*

4.1. Assemble the reactions in PCR tubes on ice, following the table below:

| **Component** | **Volume (µL)** | **Final concentration** |
|---|---|---|
| 5X Q5 Reaction Buffer | 10.0 | 1X |
| 10 mM dNTPs | 1.0 | 200 µM |
| 10 µM forward Primer | 2.5 | 0.5 µM |
| 10 µM reverse Primer | 2.5 | 0.5 µM |
| Nuclease-free water | X | to reach the final volume |
| Q5 DNA Polymerase | 0.5 | 0.02 U/µL |
| DNA (gBlock) | X | < 1000 ng |
| Total | 50 µL | |

4.2. Place the reactions in a thermal cycler, following the cycling conditions listed below:

<table>
<tr><th>Condition</th><th>Steps</th></tr>
<tr><td>98 °C - 30 sec</td><td>1 cycle</td></tr>
<tr><td>98 °C - 10 sec</td><td rowspan="3">35 cycles</td></tr>
<tr><td>X °C* - 20 sec</td></tr>
<tr><td>72 °C - 15 sec</td></tr>
<tr><td>72 °C - 5 min</td><td>1 cycle</td></tr>
<tr><td>4 °C - ∞</td><td></td></tr>
</table>

*Use a primer annealing temperature calculation tool (https://nebiocalculator.neb.com/#!/dsdnaamt) based on the primers that will be used to generate the positive control for each pathogen. *Note: For DNA pathogens, use F3 and B3 primers. For RNA pathogens, use T7-F3 and B3 primers.*

4.3. Run the PCR products on a 1.5% agarose gel for analysis.

4.4. Purify the PCR products using a spin-column-based PCR purification kit. Elute the DNA in 30 µL of nuclease-free water, following the manufacturer's instructions.

4.5. Quantify the DNA using a Nanodrop. Then, determine the concentration of DNA (https://nebiocalculator.neb.com/#!/dsdnaamt) and RNA (https://nebiocalculator.neb.com/#!/ssrnaamt) using an online NEB tool.

### 5) In vitro transcription for RNA template preparation

5.1. Assemble the reaction components on ice, following the table below. We have used the HiScribe® T7 Quick High-Yield RNA Synthesis Kit (NEB, E2050S). Optional: You may use an alternative kit available in your laboratory.

| **Component** | **Volume (µL)** | **Final concentration** |
|---|---|---|
| NTP buffer mix | 10.0 | 10 mM each NTP final |
| T7 RNA Pol. mix | 2.0 | - |
| Nuclease-free water | X | to reach the final volume |
| DNA | - | 1 µg |
| Total | 20 µL | |

5.2. Incubate the IVT reactions at 37 °C for 4 hours (overnight incubation is also acceptable), followed by DNase I treatment to remove the template DNA.

5.3. Add 30 µL of nuclease-free water to each 20 µL reaction, followed by 2 µL of DNase I (RNase-free) to remove the template DNA. Mix well and incubate for 15 minutes at 37°C.

5.4. Before starting the RNA purification process, add 48 µL of nuclease-free water to bring the final volume to 100 µL. Then, proceed with RNA purification using the RNeasy MinElute Cleanup Kit (Qiagen, 74204) and elute the RNA in 14 µL of RNase-free water.

5.5. Measure RNA concentration and purity using a Nanodrop spectrophotometer. Determine the RNA concentration (https://nebiocalculator.neb.com/#!/ssrnaamt) using an online tool.

5.6. In vitro transcribed RNA products should be stored at -80°C until used in molecular reactions. *Note: Prepare small aliquots to minimize the number of freeze-thaw cycles.*

**6) In-house LAMP reaction setup**

**Essential tips for all in-house LAMP reactions:**

- Once synthetic DNA or RNA are prepared, test the activities of Bst LF or Bst LF /M-MLV. The WarmStart® LAMP Kit (DNA & RNA) is used as a standard control for comparison. Note: In this project, we used reactions with a final volume of 10 µL.
- Use nuclease-free water for all molecular experiments and procedures.
- To avoid contamination, assemble the LAMP components in an area isolated from the amplification process. Place the reagents, such as buffers, enzymes, and primers, on ice or a cold block.
- 10X Isothermal and salt buffers were prepared according to these online protocols (https://www.protocols.io/view/low-costlamp-and-rt-lamp-bsejnbcn).
- The 10X RT-LAMP primer mix includes six primers (16 µM of FIP/BIP, 4 µM FLoop/BLoop, and 2 µM of F3/B3).
- Thaw all reagents on ice. Before starting, centrifuge all molecular reagents.
- For all LAMP experiments, use only nuclease-free water.

- Prepare small aliquots of all LAMP reagents (e.g., isothermal amplification buffer, primers, dNTPs, LAMP dye, Warm Start, etc.) and store them at -20 °C until use.
- Always use filter tips in all molecular experiments to prevent contamination.
- Before setting up reactions, decontaminate the bench surface and pipettes with 2% bleach followed by 70% alcohol.
- Reactions should be performed in triplicate.

6.1. Assemble reactions (WarmStart® Kit - NEB) on ice according to the table below:

| **Component** | **Volume (µL)** | **Final concentration** |
|---|---|---|
| 2X Warm Start LAMP Kit | 5.0 | 1X |
| Primer mix (10X) | 1.0 | - |
| LAMP Dye (50X) | 0.2 | 1X |
| Nuclease-free water | X | to reach the final volume |
| Target (DNA/RNA) or water for NTC | 1.0 | - |
| Total | 10 µL | |

6.2. Assemble the in-house LAMP reaction on ice, following the instructions in the table below:

| **Component** | **Volume (µL)** | **Final concentration** |
|---|---|---|
| 10X Isothermal Amplification Buffer | 1.0 | 1X |
| $MgSO_4$ (100 mM) | 0.4 | 4 mM (6 mM total) |
| dNTP Mix (10 mM) | 1.4 | 1.4 mM |
| Primer mix (10X) | 1.0 | 1X |
| Bst LF | X | 5.46 ng/µL |
| LAMP Dye (50X) | 0.2 | 1X |
| Nuclease-free water | X | to reach the final volume |
| DNA target or water for NTC | 1.0 | - |
| Total | 10 µL | |

6.3. Assemble the in-house RT-LAMP reaction on ice, following the instructions in the table below:

| **Component** | **Volume (µL)** | **Final concentration** |
|---|---|---|
| 10X Isothermal Amplification Buffer | 1.0 | 1X |
| $MgSO_4$ (100 mM) | 0.4 | 4 mM (6 mM total) |
| dNTP mix (10 mM) | 1.4 | 1.4 mM |
| Primer mix (10X) | 1.0 | 1X |
| Bst LF | X | 7.31 ng/µL |
| M-MLV | X | 2.15 ng/µL |
| LAMP Dye (50X) | 0.2 | 1X |
| Nuclease free-water | X | to reach the final volume |
| RNA target or water for NTC | 1.0 | - |
| Total | 10 µL | |

6.4. Prepare the reaction mix in a 1.5 mL microcentrifuge tube. Mix the reaction by vortexing, spin down, and dispense 9.0 µL into each well. *Note: For all experiments, it is recommended to include a negative control (non-template control [NTC]) and a positive control (2 nM for DNA or $10^5$ copies for RNA).*

6.5. Add 1.0 µL of template (or nuclease-free water).

6.6. Place the PCR film over the top of the plate.

6.7. Centrifuge the 96-well (or 384-well) plate at 600 x *g* for 1 min.

6.8. Incubate at the optimal isothermal temperature (X °C; refer to Table S5 for specific temperatures by pathogen) for 20–40 minutes.

*Note: Preferably, incubate your reactions using a low-cost instrument (FluoroPLUM, water bath) that aligns with the resources and equipment available in your laboratory. You can also read your reactions using a conventional thermal cycler or qPCR instrument. Please note that the representative protocol described here uses LAMP dye. The protocol should be adjusted if you choose a different readout method (e.g., SYBR, SYTO-9).*
*Note: If you are using a qPCR instrument, performing a melting curve analysis will be interesting. If you use the FluoroPLUM device, the temperature will be automatically set to 65 °C.*

See the main manuscript and the supplementary information for an example of the results.

**Supplementary Note 4**

**Key tips for successful technology deployment**

- Our international research team has made significant contributions to deploying technology platforms in resource-limited settings(*5, 10, 11*). The documents presented here were initially reported in our previous effort(*10*), and we are now sharing these new templates, which have been refined over time as we expand and travel to new locations to implement our cell-free biomanufacturing platform.
- Drawing on our previous(*10*) and ongoing efforts, the following documents are shared as examples of the documentation our team prepared before conducting international fieldwork. These documents are intended as a general reference, recognizing that customs and import/export regulations for devices, reagents, and materials differ widely across countries.
- Documents (Table 1) were kept on hand during travel, with additional copies stored in checked luggage to ensure redundancy. Local authorities requested some of the documents included, while others were not. Template versions of these documents are provided and may be adapted to suit the requirements of other research teams planning technology deployment and exchange.
- It is highly advisable to verify the specific customs and legal requirements of the destination country well in advance of travel.
- For this section, "Guest" refers to the visiting research team and/or their affiliated institution conducting the fieldwork. At the same time, "Host" denotes the local researchers and/or institution in the country where the research occurs.
- All fieldwork activities were typically accompanied by workshops and symposiums designed to facilitate knowledge transfer to the local community.
- Experiments involving patient samples must obtain approval from a research ethics board.
- Previous publications, press releases, grant award letters, and related documentation serve to demonstrate that the fieldwork is part of an ongoing, credible academic research initiative. Accordingly, these items can be brought along as supporting documents during a work-related trip(*10*).

**Table 1.** Documents carried during travel for the transport of reagents, devices, and consumables.

| Document | Prepared by | |
|---|---|---|
| | Guest | Host |
| Letter of importation | | X |
| Letter of donation | X | |
| Invitation letter | | X |
| Material list (reagents, devices, and consumables) | X | |
| Ethics Approval | X | X |
| Supporting documents | X | |

(On Host institutional letterhead) (Triplicate copy in the Host country's official language)

**Letter of importation**

From: Host Principal Investigator, Department Chair, or Faculty Dean
Re: Research study in [City], [Country]

To Whom It May Concern:

[Statement introducing the author, their position within the project, the official title of the research initiative, and the timeline for its implementation. Mention of the visiting team members from the Guest institution, including their names and responsibilities within the project. Acknowledgment of the organizations providing financial or institutional support for the project.]

The project aims to [insert a clear, lay explanation of the research objectives] and complements ongoing research activities within our institution. To support the successful implementation of the project, the visiting team will transport specific scientific devices, laboratory reagents, and supplies essential to the field implementation. Please find a list of the components that will be transported on the attached page.

We respectfully seek authorization to enter the country with these materials. It should be emphasized that these materials are non-hazardous, for research purposes only, hold no commercial value, meet international safety standards, and are necessary for the project's implementation.

If you have any questions, please don't hesitate to contact me directly through the contact information at the top of the letter.

Thank you for your time and consideration in supporting this collaborative scientific effort.

Sincerely,
[Signature]

(On Host institutional letterhead) (Triplicate copy in the Host country's official language)

**Certificate of donation**

From: Host Principal Investigator, Department Chair, or Faculty Dean
Re: Research study in [City], [Country]

Please note that the devices and supplies listed in the attached appendix are being provided as a donation from [Guest institution] in [city, country] to [Host institution] in [city, country] for the ongoing research project titled "[title of research project]." I understand that these items will be utilized solely for research by [Host institution] and their collaborators.

The items will be transported by [name(s) of traveller(s)], a [role of traveller(s)], in both carry-on and checked luggage on [date of flight]. As detailed in the attached document, the consumables will be used during experiments at [Host institution], and the [names of devices] will be returned to [Guest country] after the fieldwork. No payment was made for these items/devices, and they will not be resold by [Host institution], as they hold no commercial value.

If you have any questions, please don't hesitate to contact me directly through the contact information at the top of the letter.

Sincerely,
[Signature]

(On Host institutional letterhead) (Triplicate copy in the Host country's official language)

**Invitation letter**

From: Host Principal Investigator, Department Chair, or Faculty Dean
To: Visiting Investigator

Re: Research study in [City], [Country]

Dear (Team member name),

On behalf of the [Host team], I am pleased to invite you to attend the [Symposium/Workshop name]. The meeting will be held at the [Institute name, City, Country], on [Date]. Considering the importance of this scientific meeting and the presence of experts, we have established a [date] for a symposium with topics related to [research topic].

We hope you can attend the meeting and accept our invitation to give a talk at the planned symposium. Your contribution will be invaluable to the success of this meeting and our joint project.

If you have any questions, please contact me by phone or email.

Looking forward to your reply, we will be pleased to see you in [City, Country].

Sincerely,
[Signature]

### Supplementary Figures

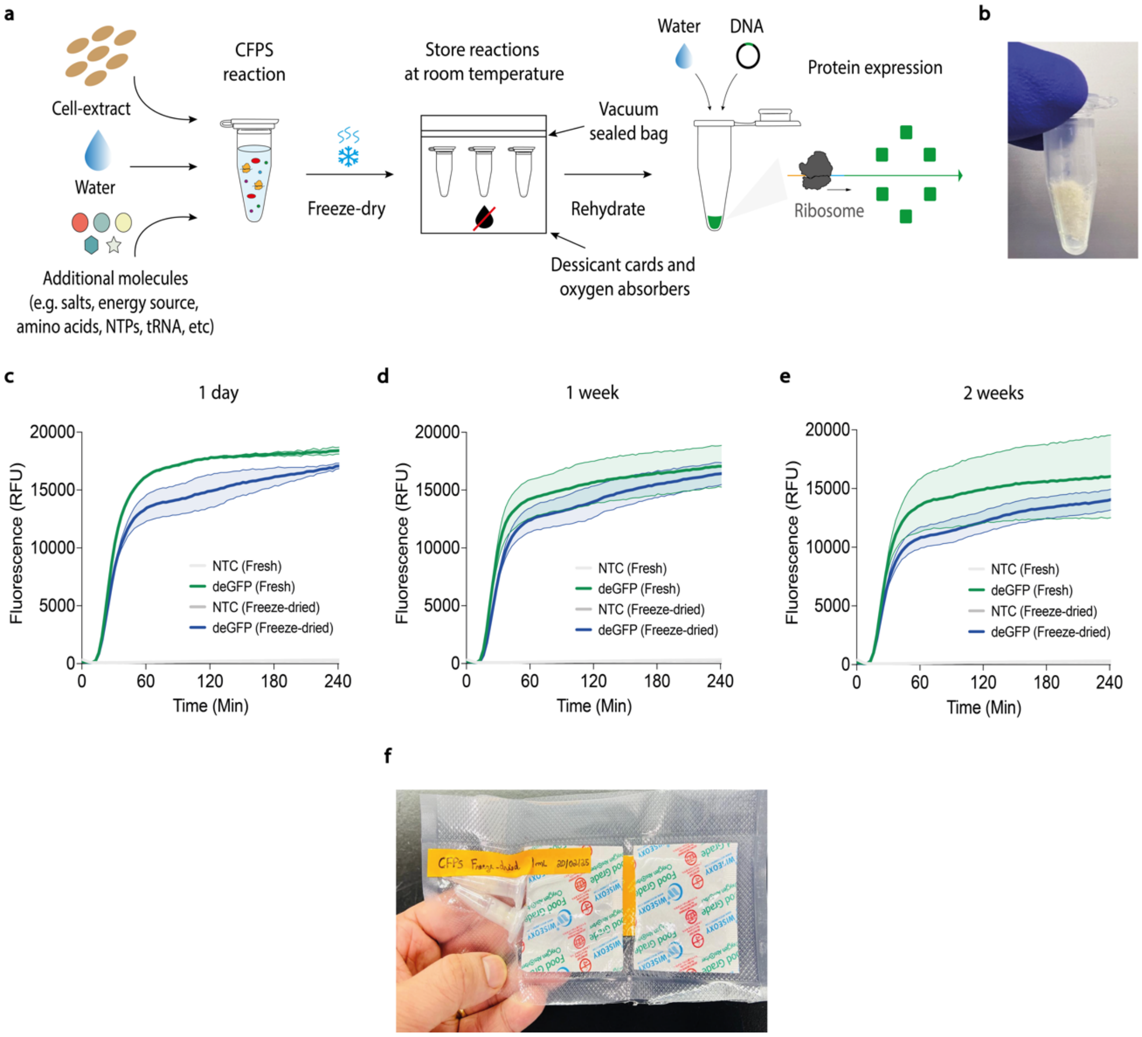

**Fig. S1: CFPS reactions can be lyophilized and stored at ambient temperature for at least two weeks, enabling global distribution across diverse settings. (a)** Schematic representation of the CFPS reaction setup and lyophilization process. CFPS reactions were freeze-dried and stored at ambient temperature (22–24 °C). At designated intervals (after one day and at 1 and 2 weeks), FD-CFPS reaction mixtures were rehydrated with water **(see Extended Methods for more details)**. **(b)** Representative image of an FD-CFPS reaction. **(c)** CFPS activity was assessed using deGFP fluorescence. FD-CFPS reactions (blue data) stored at ambient temperature are compared to fresh CFPS reactions (green data). After one day, FD-CFPS reactions were rehydrated with water. In this representative experiment, deGFP fluorescence was measured over 4 h using cell-free extracts produced on-site in Canada. Data are shown as mean ± SD, n = 3. **(d)** After one week, deGFP fluorescence was monitored over 4 h using cell-free extracts produced on-site in Canada. Data are shown as mean ± SD, n = 3. **(e)** After two weeks, deGFP fluorescence was monitored over 4 h using cell-free extracts produced on-site in Canada. Data are shown as mean ± SD, n = 3. **(f)** This image shows FD-CFPS reactions that were prepared and packaged for international shipment to team members at various locations. Abbreviations are: NTC, non-template control; CFPS, cell-free protein synthesis; Min, minutes.

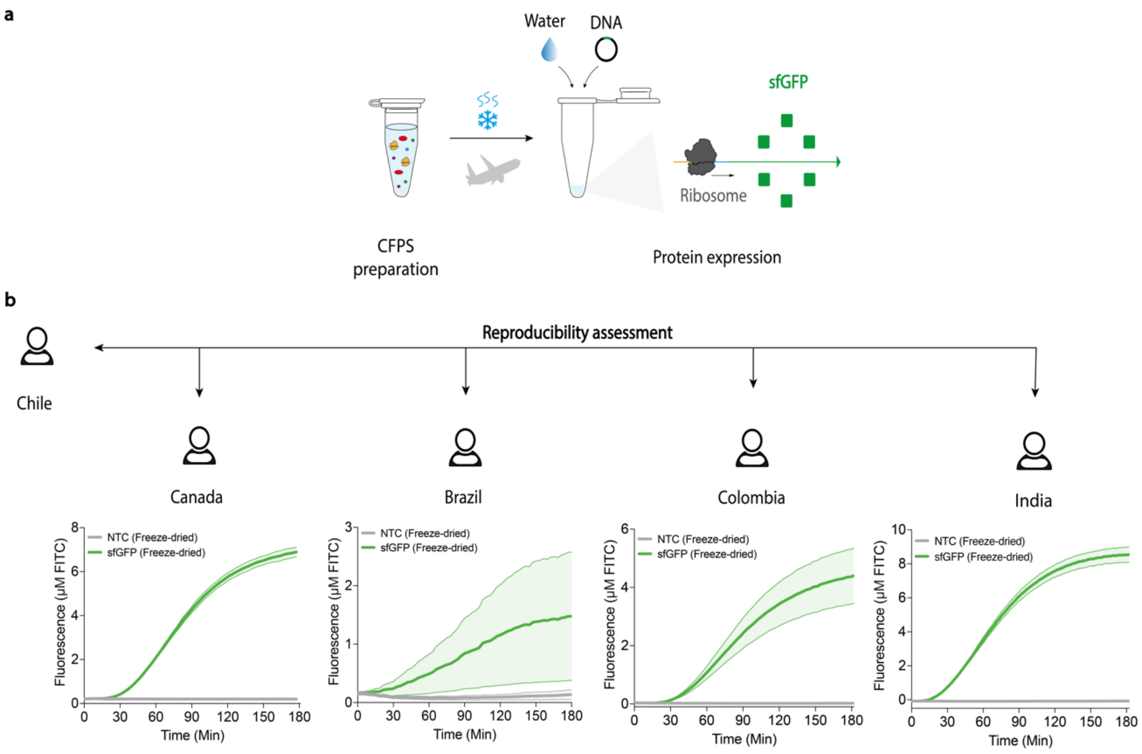

**Fig. S2: Standardized protocols and portable FD-CFPS systems enabled global distribution, enabling local protein manufacturing. (a)** Schematic overview of the CFPS reaction setup, lyophilization process, and global deployment for local protein biomanufacturing. CFPS activity was assessed using sfGFP fluorescence standardized to FITC curves, and all reactions were stored and distributed at ambient temperature using conventional logistics couriers (e.g., FedEx). **(b)** FD-CFPS reactions were shipped (prepared in Santiago, Chile) to researchers in Canada, Brazil, Colombia, and India, who were able to synthesize sfGFP after rehydration. In this reproducibility assessment, deGFP fluorescence was measured during a 3-hour reaction incubation using a conventional plate reader. While successful in Chile, Canada, Colombia, and India, test performance was inconsistent in Brazil, where shipping and customs delays impacted cell-free lysate activity, emphasizing the daily challenges encountered by researchers in LMICs. Data are shown as mean ± SD, n = 3. Abbreviations are: NTC, non-template control; CFPS, cell-free protein synthesis; Min, minutes.

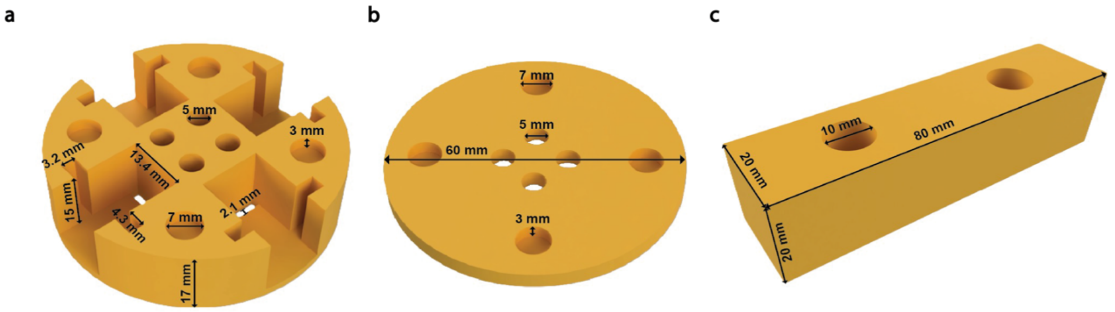

**Fig. S3: 3D-hand powered centrifuge (3D-fuge).** The main piece **(a)** and the cap **(b)** of the 3D-fuge are held together using eight magnets (four on each piece), which are glued into holes with a 7 mm diameter and 3 mm depth. All dimensions of each piece and the handles **(c)** are the same as those reported previously(*12*), except for the holes added for the magnets.

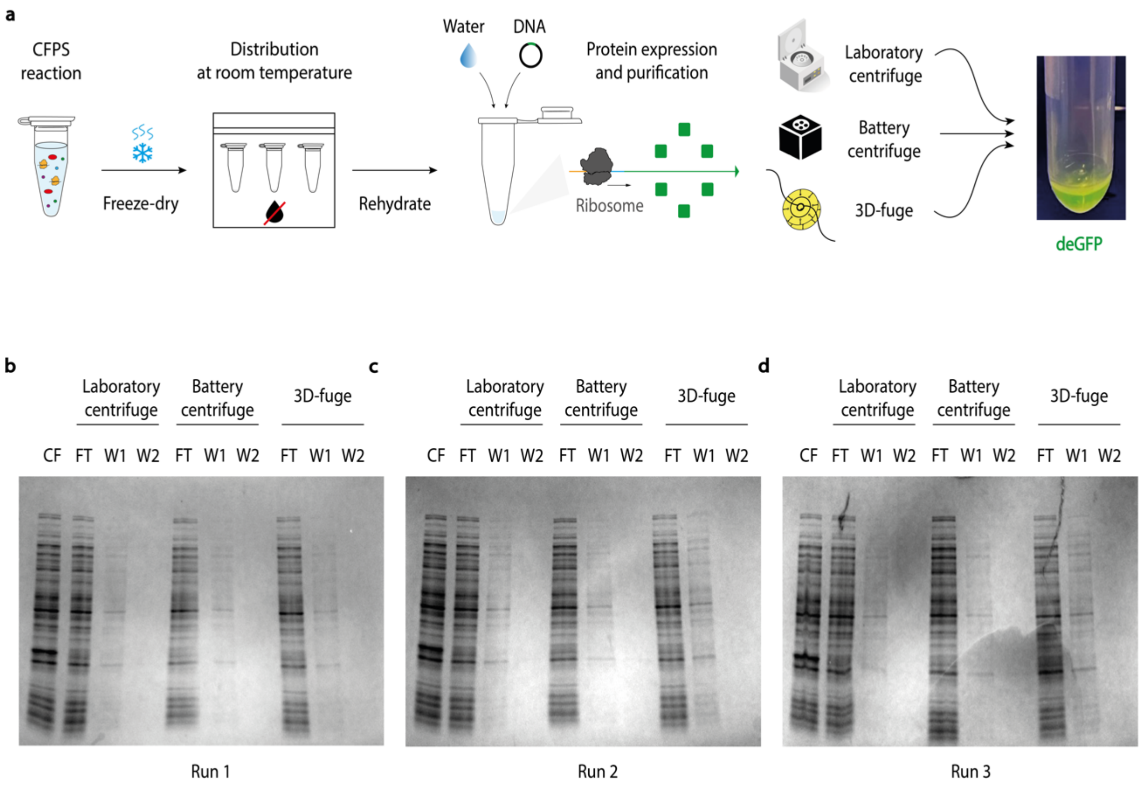

**Fig. S4: Comparable deGFP purification outcomes across three centrifugation systems. (a)** Schematic representation of the CFPS reaction setup and lyophilization process. Different centrifuges were used for deGFP purification, including a benchtop laboratory centrifuge, a battery-powered centrifuge, and our 3D-fuge. CFPS reactions were freeze-dried and distributed at ambient temperature. After one week, FD-CFPS reaction mixtures were rehydrated with water. This includes three independent experiments conducted on three different days, referred to here as run 1, run 2, and run 3. **(b-d)** All purification fractions (see elution fractions in Fig. 2f), including flow-through and wash fractions from the three purification methods, were analyzed by 4–20% gradient SDS-PAGE and stained using ProBlue Safe. These results confirm that protein purification was consistent across all centrifugation systems. This representative data was obtained using deGFP produced on-site in Colombia. Abbreviations are: CFPS, cell-free protein synthesis; CF, crude reaction; FT, flow-through; W1-2, washes.

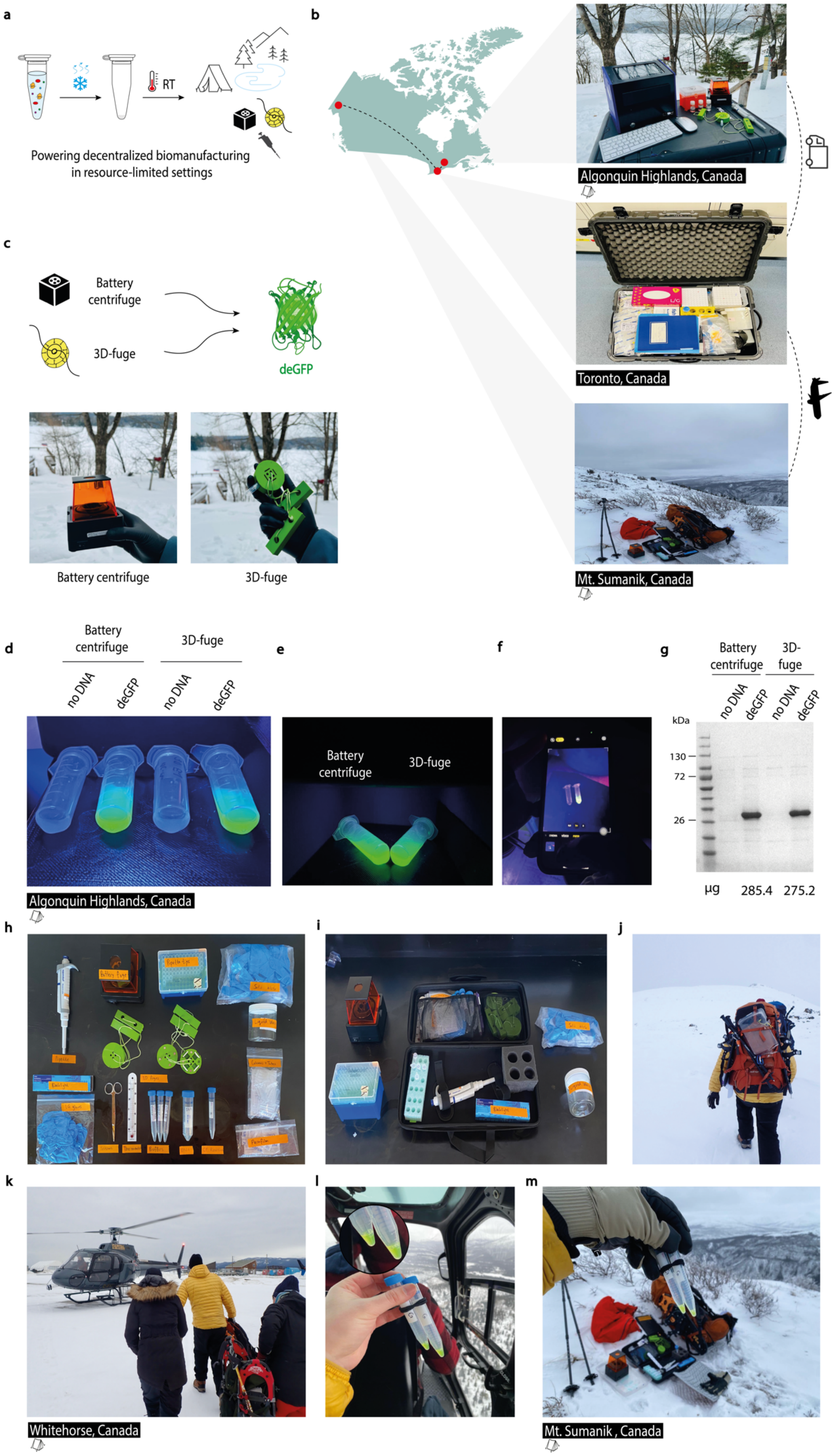
a
Powering decentralized biomanufacturing in resource-limited settings
RT
b
Algonquin Highlands, Canada
Toronto, Canada
Mt. Sumanik, Canada
c
Battery centrifuge
3D-fuge
deGFP
Battery centrifuge
3D-fuge
d
Battery centrifuge
3D-fuge
no DNA
deGFP
no DNA
deGFP
Algonquin Highlands, Canada
e
Battery centrifuge
3D-fuge
f
g
Battery centrifuge
3D-fuge
no DNA
deGFP
no DNA
deGFP
kDa
130
72
26
µg
285.4
275.2
h
i
j
k
Whitehorse, Canada
l
m
Mt. Sumanik , Canada

**Fig. S5: Portable FD-CFPS and low-burden centrifugation systems supported local protein manufacturing in resource-limited settings. (a)** After establishing the concept of cell-free biomanufacturing in controlled laboratory environments, we aimed to challenge traditional protein production models by demonstrating the system's capability to enable on-site protein production in remote settings, without reliance on conventional laboratory infrastructure. **(b)** FD-CFPS reactions, prepared in Toronto, Canada, were transported 250 km north to a remote site in Algonquin Highlands (Ontario), Mount Sumanik (Yukon), and Whitehorse (Yukon) in Northern Canada—all selected to simulate remote conditions settings. **(c)** Alongside the CFPS reactions, low-cost portable centrifugal devices (battery centrifuge and 3D-fuge) were deployed to enable local protein synthesis, as shown here with deGFP expression using minimal infrastructure. **(d-f)** Once on-site, FD-CFPS reactions (1 mL) were first used to produce deGFP (overnight at ambient temperature). Reactions were then divided into two equal parts for deGFP purification using Ni-NTA resin columns with two different centrifugation approaches. As a quality control step, reactions without the GFP-encoding plasmid were also used as negative controls in these experiments. According to our expectations, purified deGFP displayed a vivid green colour for both centrifugation strategies, while no fluorescence signal was observed in the control reactions. Elution fractions were illuminated using a standard UV flashlight, and images were captured with a smartphone camera. **(g)** Following field testing, eluates were brought back to the laboratory, and the results observed in the field were later confirmed through SDS-PAGE analysis and protein quantification assays. The purified deGFP was analyzed using 4–20% gradient SDS-PAGE and stained using ProBlue Safe. The molecular weight ladder (in kilodaltons) is shown on the left. **(h-m**) To illustrate mobile deployment in extreme conditions, deGFP expression and purification were also performed in challenging environments during a helicopter flight and a hike to Sumanik Ridge (Yukon) in Northern Canada. Equipped solely with essential supplies, all conveniently stored in a backpack, FD-CFPS reactions were rehydrated with template DNA encoding $His_6$-tagged deGFP, then incubated at ambient temperature (22–24 °C) and purified using a 3D-printed hand-powered device centrifuge. Purified deGFP displayed a vivid green colour. These findings show that producing high-yield, high-purity proteins locally in resource-limited settings is possible with low-burden FD-CFPS and simple, affordable tools. Abbreviations include: RT, room temperature.

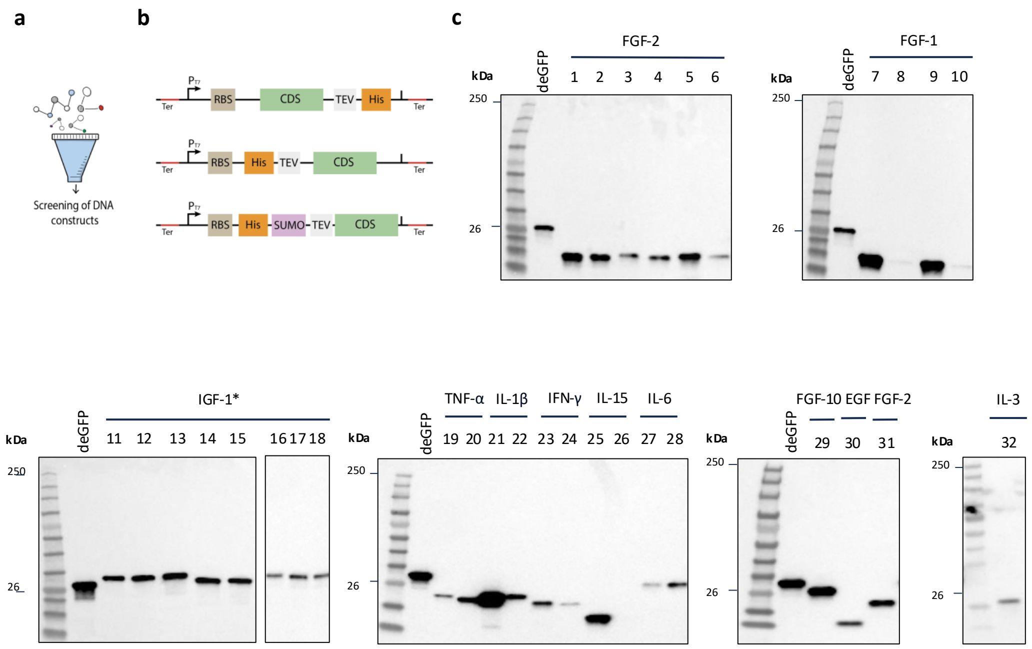

**Fig. S6: Screen of a panel of growth factors in E. coli cell-free lysates. (a,b)** A panel of DNA linear templates encoding 11 high-value growth factors was screened for expression in *E. coli* cell-free extracts (Supplementary Table S2, Supplementary Data 1). An N- or C-terminal $His_6$ was detected by Western blot analysis (see Methods for more details). A SUMO protein was also included in selected constructs (e.g., IGF-1) to enhance protein solubility when needed(*13*). **(c)** All growth factors in the DNA library were evaluated via Western blot. Consistent with our expectations, the position of the $His_6$-tag (N- or C-terminal) significantly impacted protein expression, leading to reduced or even complete loss of expression. The molecular weight ladder (in kilodaltons) is shown on the left. Images are representative of at least three independent experiments.

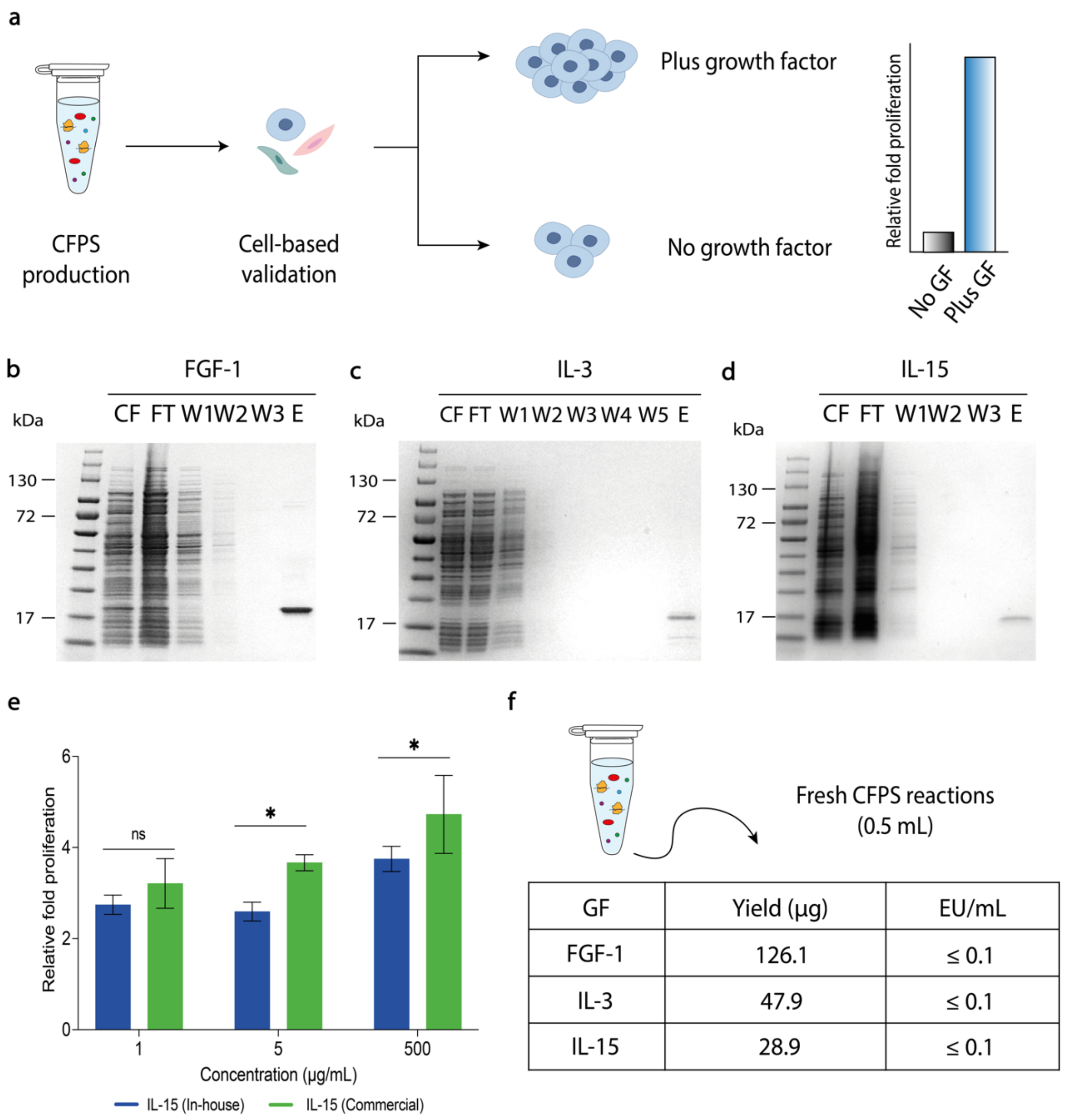

**Fig S7: CFPS system enables decentralized and scalable production of functional growth factors. (a)** Schematic representation of the growth factor production pipeline, ultimately applied to assess cell proliferation in cell culture-based assays. Fresh CFPS reactions were used to produce small batches of high-value growth factors, which were subsequently validated for their ability to induce proliferation in appropriate cell lines. **(b-d)** FGF-1, IL-3, and IL-15 were produced in CFPS reactions. Purified growth factors were analyzed using 4–20% gradient SDS-PAGE and stained using ProBlue Safe, yielding products of the expected size. The molecular weight ladder (in kilodaltons) is shown on the left. **(e)** *In vitro* proliferation assay comparing on-demand, locally produced (blue) and commercial (green) IL-15 growth factors in T cells. Cells were individually treated with varying concentrations (1, 5, and 500 μg/mL). Luminescence was read using a conventional plate reader. This representative data was obtained using reagents produced on-site in Canada. Data are shown as mean ± SD, n = 3. **(f)** Protein yield from 0.5 mL CFPS reaction and endotoxin assays confirmed compliance with standard endotoxin limits (≤ 0.1 EU/mL), with purity > 90%. Statistical differences were determined by two-way ANOVA with Šídák's post hoc multiple comparisons test: ns $p > 0.05$, * $p < 0.05$. Abbreviations are: ns, not significantly different; CFPS, cell-free protein synthesis; EU, endotoxin units; GF, growth factor; CF, crude reaction; FT, flow-through; W1-5, washes; E, elution.

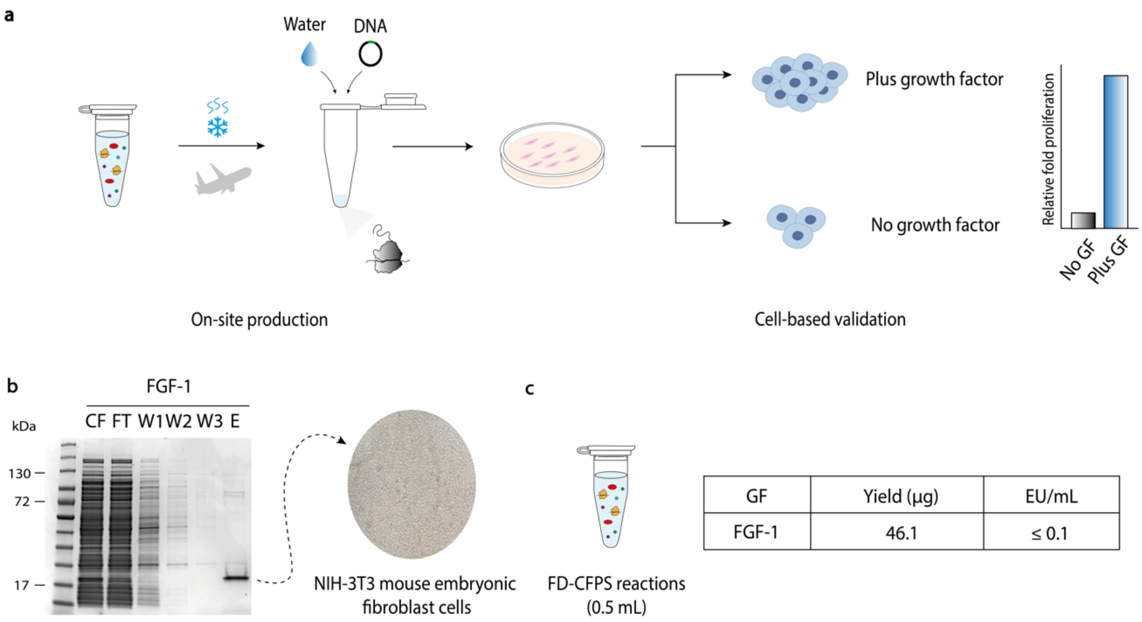

**Fig. S8: CFPS offers a robust platform for synthesizing biologically active growth factors in resource-limited settings. (a)** Schematic pipeline representation for growth factor production, which is ultimately used for cell-based proliferation assays in resource-limited settings. CFPS reactions (prepared in Toronto, Canada) were lyophilized and transported to Recife, Brazil, at ambient temperature. **(b,c)** On-site, FD-CFPS reactions (0.5 mL) were used to produce FGF-1, which was subsequently validated for its ability to induce proliferation in NIH-3T3 mouse embryonic fibroblast cells. The purified FGF-1 was subsequently analyzed using a 4–20% gradient SDS-PAGE and stained with ProBlue Safe, yielding a product with low endotoxin levels (≤ 0.1 EU/mL) and high purity (>90%). These findings confirm the ability to produce high-value growth factors locally using low-burden CFPS reactions and minimal laboratory infrastructure. The molecular weight ladder (in kilodaltons) is shown on the left. Abbreviations are: GF, growth factor; EU, endotoxin units; FD, freeze-dried; CFPS, cell-free protein synthesis; CF, crude reaction; FT, flow-through; W1-3, washes; E, elution.

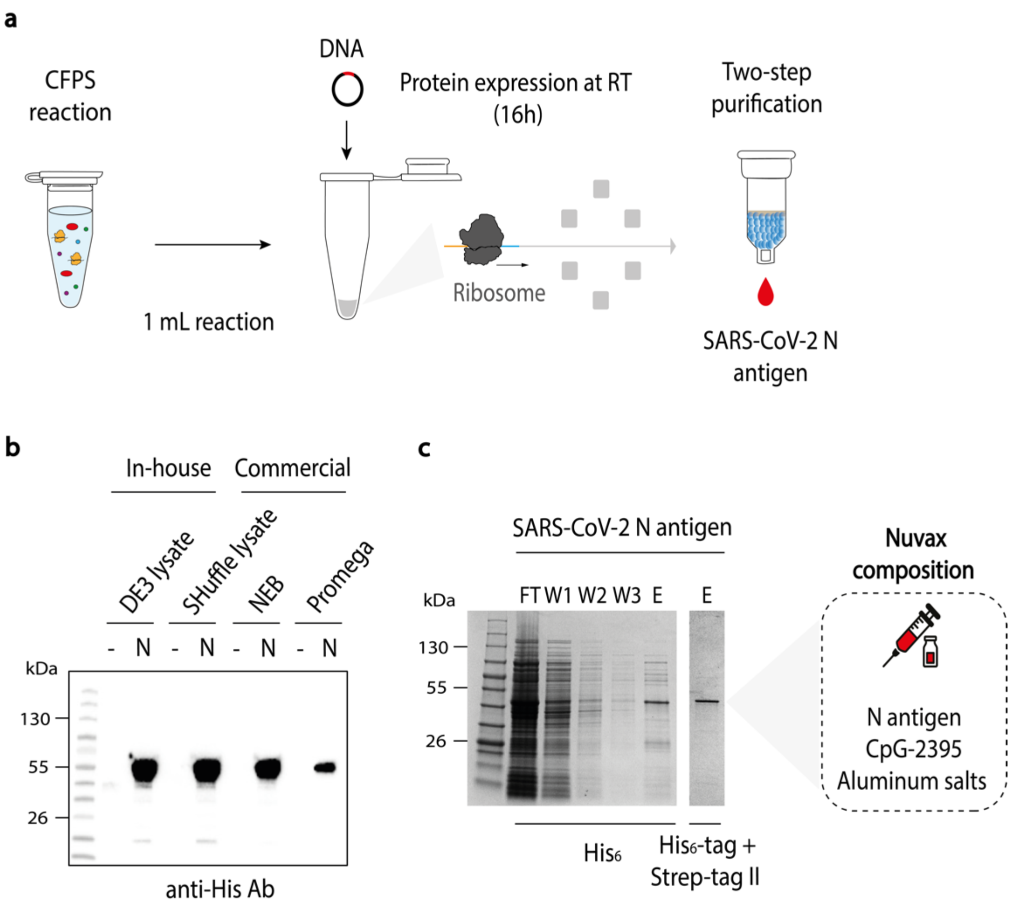

**Fig. S9: CFPS platform enabled local and portable production of vaccine candidates. (a)** Schematic representation of a one-day, cost-efficient pipeline for vaccine production. **(b)** SARS-CoV-2 nucleocapsid antigen was first expressed in small-scale reactions (10 µL, 24 °C, 16 hours), with expression confirmed by Western blot analysis using an anti-His Antibody (Ab). Here, with the top-performing DNA construct identified, the SARS-CoV-2 N antigen was successfully expressed using in-house, lysate-based CFPS from BL21(DE3) or SHuffle *E. coli* strains, as well as in commercially available *E. coli* lysates from NEB and Promega. The molecular weight ladder (in kilodaltons) is shown on the left. **(c)** To obtain a high-purity antigen suitable for therapeutic use, we combined two-step spin column purification (Ni-NTA ($His_6$-tag) and Strep-Tactin$^{TM}$ (Strep II tag)). This strategy yielded a high-purity product (>90%). The molecular weight ladder is shown on the left. Images are representative of at least three independent experiments. With the protocol for antigen production in place, we created a COVID-19 vaccine formulation containing the SARS-CoV-2 N antigen, CpG-2395, and Aluminum salts, which was designated as Nuvax. After production, the immunogenicity of the vaccine was assessed in a murine model. To our knowledge, this is the first COVID-19 subunit-based vaccine produced in CFPS systems. Abbreviations are: N, nucleocapsid; CFPS, cell-free protein synthesis; RT, room temperature; FT, flow-through; W1-3, washes; E, elution.

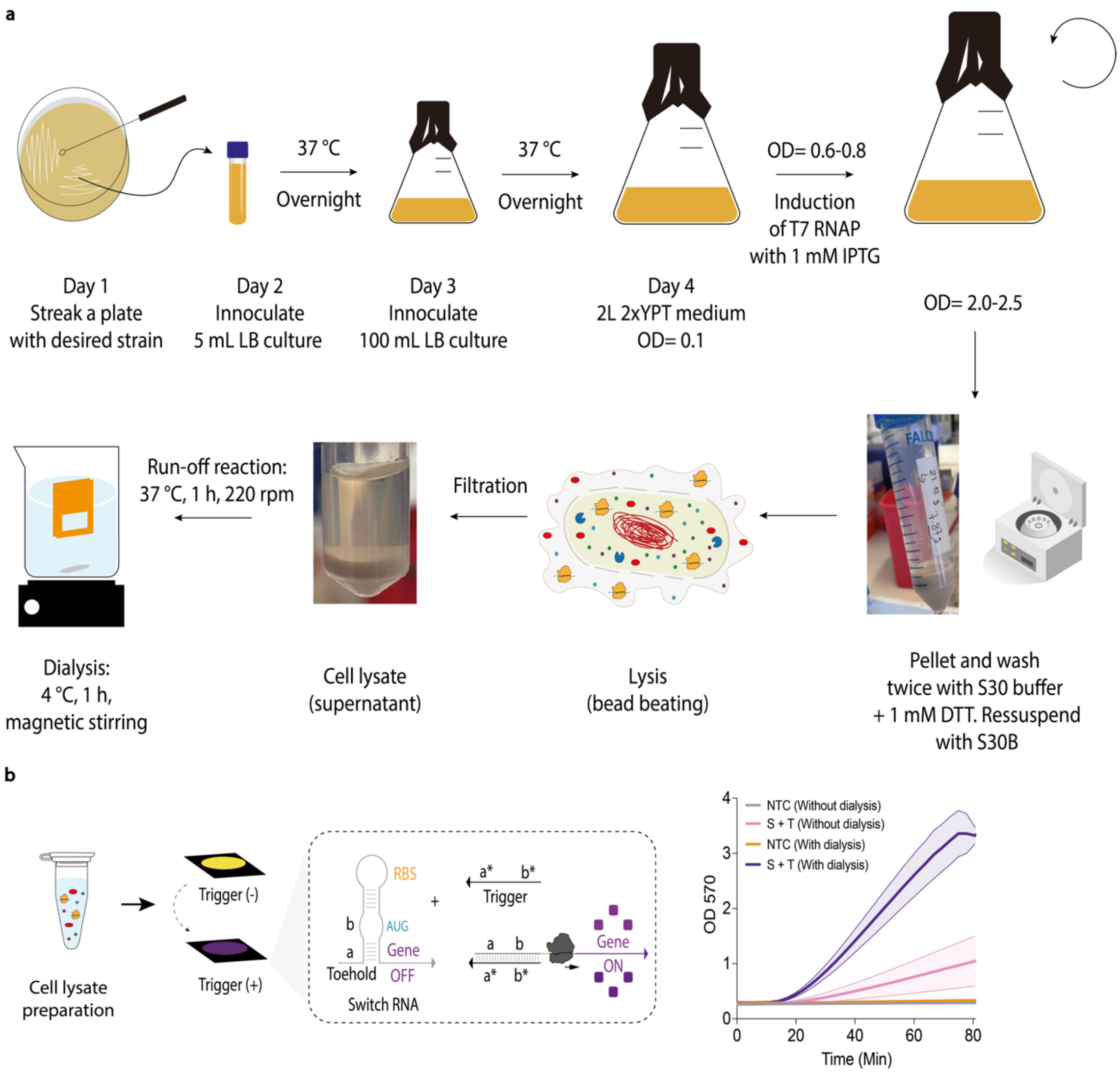

**Fig. S10: A low-cost, modular, scalable workflow for cell lysate preparation.** (a) Schematic representation of a modular and scalable workflow for the local production of cell lysate. The workflow for cell lysate preparation can be carried out in basic low-containment microbiology laboratories. Here, this protocol was specifically employed to produce low-cost cell lysates designed to operate with toehold switch-based sensors. A typical 2 L culture yields approximately 5 mL of cell lysate, which can be either frozen for later on-site use or freeze-dried for shipment and distribution at ambient temperature to any location worldwide. **(b)** Initially, two different cell lysates were produced. One batch was prepared without an additional post-lysis step, while the second included an additional dialysis step. The results demonstrated that the toehold switch-based sensors performed better when using the lysate produced with the additional dialysis step. Lysate performance, measured by β-galactosidase activity, was assessed by measuring absorbance at 570 nm over 80 minutes in a conventional plate reader. In this proof-of-concept experiment, the sensors were specifically designed for Zika virus detection and have been previously characterized in our previous studies(*2, 10, 14, 15*). With the protocol established for cell lysate preparation, CFPS reactions (prepared in Santiago, Chile) were shipped to research team members in Canada, Brazil, Colombia, and India. Data are shown as mean ± SD, n = 6. Abbreviations are: NTC, non-template control; S, switch; T, trigger; T7 RNAP, T7 RNA polymerase.

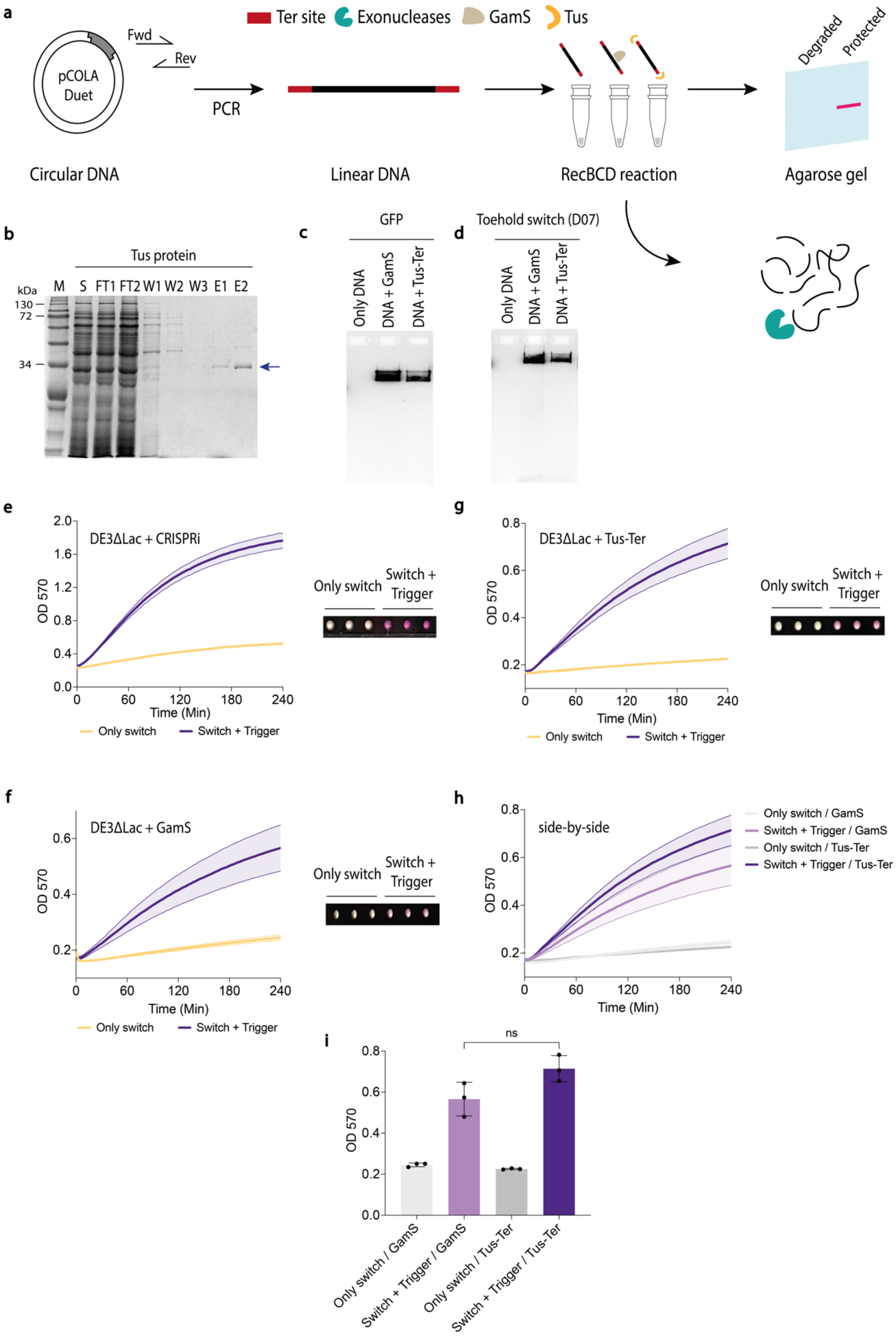

**Fig. S11: Tus-Ter-protected linear DNA enables functional toehold switch sensors in crude lysate. (a)** Schematic representation of Tus-Ter-mediated protection of linear DNA templates. Starting with circular DNA (plasmid containing the D07 toehold switch), PCR is used to produce linear DNA while simultaneously incorporating Ter sites through the Forward (Fwd) and Reverse (Rev) primers. When the Ter sites are present, the Tus protein binds specifically to them, forming the Tus-Ter complex. This protects the linear DNA ends from exonuclease degradation(*4*). On the other hand, in the absence of Ter sites, the linear DNA remains vulnerable and is rapidly degraded by the exonucleases present in the crude lysate. Here, we benchmarked the performance of our protection system against GamS (NEB, P0774S), a commercially available reagent widely used to inhibit nuclease activity in CFPS reactions. **(b)** The Tus protein was expressed in *E. coli* BL21 (DE3)-Gold-ΔLac cells and purified as reported previously(*4*). Purified product was then analyzed using SDS-PAGE and stained using ProBlue Safe, yielding a product of the expected size. The molecular weight ladder (in kilodaltons) is shown on the left. **(c,d)** We first tested the system in the presence of a commercial *E. coli* RecBCD complex (NEB, M0345S), which exhibits both endonuclease and exonuclease activity. In this proof-of-concept experiment, both strategies effectively protected linear DNA templates with similar performance, while unprotected DNA templates were rapidly degraded by nucleases (indicated as no bands). Agarose gel electrophoresis (2%) confirmed the presence of both the GFP (control) and the toehold switch (D07) DNA corresponding bands. **(e)** Having established the system in vitro, we advanced to test the system in crude lysate. Activation of the D07 toehold switch was observed in fresh lysate prepared from an *E. coli* BL21(DE3) Star strain engineered with CRISPRi (*E. coli* BL21(DE3) Star/CRISPRi+) to improve linear DNA stability in crude lysates(*15*). ssDNA trigger (4 µM) was added to D07 toehold switch-containing lysate reactions, and β-galactosidase activity was measured by absorbance at 570 nm for 4 h. Data are shown as mean ± SD, n = 3. **(f,g)** Similarly, activation of the D07 toehold switch was successfully observed in fresh lysates prepared from an *E. coli* BL21 (DE3)-Gold-ΔLac strain supplemented individually with either GamS or (NEB) or Tus protein. ssDNA trigger (4 µM) was added to D07 toehold switch-containing lysate reactions, and β-galactosidase activity was measured by absorbance at 570 nm for 4 h. Data are shown as mean ± SD, n = 3. **(h,i)** The graphs present side-by-side comparisons of 4-hour absorbance kinetics at 570 nm and end-point measurements for both protection strategies, showing comparable performance. Following incubation, reactions exhibited a visible colour change, enabling detection and interpretation by the naked eye without the need for a specialized instrument. A positive reaction resulted in a colour change from yellow to purple, while a negative reaction remained yellow. Colorimetric results are displayed on the right side of the corresponding graphs. These findings underscore the Tus-Ter as a promising mechanism for protecting toehold switch-based linear DNA inputs in crude lysates. Data are shown as mean ± SD, n = 3. An unpaired two-tailed Student's t-test was performed. Abbreviations are: ns, not significantly different; Min, minutes; S, supernatant; FT, flow-through; W1-3, washes; E, elution.

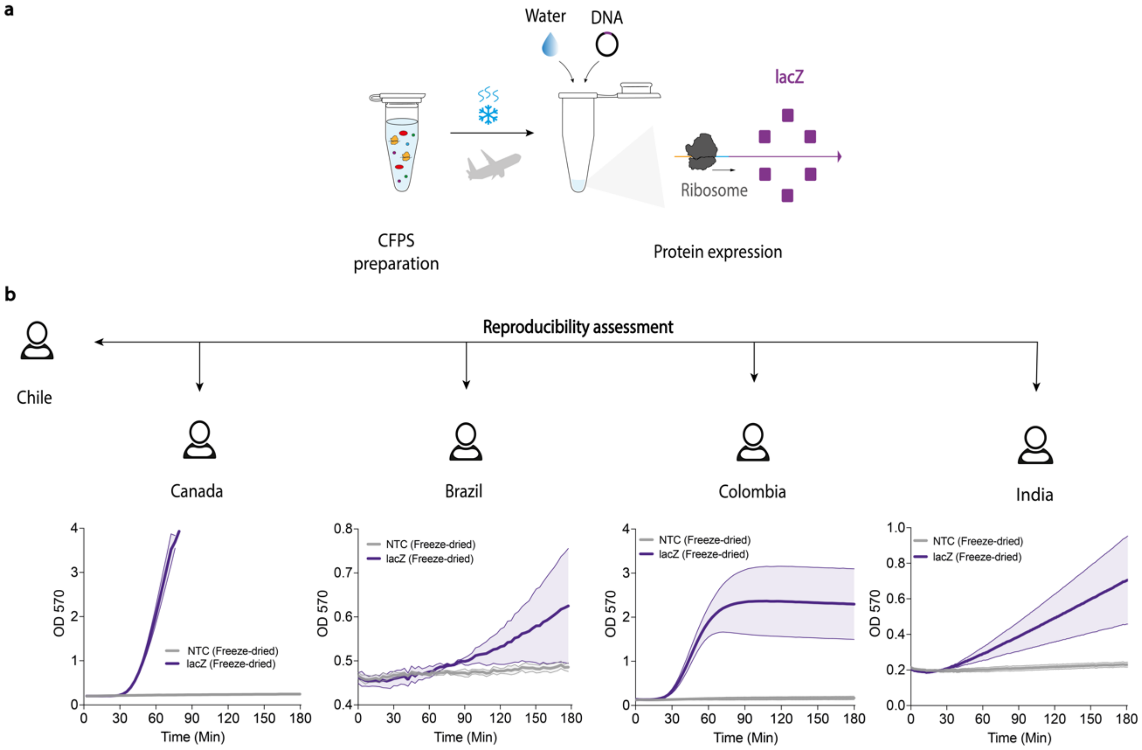

**Fig. S12: Portable FD-CFPS systems enabled local protein manufacturing across diverse settings. (a)** Schematic representation of the CFPS reaction setup, lyophilization procedure, and reproducibility conducted across collaborating teams. CFPS reactions (prepared in Santiago, Chile) were lyophilized and distributed at ambient temperature using conventional logistics couriers (e.g., FedEx) to multiple international sites, including Canada, Brazil, Colombia, and Chile. **(b)** Here, FD-CFPS activity was assessed using a *lacZ* expression construct by measuring absorbance at 570 nm over 3 h in a conventional plate reader. While successful in Chile, Canada, Colombia, and India, test performance was inconsistent in Brazil, where shipping and customs delays impacted cell-free lysate activity, underscoring the everyday challenges experienced by researchers in LMICs. Data are shown as mean ± SD, n = 3. Abbreviations are: NTC, non-template control; Min, minutes; CFPS, cell-free protein synthesis.

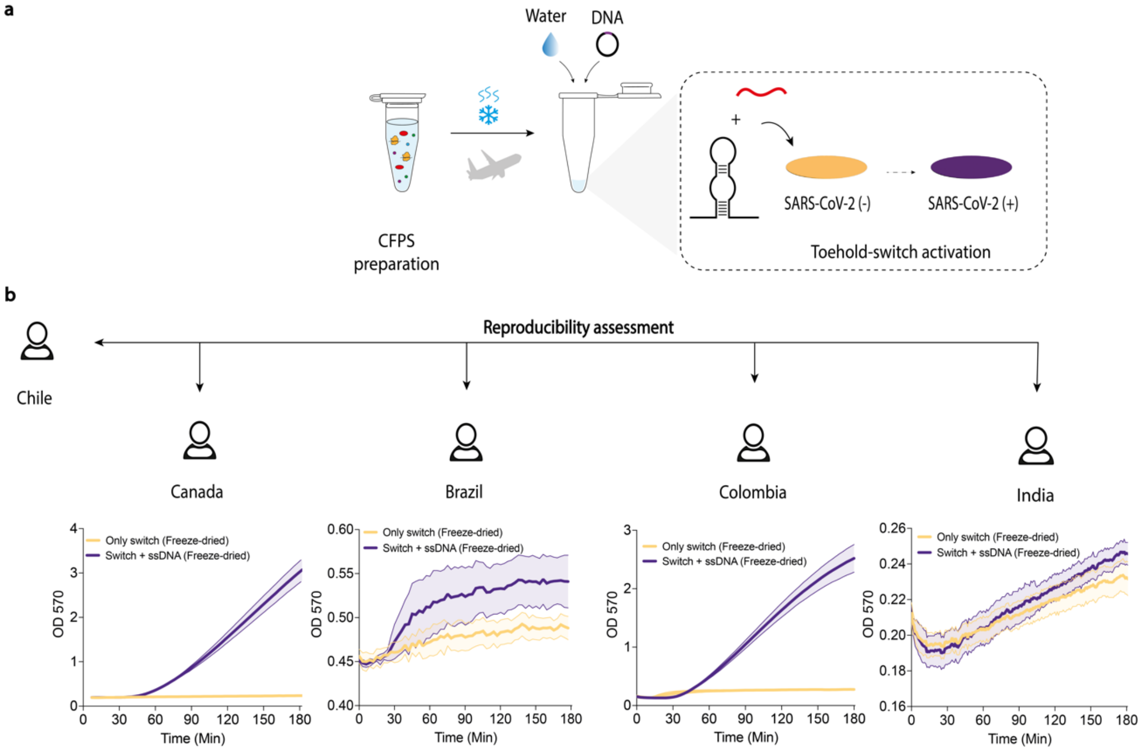

**Fig. S13: Portable FD-CFPS systems enabled global distribution of toehold switch-based diagnostics. (a)** Schematic representation of the CFPS reaction setup, lyophilization process, and reproducibility assessment across collaborating teams. FD-CFPS reactions and synthetic controls (prepared in Chile and Canada) were distributed at ambient temperature using conventional logistics couriers (e.g., FedEx) to multiple international sites. **(b)** Once on-site, activation of the D07 toehold switch was assessed in freeze-dried lysates at each site. ssDNA trigger was added to D07 toehold switch-containing lysate reactions, and β-galactosidase activity was measured by absorbance at 570 nm for 3 h of reaction incubation. While successful in Chile, Canada, and Colombia, test performance was inconsistent in Brazil and India due to shipping-related lysate degradation, highlighting the daily challenges encountered by researchers in LMICs. Data are shown as mean ± SD, n = 3. Abbreviations are: NTC, non-template control; Min, minutes; CFPS, cell-free protein synthesis.

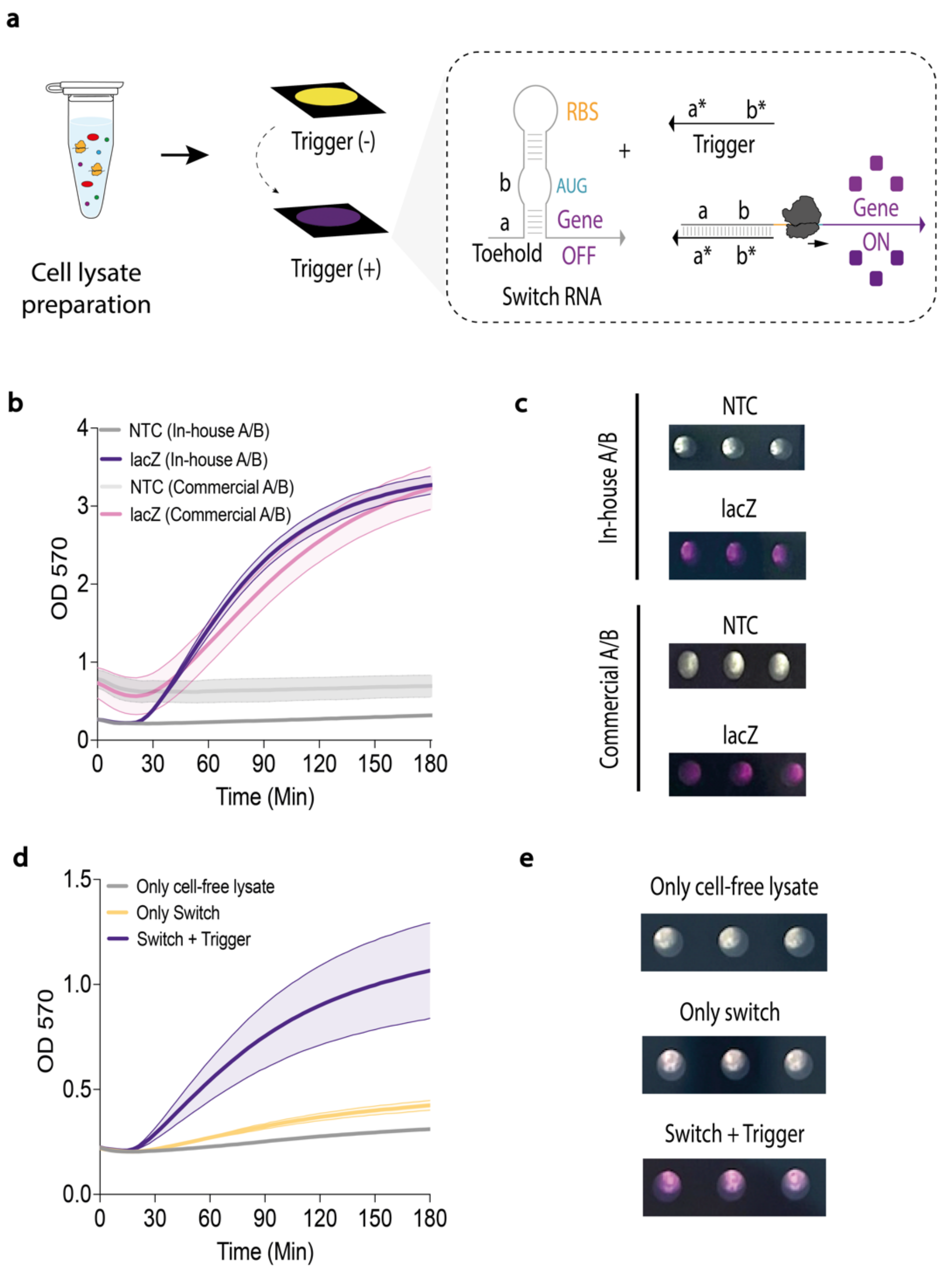

**Fig. S14: Standardized, streamlined protocols enabled the rapid implementation of lysate preparation and toehold switch-based diagnostics across diverse settings. (a)** Schematic representation of the toehold switch-based reaction mechanism. Using standardized protocols, we successfully established local production of cell lysate and toehold switch diagnostics at sites across South and North America. **(b,c)** With the protocol for lysate preparation and optimized reaction supplements (referred to as Solution A and Solution B) in hand, side-by-side benchmarking was carried out with commercially available cell-free reagent supplement solutions, providing comparable results performance. Lysate performance, measured by β-galactosidase activity, was assessed by measuring absorbance at 570 nm over 1 h. This representative data was obtained using cell-free lysates produced on-site in Canada. Colourimetric results are displayed on the right side of the corresponding graphs. A positive reaction resulted in a colour change from yellow to purple, while a negative reaction remained yellow. Data are shown as mean ± SD, n = 3. **(d,e)** With the system established in place, an additional experiment demonstrated the activation of the toehold switch-based diagnostics.

ssDNA trigger (4 µM) was added to D07 toehold switch-containing lysate reactions, and β-galactosidase activity was measured by absorbance at 570 nm for 3 h. Colourimetric results are displayed on the right side of the corresponding graphs. Representative data was obtained using reagents produced on-site in Canada. Data are shown as mean ± SD, n = 3. Abbreviations are: NTC, non-template control; Min, minutes.

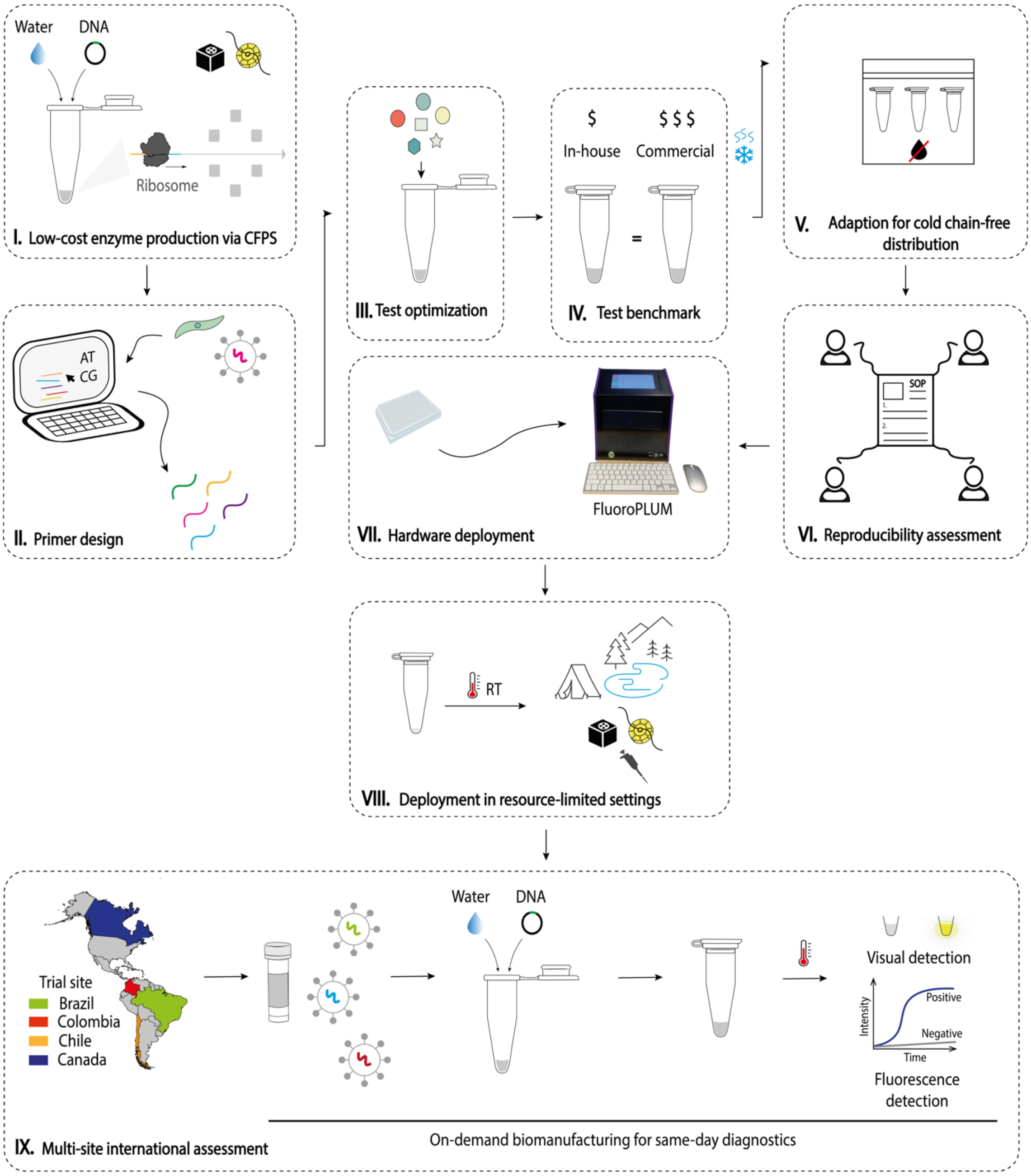

**Fig. S15: On-demand biomanufacturing for same-day diagnostics.** Pipeline illustrating all steps (I-IX) used to build a streamlined framework for local production of diagnostic reagents within a single day, enabling the establishment of disease diagnostic programs across four countries and on-site testing. This included the following steps: I. Low-cost enzyme production via CFPS; II. Primer design; III. Test optimization; IV. Test benchmark; V. Adaptation for cold chain-free distribution; VI. Reproducibility assessment; VII. Hardware deployment; VIII. Deployment in resource-limited settings; and IX. Multi-site international assessment in Brazil, Colombia, Chile, and Canada. Abbreviations are: CFPS, cell-free protein synthesis; RT, room temperature.

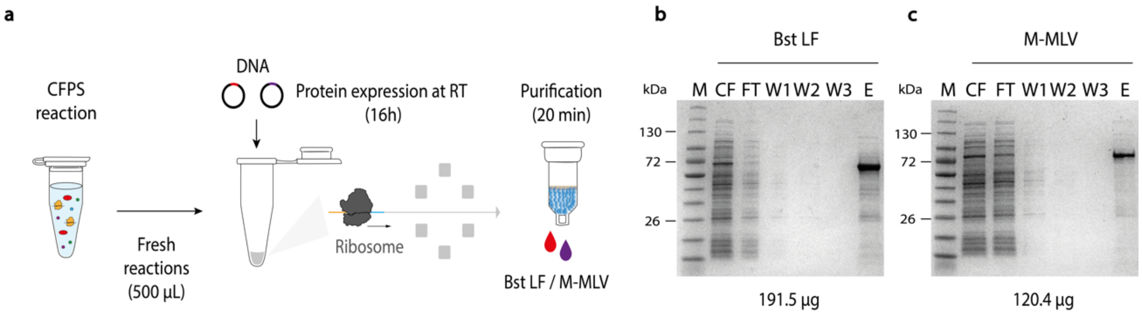

**Fig. S16: Low-burden CFPS reactions enabled decentralized, same-day biomanufacturing of essential diagnostic enzymes. (a)** Schematic representation of the CFPS reaction setup and protein purification process. Fresh CFPS reactions were utilized to produce batches of high-value diagnostic enzymes, which were subsequently employed to develop an in-house LAMP/RT-LAMP diagnostic workflow for detecting clinically relevant pathogens. Bst LF and M-MLV were produced in CFPS reactions (500 µL reactions, at ambient temperature). After overnight expression, the enzymes were purified using centrifugation-based affinity chromatography with hexa-histidine [$His_6$], which can be performed using readily available resin-packed microcentrifuge columns (a 20-minute protocol). This enabled the development of a rapid and straightforward framework for producing diagnostic enzymes locally within a single day, which was later implemented across multiple locations worldwide. **(b-c)** Purified diagnostic enzymes were analyzed using 4–20% gradient SDS-PAGE and stained using ProBlue Safe, yielding products of the expected size. The molecular weight ladder (in kilodaltons) is shown on the left. Protein quantification was performed using the Pierce BCA protein assay kit, and the resulting quantification values are presented below the gels. Representative gel results were achieved using diagnostic enzymes produced on-site in Canada, laying the groundwork for later implementation by international collaborators across diverse settings, ranging from well-resourced labs to environments with minimal infrastructure. Abbreviations are: CFPS, cell-free protein synthesis; RT, room temperature; CF, crude reaction; FT, flow-through; W1-3, washes; E, elution.

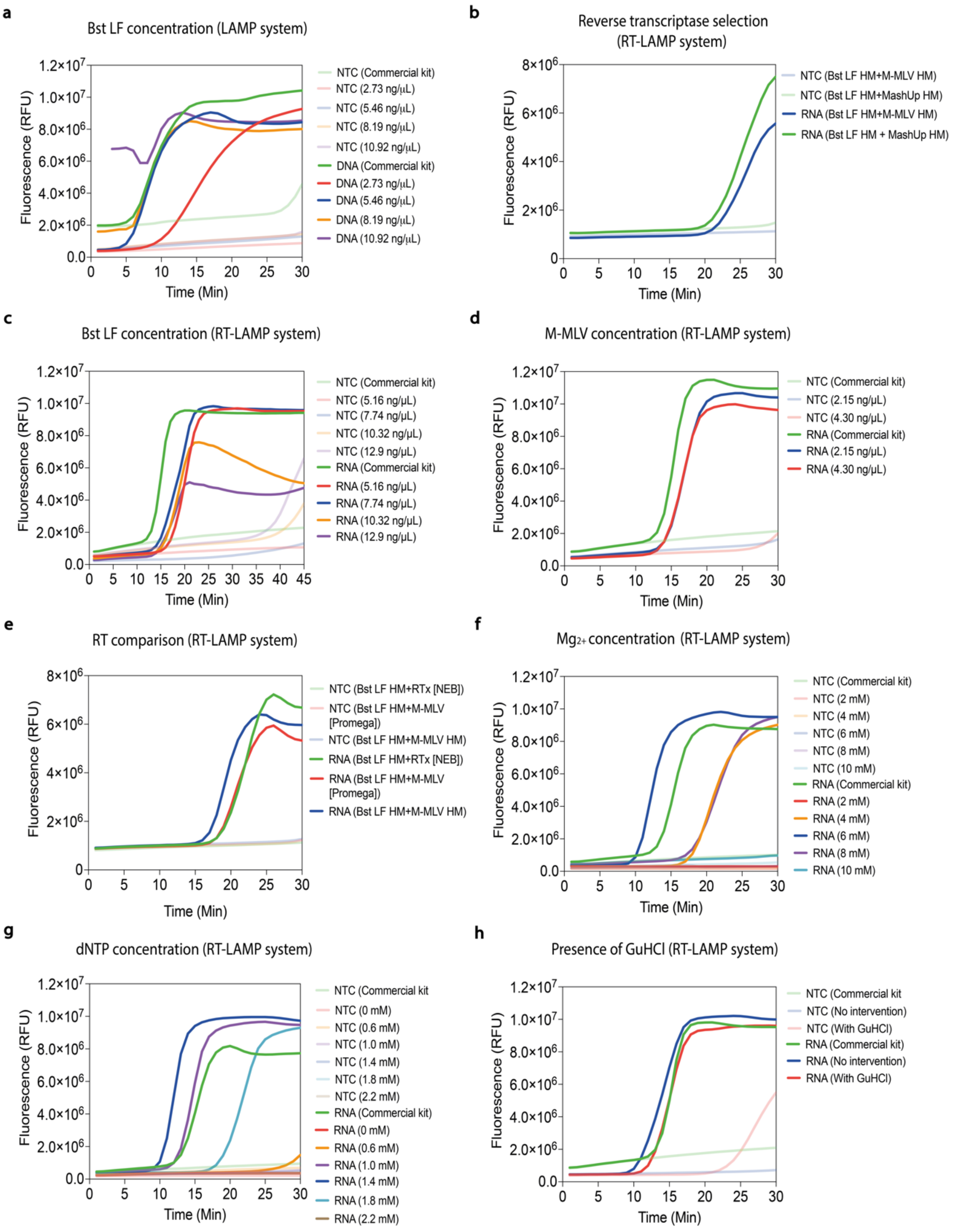

**Fig. S17: Screening for optimal conditions enabled the establishment of a reliable LAMP/RT-LAMP diagnostic platform for detecting DNA and RNA.** Molecular reactions were optimized to achieve performance comparable to commercial kits (WarmStart, NEB). To this end, multiple parameters and reaction conditions were systematically evaluated alongside a commercial kit for direct comparison (green data), including **(a)** Bst LF concentration for DNA detection, **(b)** different reverse transcriptase (RT) enzymes locally produced (M-MLV and MashUP), **(c)** Bst LF concentration for RNA detection, **(d)** M-MLV concentration for RNA detection, **(e)** direct comparison of our in-house RT enzyme against two enzymes from NEB and Promega, **(f)** $Mg^{2+}$ concentration, **(g)** dNTP concentration, and **(h)** the addition of guanidine hydrochloride (GuHCl). Real-time fluorescence experiments were conducted for the amplification of synthetic *P. falciparum* DNA (LAMP) or SARS-CoV-2 RNA (RT-LAMP) using a conventional qPCR instrument. After optimization, the optimal conditions for all parameters (blue data) were selected for further experiments. As a result of this effort, we have successfully developed and established a robust and universal diagnostic pipeline using locally manufactured inputs that match the performance of leading commercial kits. With the optimized system in place, the molecular assays were evaluated and benchmarked side by side against a commercial kit in Canada, Chile, Brazil, and Colombia. Abbreviations are: NTC, non-template control; Min, minutes; Mg, magnesium; RT, reverse transcriptase.

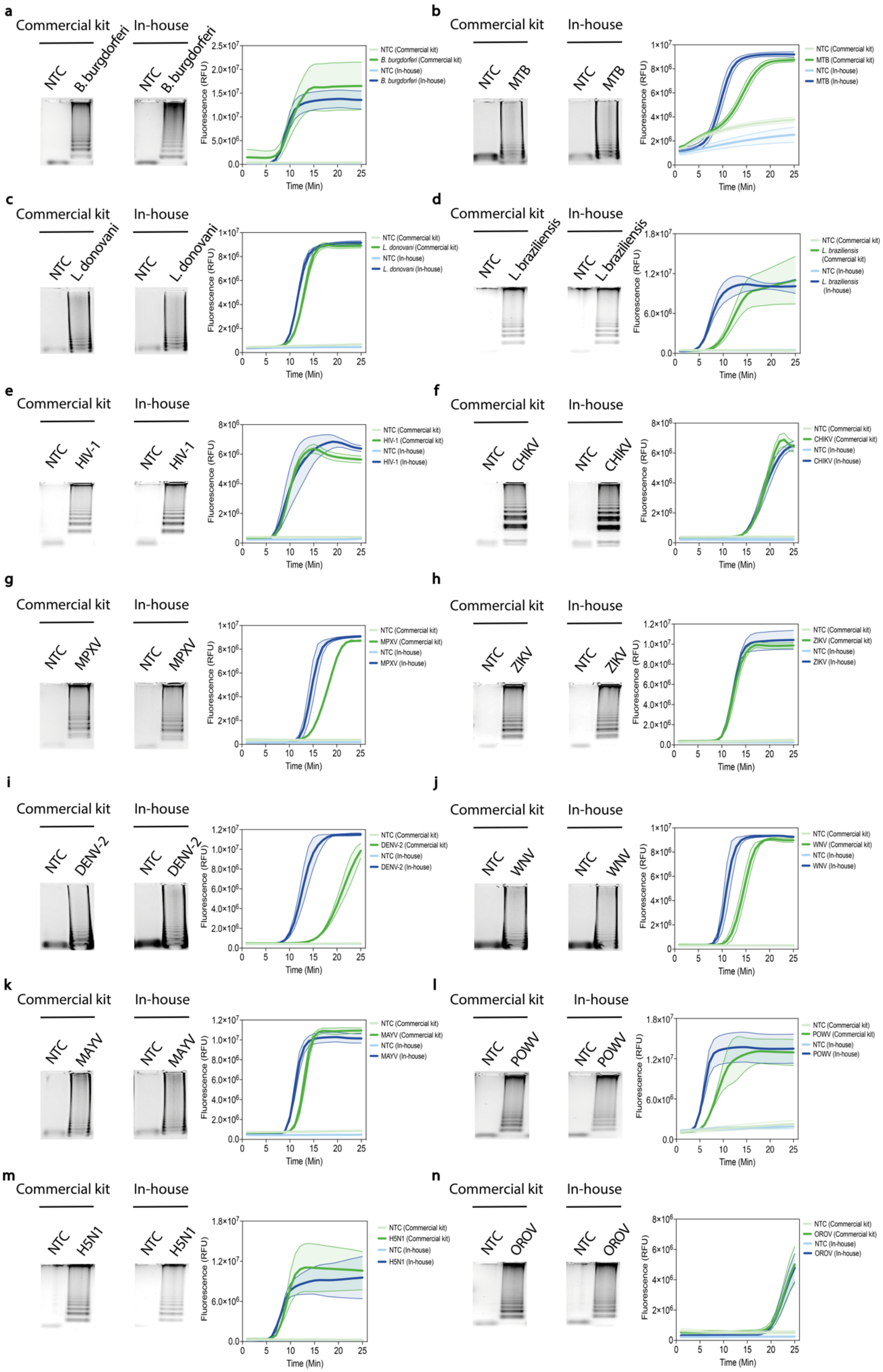

**Fig. S18: In-house LAMP/RT-LAMP reactions performed similarly or even better than commercial kits.** Having optimized the diagnostic platform for detecting DNA and RNA elements, we benchmarked the activity of our in-house LAMP/RT-LAMP reactions (blue data) against an available commercial kit (green data), with results showing equivalent performance for DNA and RNA targets across all pathogens sequences: **(a)** *Borrelia burgdorferi,* **(b)** *Mycobacterium tuberculosis,* **(c)** *Leishmania donovani,* **(d)** *Leishmania braziliensis,* **(e)** human immunodeficiency virus 1 (HIV-1), **(f)** chikungunya virus (CHIKV), **(g)** monkeypox virus (MPXV), **(h)** Zika virus (ZIKV), **(i)** dengue virus serotype 2 (DENV-2), **(j)** West Nile virus (WNV), **(k)** Mayaro virus (MAYV), **(l)** Powassan virus (POWV), **(m)** avian influenza A (H5N1) virus, and **(n)** Oropouche virus (OROV). Real-time fluorescence measurements (25 minutes for all targets) were visualized using a conventional qPCR instrument with fluorescence reads every minute, comparing in-house and commercial LAMP systems. Data are shown as mean ± SD, n = 3. For visual detection, amplification products were visualized by naked eye under natural light **(see Fig. 6 b)**, and the amplicons were analyzed using agarose gel electrophoresis (1.5%). The agarose gels are shown on the left side of each panel. Abbreviations are: NTC, non-template control; Min, minutes; MTB, *Mycobacterium tuberculosis*.

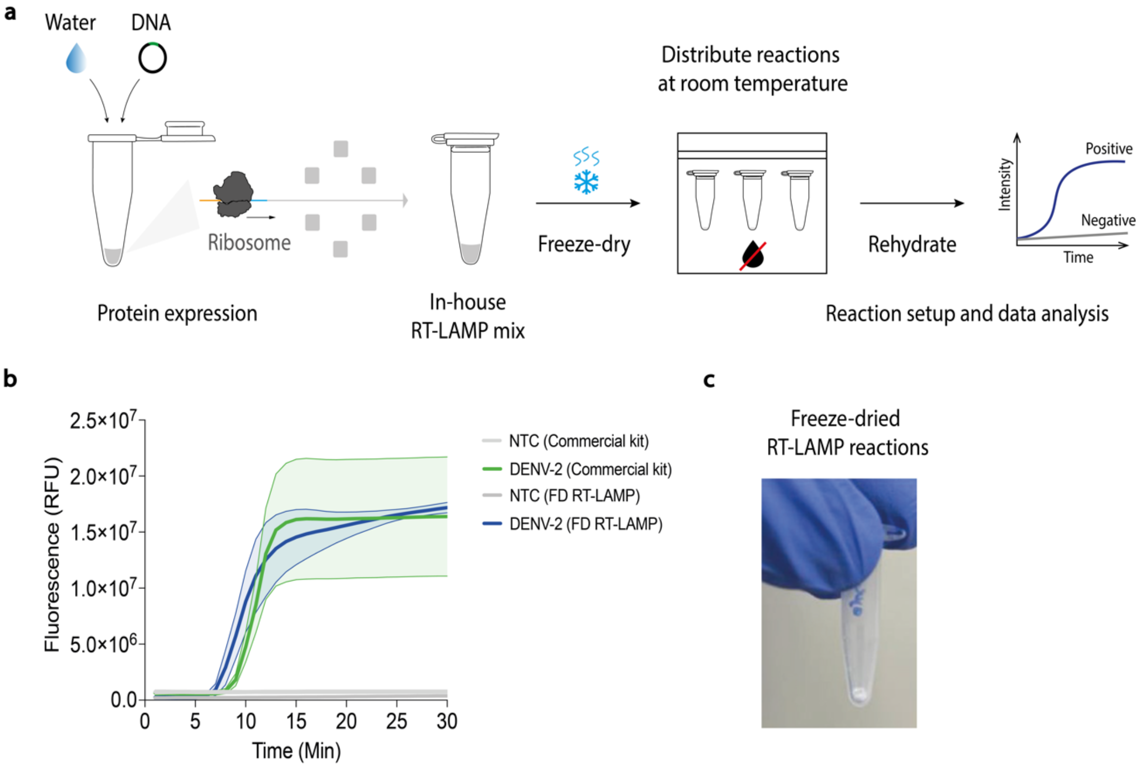

**Fig. S19: In-house RT-LAMP reactions are stable at ambient temperature and can be stored and distributed without cold chain logistics. (a)** Schematic representation illustrating the CFPS-based enzyme biosynthesis, RT-LAMP assay setup, lyophilization process, and fluorescence measurement. RT-LAMP reactions were freeze-dried and stored at ambient temperature (22–24 °C). After rehydrating with the appropriate buffer solution, we benchmarked the activity of our in-house RT-LAMP reactions (blue data) against an available commercial kit (green data) to detect DENV-2 RNA, with results showing similar performance. **(b)** Real-time fluorescence measurements were visualized using a conventional qPCR instrument with fluorescence reads every minute. Data are shown as mean ± SD, n = 3. **(c)** Representative photograph of a freeze-dried RT-LAMP reaction. Abbreviations are: NTC, non-template control; DENV-2, dengue virus, serotype 2; Min, minutes; FD, freeze-dried.

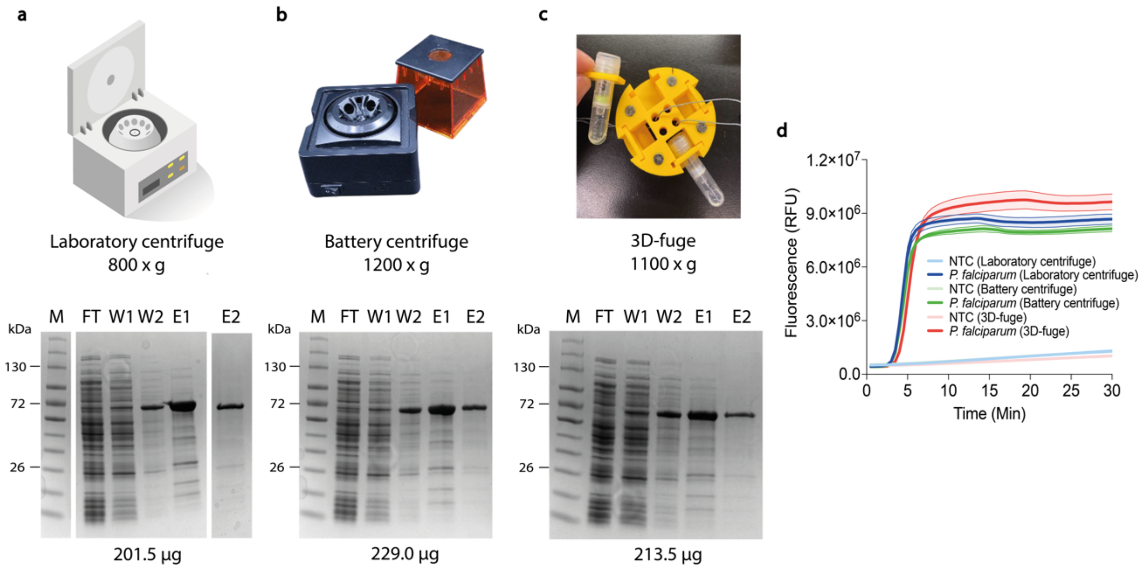

**Fig. S20: Local production of diagnostic enzymes is achievable with either standard laboratory infrastructure or minimal, low-burden tools. (a,b,c)** Different centrifugal devices were used for the affinity purification of Bst LF, which was later employed to build in-house LAMP reactions. These include a benchtop lab centrifuge, a battery-powered centrifuge, and our 3D-fuge **(see table S1 for more details)**. In brief, a fresh CFPS reaction (1 mL) was prepared to produce Bst LF. After overnight expression at ambient temperature, reactions were divided into three equal parts for protein purification with Ni-NTA resin columns. The purified Bst LF was subsequently analyzed using 4–20% gradient SDS-PAGE and stained using ProBlue Safe, yielding a product of the expected size. The molecular weight ladder (in kilodaltons) is shown on the left. Protein quantification was performed using the Pierce BCA protein assay kit, and the resulting quantification values are presented below the gels, which confirmed consistent protein yields across all purification strategies. **(d)** Real-time fluorescence measurements targeting *P. falciparum* synthetic DNA were visualized using a conventional qPCR instrument with fluorescence reads every minute. Data are shown as mean ± SD, n = 3. Abbreviations are: NTC, non-template control; Min, minutes.

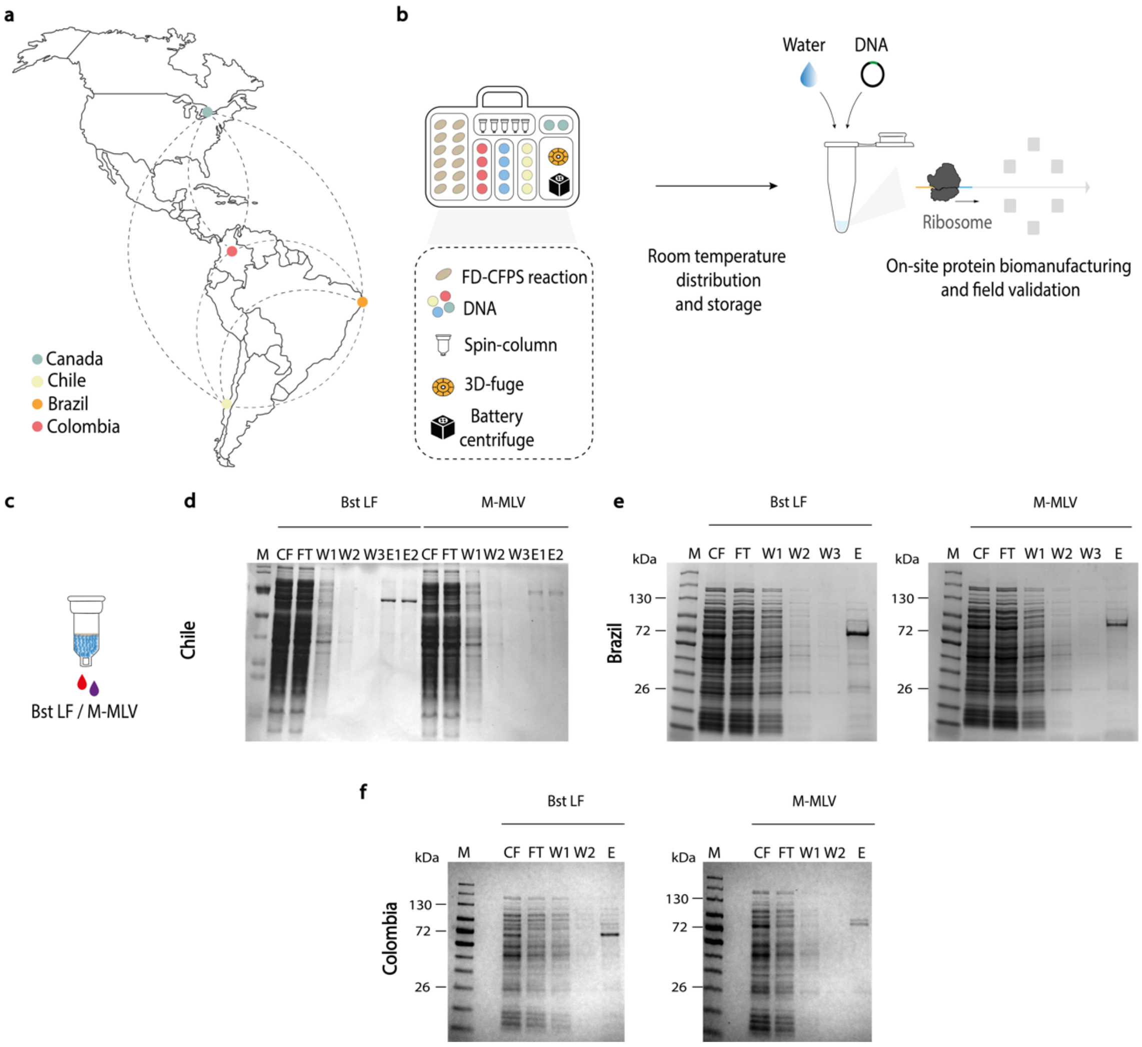

**Fig. S21: FD-CFPS enabled decentralized production of high-value diagnostic enzymes under minimal infrastructure conditions across diverse settings. (a,b)** Cell lysates were produced locally in basic low-containment microbiology laboratories at sites in North or South America. Here, CFPS reactions were either used at the production site or freeze-dried and distributed to multiple locations at ambient temperature (Chile, Brazil, and Colombia). **(c)** At each site, a straightforward one-day protocol enabled the local production of Bst LF and M-MLV enzymes from FD-CFPS reactions (500 µL, at ambient temperature). After overnight expression, the enzymes were purified using centrifugation-based affinity chromatography (20-minute protocol). **(d,e,f)** Purified diagnostic enzymes were analyzed using 4–20% gradient SDS-PAGE and stained using ProBlue Safe, yielding products of the expected size. The molecular weight ladder (in kilodaltons) is shown on the left. Representative gels were obtained using diagnostic enzymes produced on-site in Chile, Brazil, and Colombia. After production, diagnostic enzymes were utilized to develop in-house molecular diagnostics for pathogen detection, laying the groundwork for decentralized diagnostic programs addressing endemic infections and local healthcare needs. Abbreviations are: FD, freeze-dried; CFPS, cell-free protein synthesis; CF, crude reaction; FT, flow-through; W1-3, washes; E, elution.

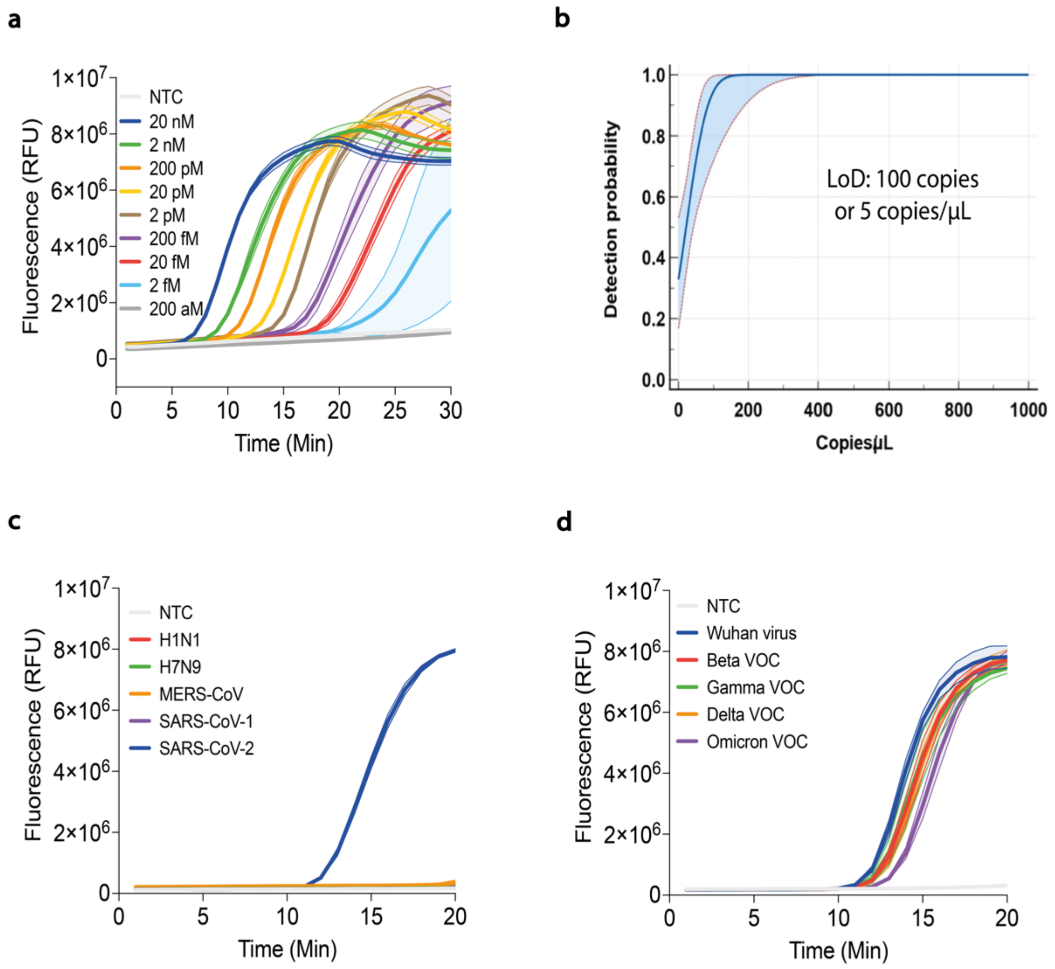

**Fig. S22: Optimized in-house reactions allowed precise detection of P. falciparum and SARS-CoV-2, achieving high sensitivity and specificity.** Prior to implementation and as part of the multi-site reproducibility assessment, we selected the diagnostic systems for *P. falciparum* (LAMP) and SARS-CoV-2 (RT-LAMP) for further evaluation. **(a)** Analytical sensitivity of the LAMP assay for *P. falciparum* detection was evaluated using a serial dilution of synthetic DNA. The assay detected the DNA target at clinically relevant concentrations (2 fM). Real-time fluorescence measurements were visualized using a conventional qPCR instrument with fluorescence reads every minute. Data are shown as mean ± SD, n = 3. **(b)** Similarly, analytical sensitivity of the RT-LAMP assay for SARS-CoV-2 detection was assessed using a serial dilution of synthetic RNA. Here, analytical sensitivity was determined using probit analysis (each concentration was tested 10 times), as previously described(*16*), confirming a sensitivity of 100 copies or 5 copies/µL. Real-time fluorescence measurements were visualized using a conventional qPCR instrument with fluorescence reads every minute. Data are shown as mean ± SD, n = 3**.** **(c,d)** Having confirmed the high sensitivity of the assay, subsequent experiments were conducted to evaluate the specificity of the in-house LAMP system for SARS-CoV-2 detection. Tests using a panel of related respiratory viruses, H1N1, H7N9, MERS-CoV, and SARS-CoV-1, demonstrated that the system was specific for SARS-CoV-2. In addition, an experiment confirmed the system's ability to detect all SARS-CoV-2 variants (alpha, beta, gamma, and delta), including the original Wuhan strain virus. Real-time fluorescence measurements were visualized using a conventional qPCR instrument with fluorescence reads every minute. Data are shown as mean ± SD, n = 3. Abbreviations are: NTC, non-template control; Min, minutes; VOC, variant of concern.

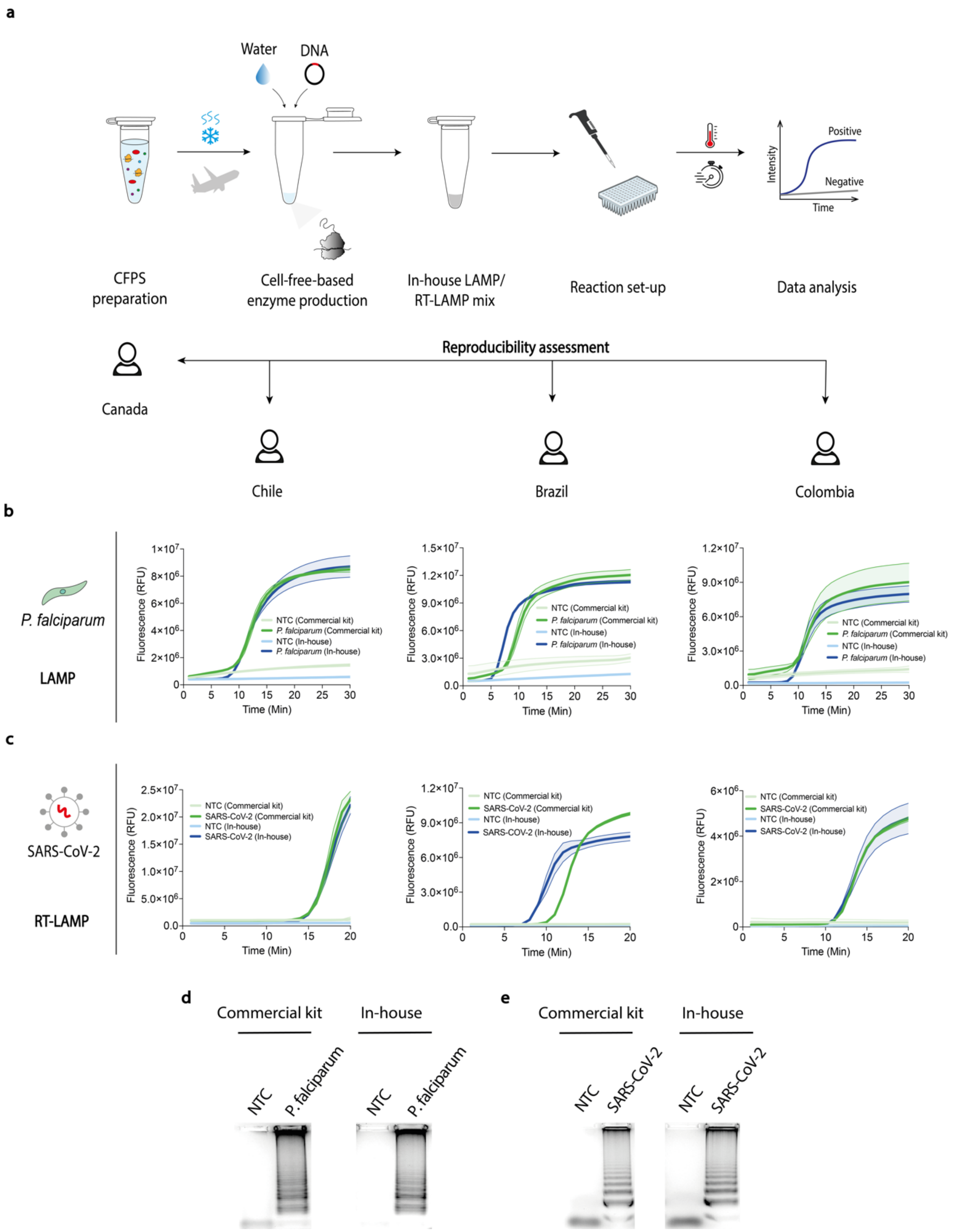

**Fig. S23: Standardized protocols and FD-CFPS reactions enabled consistent and reproducible results across laboratories with different resource levels, matching the performance of commercial reagents. (a)** CFPS reactions were freeze-dried (prepared in Toronto, Canada) and distributed to multiple locations at ambient temperature (Chile, Brazil, and Colombia). **(c)** At each site, a straightforward, one-day protocol enabled the on-site production of Bst LF and M-MLV enzymes from FD-CFPS reactions (500 µL reactions, at ambient temperature). After overnight expression, the enzymes were purified using centrifugation-based affinity chromatography. After local production of the diagnostic reagents, in-house LAMP/RT-LAMP reactions were assembled to detect

**(b)** *P. falciparum* and **(c)** SARS-CoV-2 targets. Here, team members independently performed the molecular assays using a standardized protocol, demonstrating high reproducibility across different settings. Real-time fluorescence measurements were visualized using a conventional qPCR instrument with fluorescence reads every minute. Data are shown as mean ± SD, n = 3. **(d,e)** In addition to fluorescence measurements, side-by-side experiments were performed using the colorimetric output, along with confirmation by agarose gel electrophoresis (1.5%). These assays further demonstrated comparable performance between in-house and commercial reactions to detect *P. falciparum* and SARS-CoV-2 nucleic acids. Data are presented from one representative biological replicate out of three independent experiments. Abbreviations are: NTC, non-template control; Min, minutes; CFPS, cell-free protein synthesis.

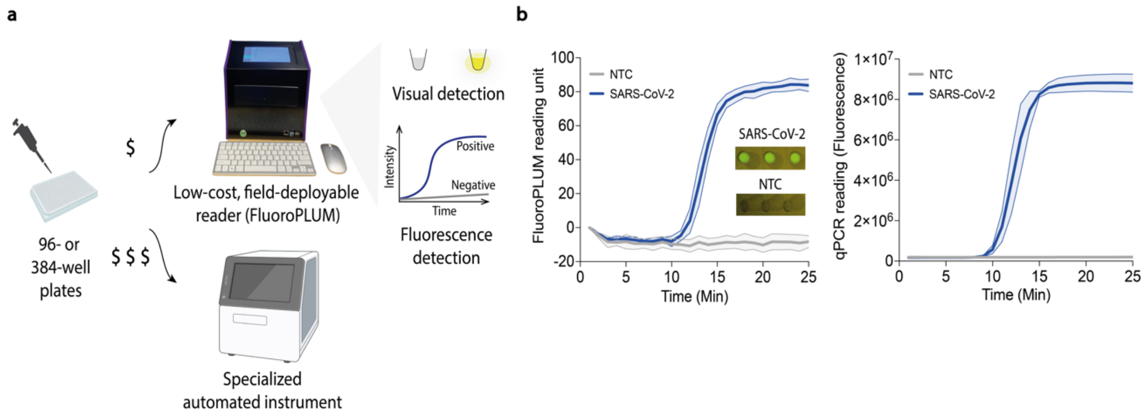

**Fig. S24: Low-cost, field-deployable FluoroPLUM delivered high-quality readouts comparable to a qPCR instrument. (a)** In-house RT-LAMP reactions were assembled and evaluated in parallel using the FluroPLUM, along with a qPCR instrument for comparison. **(b)** Real-time fluorescence measurements targeting SARS-CoV-2 synthetic RNA were performed simultaneously in both instruments, with fluorescence signals monitored every minute. For fluorescence measurements, amplicons were visualized by adding 1x LAMP fluorescent dye or 10 µM SYTO 9 Green Fluorescent Nucleic Acid dye if FluoroPLUM was used. The FluroPLUM also allowed for an alternative result readout, with endpoint fluorescence visible to the naked eye after incubation. Visual outputs are displayed within the designated graph area. Together, these findings demonstrate comparable performance across both platforms, confirming the utility of FluoroPLUM as an affordable alternative for diagnostic measurements in resource-limited settings. Data are shown as mean ± SD, n = 3. Abbreviations are: NTC, non-template control; Min, minutes.

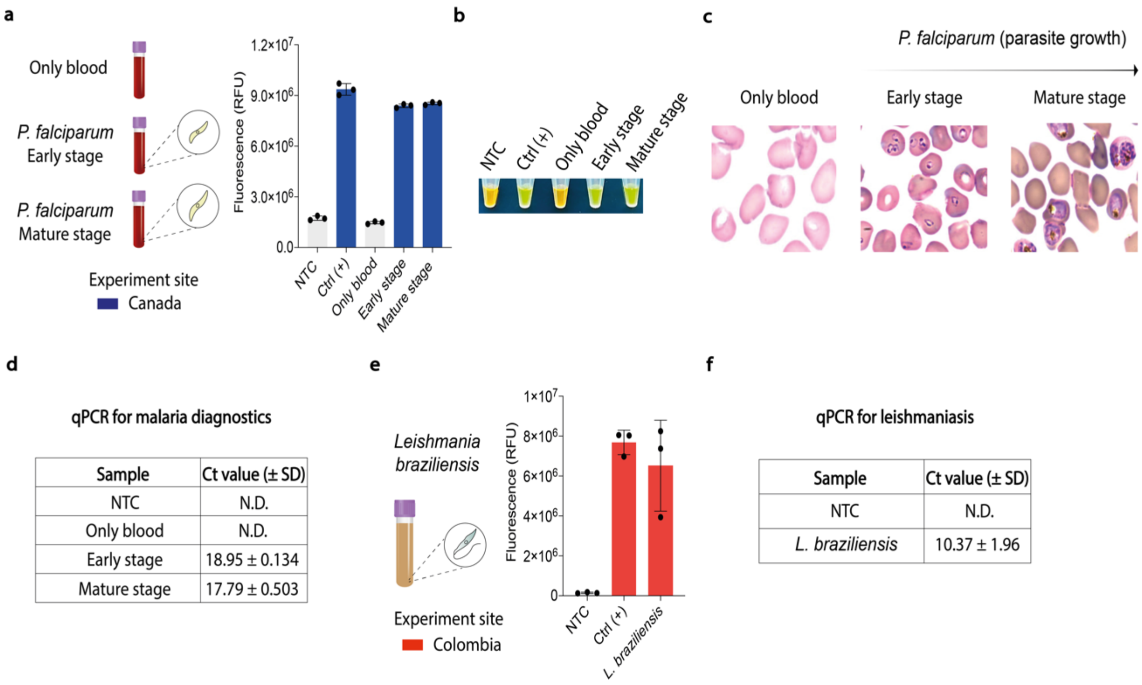

| Sample | Ct value (± SD) |
|---|---|
| NTC | N.D. |
| Only blood | N.D. |
| Early stage | 18.95 ± 0.134 |
| Mature stage | 17.79 ± 0.503 |

| Sample | Ct value (± SD) |
|---|---|
| NTC | N.D. |
| *L. braziliensis* | 10.37 ± 1.96 |

**Fig. S25: In-house LAMP assays reliably detected human parasites, with results validated against gold-standard methods.** Following the local production of diagnostic reagents, in-house LAMP reactions were assembled to detect human parasites using cultured-pathogen templates. **(a)** *P. falciparum* was cultured in its appropriate medium at both early and late stages of parasite growth, and the extracted DNA was used as input for LAMP reactions. Medium in the absence of parasites was used as a negative control. On-site-produced LAMP reactions containing 1X LAMP dye were used to detect *P. falciparum*, with fluorescence monitored in real-time using a conventional qPCR instrument to identify pathogen-positive samples. Fluorescence measurements after 30 minutes of incubation were plotted. Data are shown as mean ± SD, n = 3. **(b)** Using colorimetric outputs (measured at a 30-minute endpoint), in-house LAMP reactions also accurately detected *P. falciparum* across early and mature life stages. A positive sample was indicated by a colour change from orange to green, whereas a negative reaction stayed orange. Data are shown as mean ± SD, n = 3. Data shown are from one representative biological replicate out of three independent experiments. **(c)** Parallel testing by optical microscopy and **(d)** qPCR, used as gold-standard techniques, validated the in-house LAMP assays, achieving 100% concordance. This representative data was obtained using reagents produced on-site in Canada (blue data). **(e)** In South America, locally produced LAMP reactions detected *L. braziliensis*, a neglected parasite with substantial global health impact. Fluorescence measurements after 30 minutes of incubation were plotted. Data are shown as mean ± SD, n = 3. **(f)** Analysis of qPCR data obtained from cultured samples confirmed the presence of *L. braziliensis,* consistent with the results of the LAMP assay. Data are shown as mean ± SD, n = 3. This representative data was obtained using reagents produced on-site in Colombia (red data). Abbreviations are: NTC, non-template control; Ct, cycle threshold; N.D., not detected; Ctrl, control; SD, standard deviation.

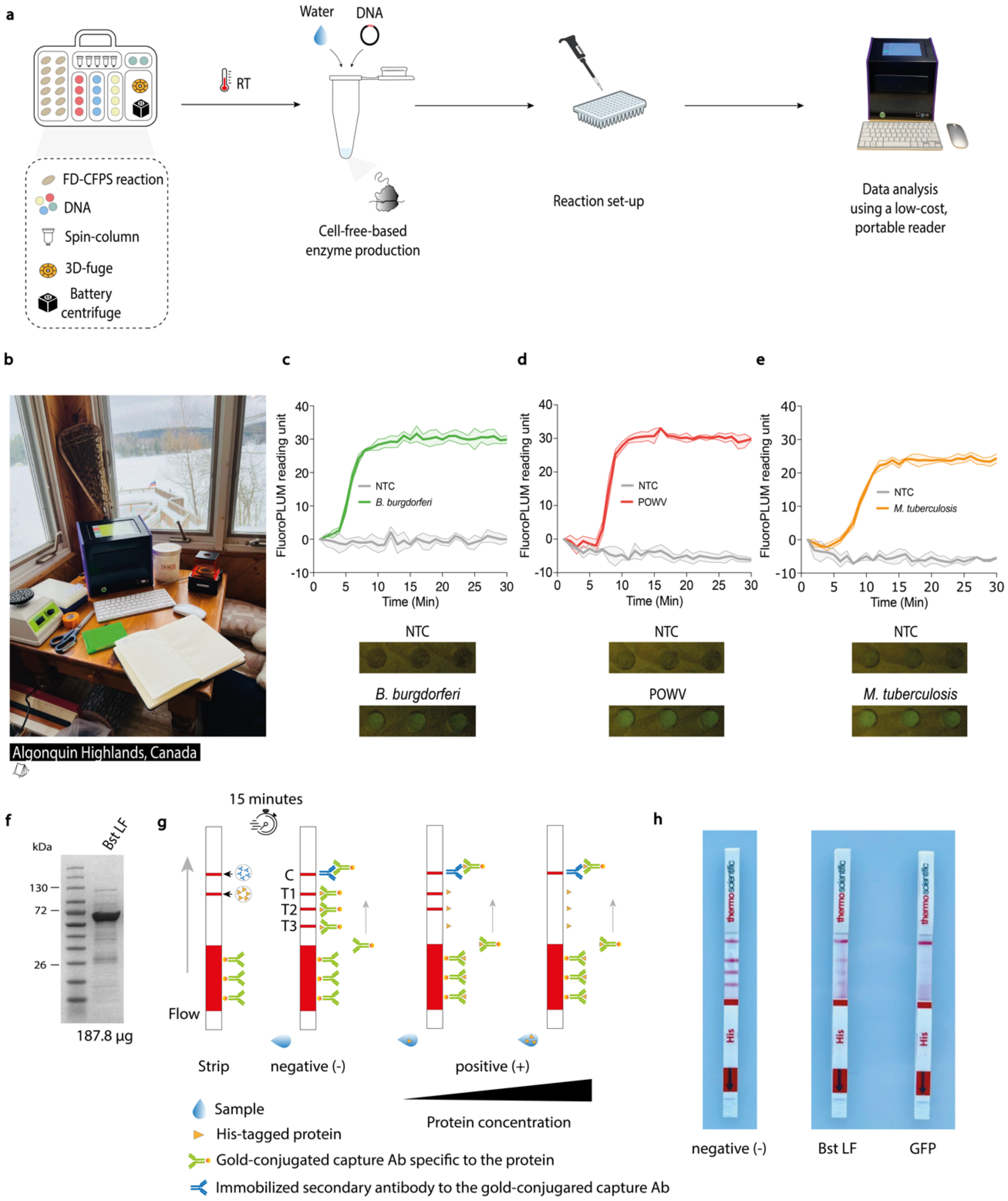

**Fig. S26: Low-burden FD-CFPS and portable, user-friendly hardware supported local enzyme manufacturing and molecular diagnostics in resource-limited settings. (a)** Molecular diagnostics for infectious diseases have traditionally depended on centralized laboratories, which are often equipped with specialized instruments, access to electricity, technical expertise, and commercial reagents. Following sample collection, specimens are transported to these facilities for processing and testing. While this approach has reliably supported diagnostic needs, supply chain interruptions and their high cost can significantly reduce diagnostic access, especially in resource-limited or remote settings. Here, we challenged the traditional paradigm of centralized reagent manufacturing and molecular diagnostics. Equipped only with essential materials, we successfully produced the diagnostic enzymes and reagents needed to perform molecular diagnostics in resource-limited settings. To meet this goal, FD-CFPS reactions, prepared in Toronto, Canada, were transported 250 km north to a rustic site in Algonquin Highlands (Ontario) and Whitehorse (Yukon) in Northern Canada—all chosen to simulate remote settings. **(b)** Once on-site, FD-CFPS reactions (0.5 mL) were first used to produce Bst LF and M-MLV (overnight at ambient temperature). Reactions were then purified using Ni-NTA resin columns with both battery-powered and hand-powered centrifuges. **(c-e)** After producing diagnostic inputs, in-house molecular reactions were assembled to detect synthetic nucleic acids (2 nM) of *B. burgdorferi* (green data)*,* POWV (red data), and *M. tuberculosis* (orange data). Without standard laboratory equipment, an enzyme titration assay was conducted to determine the optimal concentration for efficient amplification of molecular targets. With the system established, in-house molecular reactions were incubated in the FluoroPLUM using 10 µM SYTO 9 Green fluorescent nucleic acid dye, where fluorescence increases indicated successful amplification. Visual outputs obtained at the end of the reaction incubation (30 min) confirmed the detection of the corresponding pathogens. This representative data was obtained using diagnostic reagents manufactured on-site in Algonquin Highlands, Ontario, Canada. Data are shown as mean ± SD, n = 3. **(f)** Following field testing, eluates were returned to the lab and later confirmed through SDS-PAGE analysis and protein quantification assays. The molecular weight ladder (in kilodaltons) is shown on the left. **(g,h)** While field-based protein production represents a significant advancement, certain limitations remain. For instance, confirmation of protein expression typically depends on GFP reporter constructs expressed simultaneously, underscoring the need for complementary validation strategies. Alternatively, a His-tag lateral flow assay (Thermo Fisher Scientific, A38507) was tested as a simple method for protein expression confirmation in field settings where downstream analysis is unavailable. The assay reliably detected proteins produced under field conditions, supporting its use in decentralized biomanufacturing workflows. Abbreviations are: NTC, non-template control; Min, minutes; FD, freeze-dried; CFPS, cell-free protein synthesis; RT, room temperature; POWV, Powassan virus.

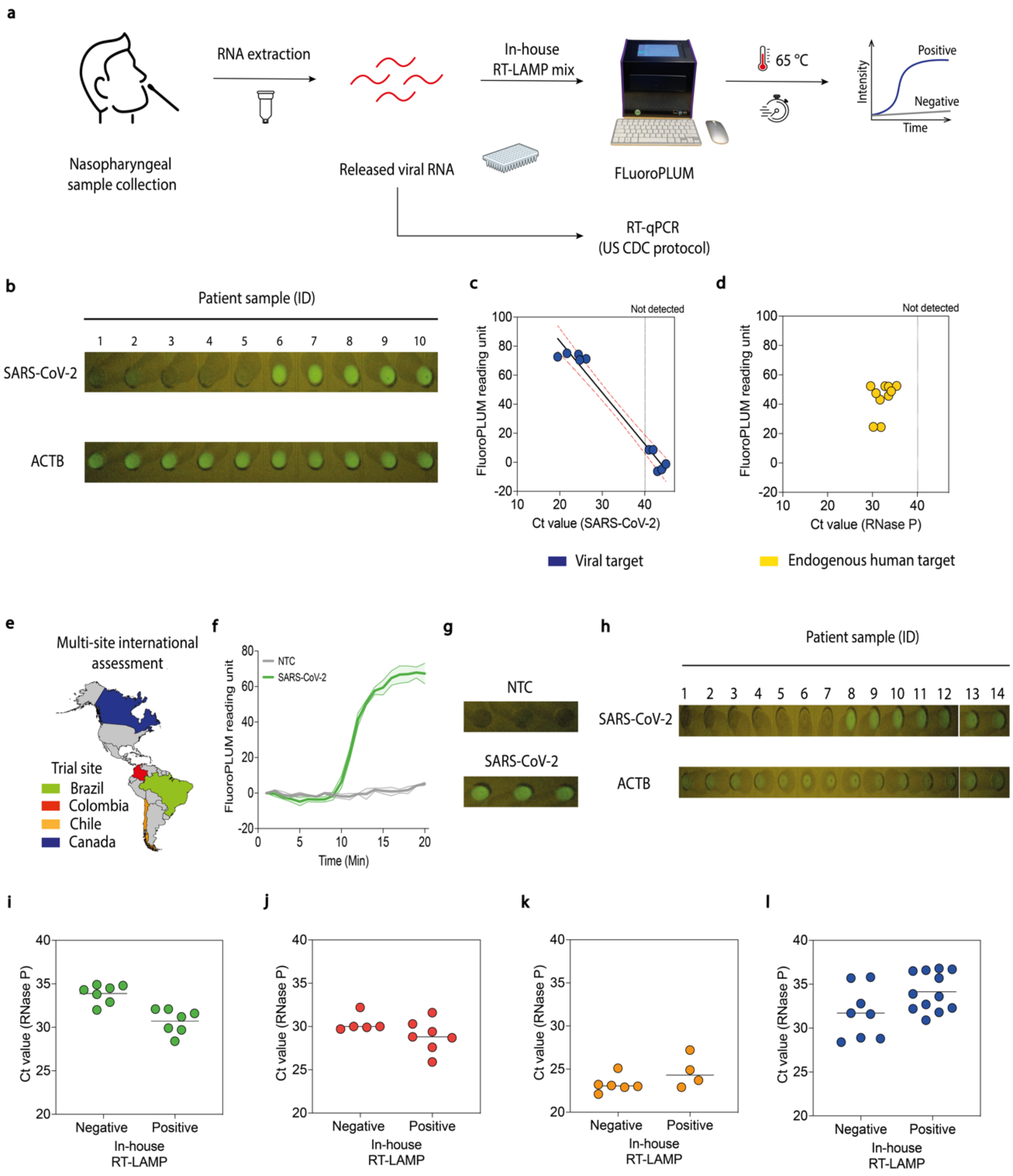

**Fig. S27: Locally produced diagnostic reagents enabled the implementation of COVID-19 testing programs in multiple countries, achieving performance comparable to the RT-qPCR. (a)** Before deploying the protocol across multiple countries, a straightforward and user-friendly diagnostic protocol was developed using locally produced reagents and the FluoroPLUM. Once established, a small-scale patient trial demonstrated reliable performance, achieving 100% concordance with RT-PCR results. In brief, a total of 10 extracted patient samples were tested using our in-house RT-LAMP system in parallel with the US CDC RT-qPCR protocol as the gold-standard comparison. RNA quality and integrity were confirmed in all patient samples using the human endogenous controls RNase P (RT-qPCR) and ACTB (RT-LAMP). **(b)** Visual outputs obtained at the end of the RT-LAMP incubation period, showing amplification of SARS-CoV-2 and ACTB targets. **(c,d)** Patient trial targeting SARS-CoV-2 (blue data) and RNAse P (yellow data) were conducted in Canada using locally produced, on-demand diagnostics. Samples were analyzed via RT-LAMP using 10 µM SYTO 9 Green fluorescent nucleic acid dye, where fluorescence increases indicated successful amplification. Fluorescence (y-axis) was plotted against the corresponding Ct values obtained using the RT-qPCR gold-standard assays (x-axis). The dashed line represents the threshold value defined for RT-qPCR analysis. **(e)** With diagnostic reagent production established at each site, the system was used to implement disease diagnostic programs in Brazil, Colombia, Chile, and Canada. Colours correspond to the data shown in subsequent panels, representing the countries where the data was collected. **(f,g)** Working in a national reference laboratory in Brazil, we initially tested our system using cultured SARS-CoV-2, with results showing successful amplification as indicated by both visual and real-time monitoring outputs. Data are shown as mean ± SD, n = 3**. (h)** End-point visual readouts of patient samples (14 samples) processed on the FluoroPLUM device in Brazil. **(i-l)** Following this initial phase, multi-site patient trials were carried out across different countries (Brazil, Colombia, Chile, and Canada) to validate the approach in clinical settings **(related to Fig. 7 c,h, Tables S6-12)**. Positive amplification of RNase P (Ct value, y-axis) and ACTB (diagnostic result, x-axis) was observed in all clinical samples tested across participating countries. This ensured quality control for this project phase and reinforced the reliability of viral target detection results. Abbreviations are: NTC, non-template control; Ct, cycle threshold; Min, minutes; ACTB, Actin Beta; RNase P, Ribonuclease P; ID, identification number.

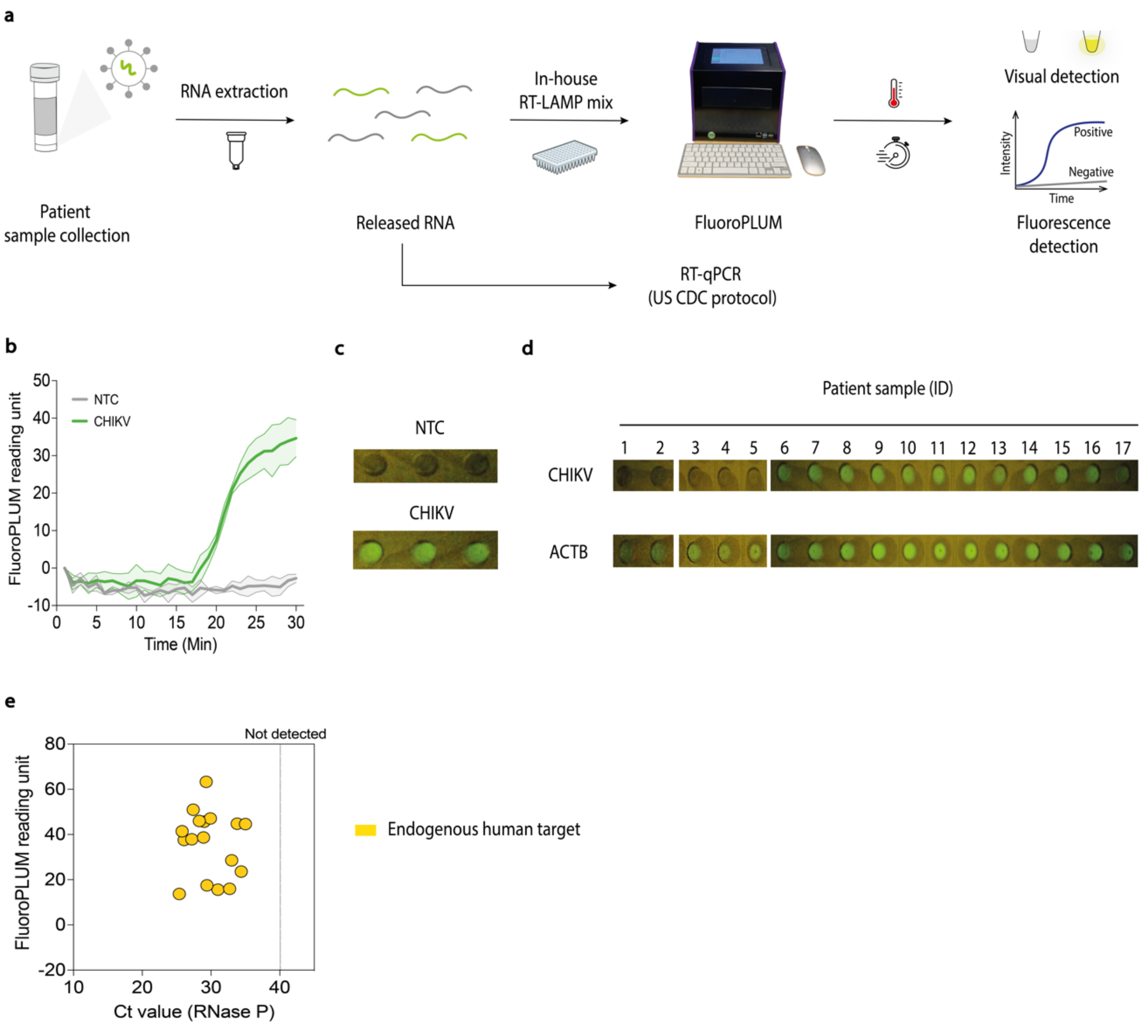

**Fig. S28: Implementation of low-cost molecular diagnostics for chikungunya virus detection in endemic infection areas using in-house inputs. (a)** With the local production protocol for reagents and diagnostic systems established at each site, we selected a few pathogens to initiate disease-focused clinical programs. Among these, we selected CHIKV because of its substantial public health impact in tropical and subtropical countries. **(b,c)** Working in Brazil, the epicenter of chikungunya epidemics in the Americas, we initially tested our in-house RT-LAMP system with cultured CHIKV, observing successful amplification through real-time monitoring and visual (30-minute endpoint) outputs. Data are shown as mean ± SD, n = 3**. (d)** End-point visual readouts of patient samples (17 samples) tested with in-house RT-LAMP reactions and visualized on the FluoroPLUM in Brazil **(related to Fig. 7e)**. **(e)** Side-by-side testing of 17 patient samples with RT-qPCR resulted in an accuracy of 100% **(related to Fig. 7e,h, Table S11)**. As part of sample verification quality, all samples tested positive for RNase P (Ct value, x-axis) and ACTB (FluoroPLUM reading unit, y-axis), confirming the high quality of the samples used for testing. Abbreviations are: NTC, non-template control; Ct, cycle threshold; Min, minutes; CHIKV, chikungunya virus; ACTB, Actin Beta; RNase P, Ribonuclease P.

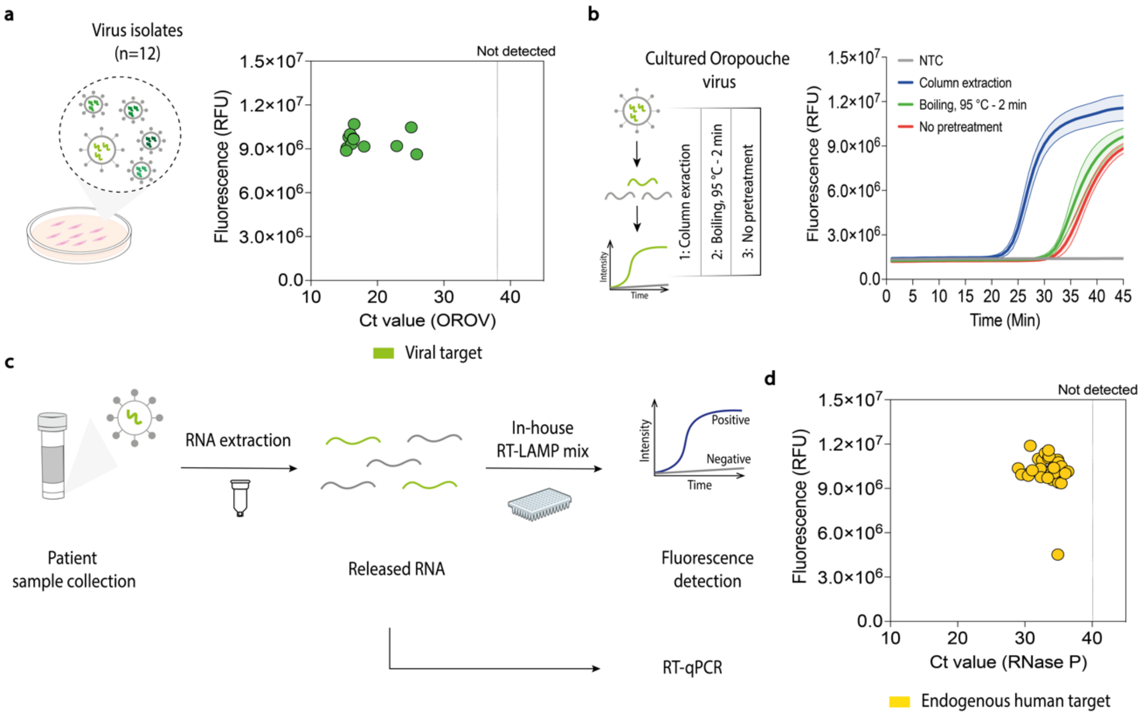

**Fig. S29: Building local biotechnology capacity through global partnerships enabled the rapid implementation of low-cost diagnostics in response to the emerging Oropouche virus in Latin America. (a)** As we worked to develop biotechnology capacity and local biomanufacturing in the countries involved in this initiative, the rapid emergence of the OROV as a public health threat offered a valuable opportunity to show how quickly and effectively local solutions can be mobilized to address urgent public health needs. In response, we rapidly developed an in-house RT-LAMP assay to detect this virus. Initial tests demonstrated the system's ability to detect 12 viral isolates obtained from patient samples collected at the beginning of the outbreak in Brazil. RNA samples isolated from virus isolates were analyzed via RT-LAMP using 1X LAMP fluorescent dye, where fluorescence increases indicated successful amplification. Fluorescence after 40 minutes (y-axis) was plotted against the corresponding Ct values obtained using the RT-qPCR gold-standard assays (x-axis). The dashed line represents the threshold value defined for RT-qPCR analysis. **(b)** Additional tests demonstrated the ability to detect OROV using a simple boiling step for viral lysis (e.g., 95 °C for 2 minutes) as well as directly without sample pretreatment **(related to Fig. 7f)**. Data are shown as mean ± SD, n = 3**. (c)** Having established the system with locally produced reagents, a patient trial was conducted using 33 serum samples collected from suspected cases of mosquito-borne infection in Brazil, the epicenter of the ongoing Oropouche epidemic in Latin America. **(d)** Side-by-side testing of 33 patient samples using RT-qPCR yielded 100% accuracy **(see Fig. 7g,h, and Table S12)**. Additionally, all samples showed positive results for RNase P (Ct value, x-axis) and ACTB (fluorescence, y-axis), confirming their high quality. Abbreviations are: NTC, non-template control; Ct, cycle threshold; Min, minutes; OROV, Oropouche virus; ACTB, Actin Beta; RNase P, Ribonuclease P.

**Supplementary Tables**

**Table S1. A summary of the centrifuges employed in this study.**

| Equipment | Speed | Price ($) USD |
|---|---|---|
| Benchtop laboratory centrifuge | Flexible (0-21000 x g) | ~10K |
| Battery centrifuge | 1200 x g | 149 |
| 3D-fuge | 1100 x g | 3 |

**Table S2. A summary of the growth factors used in this work.**

| Growth factor | Abbreviation | Functions | References |
|---|---|---|---|
| Fibroblast growth factor 1 | FGF-1 | Plays a role in embryonic development and tissue repair. In addition, plays crucial roles in normal and disease-related processes such as embryo development, tissue formation, blood vessel growth, wound healing, atherosclerosis, and cancer. | (*17, 18*) |
| Fibroblast growth factor 2 | FGF-2 | It is a highly specific chemotactic and mitogenic factor for numerous cell types, playing a role in tissue remodeling during healing processes such as ulcer repair, vascular regeneration, and recovery from traumatic brain injury. It is a heparin-binding cationic protein that plays a role in various pathological processes, such as angiogenesis and the growth of solid tumors. Additionally, it is an essential component of human embryonic stem cell culture medium. | (*19-21*) |
| Fibroblast growth factor 10 | FGF-10 | Demonstrates a wide range of roles in promoting cell division and survival, which are essential for many biological processes such as embryonic development, cellular proliferation, morphogenesis, tissue regeneration, as well as tumor growth and invasion. | (*22-24*) |
| Tumor necrosis factor alpha | TNF-α | It is a multifunctional molecule that regulates a broad range of biological processes, including cell proliferation, differentiation, | (*25, 26*) |

| | | | |
|---|---|---|---|
| | | apoptosis, lipid metabolism, and blood coagulation. Moreover, it is implicated in the defense against tumorigenesis. | |
| Interleukin-1 beta | IL-1β | It helps regulate inflammation, cell growth, and tissue repair. | (*27, 28*) |
| Interferon-gamma | IFN-γ | Exhibits antiviral, immunomodulatory, and antitumor functions. In addition, it plays a vital role in immune regulation. | (*29, 30*) |
| Interleukin-15 | IL-15 | Involved in the regulation of T cell and natural killer cell activation and expansion. It has been used in clinical trials for cancer treatment. | (*31, 32*) |
| Interleukin-6 | IL-6 | Regulates cell growth and differentiation, particularly in immune responses to specific pathogens. In addition, also plays a role in hematopoiesis, bone metabolism, and the progression of cancer. | (*33-35*) |
| Epidermal growth factor | EGF | Promotes proliferation, differentiation, and survival in certain cell types. Also, it plays a key physiological role in preserving the integrity of oro-esophageal and gastric tissues. | (*36-39*) |
| Insulin-like growth factor 1 | IGF-1 | Plays a crucial role in cellular growth and development and is involved in bone formation and metabolic regulation. Also, it stimulates the transport of glucose into cells. | (*40, 41*) |
| Interleukin-3 | IL-3 | Promotes growth and plays roles in cell proliferation, differentiation, and survival. In addition, it has neurotrophic effects, could be associated with neurological disorders, and is a key regulator of inflammation. | (*42-44*) |

**Table S3. Diagnostic performance of toehold-switch sensors for SARS-CoV-2 detection in Canadian patient samples (Ct ≤30).**

| | RT-qPCR - | RT-qPCR + | Total |
|---|---|---|---|
| **Biosensors -** | 6 | 1 | 7 |
| **Biosensors +** | 0 | 4 | 4 |
| **Total** | 6 | 5 | **11** |

| | |
|---|---|
| **Sensitivity** | 80.00% (95% CI 28.36% to 99.49%) |
| **Specificity** | 100.00% (95% CI 54.07% to 100.00%) |
| **Disease prevalence** | 45.45% (95% CI 16.75% to 76.62%) |
| **Positive Predictive Value (PPV)** | 100.00% (95% CI 39.76% to 100.00%) |
| **Negative Predictive Value (NPV)** | 85.71% (95% CI 50.97% to 97.19%) |
| **Accuracy** | 90.91% (95% CI 58.72% to 99.77%) |

**Table S4. Diagnostic performance of toehold-switch sensors for SARS-CoV-2 detection in Canadian patient samples (Ct ≤35).**

| | RT-qPCR - | RT-qPCR + | Total |
|---|---|---|---|
| **Biosensors -** | 6 | 2 | 8 |
| **Biosensors +** | 0 | 4 | 4 |
| **Total** | 6 | 6 | **12** |

| | |
|---|---|
| **Sensitivity** | 66.67% (95% CI 22.28% to 95.67%) |
| **Specificity** | 100.00% (95% CI 54.07% to 100.00%) |
| **Disease prevalence** | 50.00% (95% CI 21.09% to 78.91%) |
| **Positive Predictive Value (PPV)** | 100.00% (95% CI 39.76% to 100.00%) |
| **Negative Predictive Value (NPV)** | 75.00% (95% CI 49.18% to 90.29%) |
| **Accuracy** | 83.33% (95% CI 51.59% to 97.91%) |

**Table S5. Optimal temperature for our in-house LAMP/RT-LAMP systems.**

| | Pathogen | Temperature (°C) |
|---|---|---|
| **DNA targets** | *Borrelia burgdorferi* | 65 |
| | *Mycobacterium tuberculosis* | 65 |
| | *Plasmodium falciparum* | 60 |
| | *Leishmania donovani* | 64 |
| | *Leishmania braziliensis* | 64 |
| | Monkeypox virus | 63 |

| | Pathogen | Temperature (°C) |
|---|---|---|
| **RNA targets** | HIV-1 | 60 |
| | Chikungunya virus | 65 |
| | Zika virus | 65 |
| | Dengue virus-2 | 63 |
| | West Nile virus | 63 |
| | Mayaro virus | 65 |
| | SARS-CoV-2 | 65 |
| | Powassan virus | 65 |
| | H5N1 | 65 |
| | Oropouche virus | 65 |

**Table S6. Diagnostic performance of in-house RT-LAMP for SARS-CoV-2 detection in Brazilian patient samples.**

| | RT-qPCR - | RT-qPCR + | Total |
|---|---|---|---|
| **In-house RT-LAMP -** | 7 | 0 | 7 |
| **In-house RT-LAMP +** | 0 | 7 | 7 |
| **Total** | 7 | 7 | **14** |

| | |
|---|---|
| **Sensitivity** | 100.00% (95% CI 59.04% to 100.00%) |
| **Specificity** | 100.00% (95% CI 59.04% to 100.00%) |
| **Disease prevalence** | 50.00% (95% CI 23.04% to 76.96%) |
| **Positive Predictive Value (PPV)** | 100.00% (95% CI 59.04% to 100.00%) |
| **Negative Predictive Value (NPV)** | 100.00% (95% CI 59.04% to 100.00%) |
| **Accuracy** | 100.00% (95% CI 76.84% to 100.00%) |

**Table S7. Diagnostic performance of in-house RT-LAMP for SARS-CoV-2 detection in Colombian patient samples.**

| | RT-qPCR - | RT-qPCR + | Total |
|---|---|---|---|
| **In-house RT-LAMP -** | 5 | 0 | 5 |
| **In-house RT-LAMP +** | 0 | 7 | 7 |
| **Total** | 5 | 7 | **12** |

| | |
|---|---|
| **Sensitivity** | 100.00% (95% CI 59.04% to 100.00%) |
| **Specificity** | 100.00% (95% CI 47.82% to 100.00%) |
| **Disease prevalence** | 58.33% (95% CI 27.67% to 84.83%) |
| **Positive Predictive Value (PPV)** | 100.00% (95% CI 59.04% to 100.00%) |
| **Negative Predictive Value (NPV)** | 100.00% (95% CI 47.82% to 100.00%) |
| **Accuracy** | 100.00% (95% CI 73.54% to 100.00%) |

**Table S8. Diagnostic performance of in-house RT-LAMP for SARS-CoV-2 detection in patient samples from Chile.**

| | **RT-qPCR -** | **RT-qPCR +** | **Total** |
|---|---|---|---|
| **In-house RT-LAMP -** | 5 | 1 | 6 |
| **In-house RT-LAMP +** | 0 | 4 | 4 |
| **Total** | 5 | 5 | **10** |

| | |
|---|---|
| **Sensitivity** | 80.00% (95% CI 28.36% to 99.49%) |
| **Specificity** | 100.00% (95% CI 47.82% to 100.00%) |
| **Disease prevalence** | 50.00% (95% CI 18.71% to 81.29%) |
| **Positive Predictive Value (PPV)** | 100.00% (95% CI 39.76% to 100.00%) |
| **Negative Predictive Value (NPV)** | 83.33% (95% CI 46.42% to 96.65%) |
| **Accuracy** | 90.00% (95% CI 55.50% to 99.75%) |

**Table S9. Diagnostic performance of in-house RT-LAMP for SARS-CoV-2 detection in Canadian patient samples.**

| | RT-qPCR - | RT-qPCR + | Total |
|---|---|---|---|
| **In-house RT-LAMP -** | 7 | 1 | 8 |
| **In-house RT-LAMP +** | 0 | 12 | 12 |
| **Total** | 7 | 13 | **20** |

| | |
|---|---|
| **Sensitivity** | 92.31% (95% CI 63.97% to 99.81%) |
| **Specificity** | 100.00% (95% CI 59.04% to 100.00%) |
| **Disease prevalence** | 65.00% (95% CI 40.78% to 84.61%) |
| **Positive Predictive Value (PPV)** | 100.00% (95% CI 73.54% to 100.00%) |
| **Negative Predictive Value (NPV)** | 87.50% (95% CI 51.57% to 97.87%) |
| **Accuracy** | 95.00% (95% CI 75.13% to 99.87%) |

**Table S10. Diagnostic performance of in-house RT-LAMP for SARS-CoV-2 detection in Canadian patient samples.**

| | RT-qPCR - | RT-qPCR + | Total |
|---|---|---|---|
| **In-house RT-LAMP -** | 5 | 0 | 5 |
| **In-house RT-LAMP +** | 0 | 5 | 5 |
| **Total** | 5 | 5 | **10** |

| | |
|---|---|
| **Sensitivity** | 100.00% (95% CI 47.82% to 100.00%) |
| **Specificity** | 100.00% (95% CI 47.82% to 100.00%) |
| **Disease prevalence** | 50.00% (95% CI 18.71% to 81.29%) |
| **Positive Predictive Value (PPV)** | 100.00% (95% CI 47.82% to 100.00%) |
| **Negative Predictive Value (NPV)** | 100.00% (95% CI 47.82% to 100.00%) |
| **Accuracy** | 100.00% (95% CI 69.15% to 100.00%) |

*Note: Data collection using our low-cost diagnostic reader (FluoroPLUM).*

**Table S11. Diagnostic performance of in-house RT-LAMP for CHIKV detection in Brazilian patient samples.**

| | RT-qPCR - | RT-qPCR + | Total |
|---|---|---|---|
| **In-house RT-LAMP -** | 5 | 0 | 5 |
| **In-house RT-LAMP +** | 0 | 12 | 12 |
| **Total** | 5 | 12 | **17** |

| | |
|---|---|
| **Sensitivity** | 100.00% (95% CI 73.54% to 100.00%) |
| **Specificity** | 100.00% (95% CI 47.82% to 100.00%) |
| **Disease prevalence** | 70.59% (95% CI 44.04% to 89.69%) |
| **Positive Predictive Value (PPV)** | 100.00% (95% CI 73.54% to 100.00%) |
| **Negative Predictive Value (NPV)** | 100.00% (95% CI 47.82% to 100.00%) |
| **Accuracy** | 100.00% (95% CI 80.49% to 100.00%) |

**Table S12. Diagnostic performance of in-house RT-LAMP for OROV detection in Brazilian patient samples.**

| | RT-qPCR - | RT-qPCR + | Total |
|---|---|---|---|
| **In-house RT-LAMP -** | 12 | 0 | 12 |
| **In-house RT-LAMP +** | 0 | 21 | 21 |
| **Total** | 12 | 21 | **33** |

| | |
|---|---|
| **Sensitivity** | 100.00% (95% CI 83.89% to 100.00%) |
| **Specificity** | 100.00% (95% CI 73.54% to 100.00%) |
| **Disease prevalence** | 63.64% (95% CI 45.12% to 79.60%) |
| **Positive Predictive Value (PPV)** | 100.00% (95% CI 83.89% to 100.00%) |
| **Negative Predictive Value (NPV)** | 100.00% (95% CI 73.54% to 100.00%) |
| **Accuracy** | 100.00% (95% CI 89.42% to 100.00%) |
